# Impact of violence on HIV outcomes among female sex workers: A global systematic review and meta-analysis

**DOI:** 10.64898/2026.02.27.26346881

**Authors:** Joshua Dawe, Khadimul Anam Mazhar, Saher Aijaz Khan, Belinda Jackson Njiro, Victoria Bendaud, Keith Sabin, Julie Ambia, Adam Trickey, Lucy Barrass, Asra Asgharzadeh, Jack Stone, Adelina Artenie, Peter Vickerman

**Affiliations:** Bristol Population Health Science Institute, Bristol Medical School, Bristol, England; Health Protection Research Unit, Bristol Medical School, Bristol, England; Data for Impact, The Joint United Nations Program on HIV/AIDS (UNAIDS), Geneva, Switzerland; Department of Family and Emergency Medicine, Faculty of Medicine, University of Montreal; Centre de recherche du Centre hospitalier de l’Université de Montréal, Montreal, Canada

## Abstract

**Background:** Female sex workers (FSW) are a key population for HIV prevention and care. Increasing evidence suggests that social and structural barriers are key drivers of HIV transmission. This global systematic review assesses whether experiencing violence is associated with worse HIV outcomes among FSW.

**Methods and Findings:** We searched MEDLINE, Embase, and PsycINFO databases for studies published from January 1^st^, 2010 to February 10^th^, 2025 assessing the impact of violence on HIV outcomes among FSW, without restriction to language and study design. Some studies had multiple estimates due to reporting on multiple outcomes or exposures of interest. We pooled data from eligible studies using multi-level random-effects meta-analyses to quantify associations between recent (past year) or lifetime exposure to violence (physical, sexual, emotional/psychological and/or financial) and HIV outcomes (prevalent and incident HIV infection, HIV testing, ART use, ART adherence, and viral suppression) among FSW. We preferentially used adjusted estimates over unadjusted estimates if both were available.

We included 91 studies with 221 estimates, comprising 179,727 FSWs in 37 countries. We found higher odds of prevalent HIV infection among FSWs with recent (pooled odds ratio (pOR):1.33; 95%CI:1.17–1.51; I^2^:64%; n=73 estimates) and lifetime (pOR:1.36; 95%CI:1.24–1.49; I^2^:38%; n=67) experiences of violence. Recently experiencing violence was associated with reduced odds of ART use (pOR:0.78; 95%CI:0.64–0.94; I^2^:8%; n=17). Lifetime exposure to violence was associated with reduced odds of viral suppression (pOR:0.88; 95%CI:0.79–0.98; I^2^:20%; n=6). There was no evidence of associations between violence and HIV incidence, HIV testing and ART adherence.

**Conclusions:** Experiencing violence may increase HIV transmission risk and worsen HIV treatment outcomes among FSW. HIV interventions for FSWs must address violence as a structural determinant of HIV.

## Introduction

Despite ongoing global efforts, the HIV epidemic continues to have considerable global health impacts, particularly in lower- and middle-income countries (LMICs).^1^ Whilst HIV incidence has declined in many countries, new infections are increasingly concentrated among key populations, such as female sex workers (FSWs).^2^ In 2022, a global modelling analysis estimated that FSWs had nine times higher risk of acquiring HIV than the adult male and female population, and 18% of incident HIV infections globally occurred among FSWs and their clients.^3, 4^ Furthermore, evidence suggests that in many settings inequities exist in the HIV cascade of care among FSWs.^5^

The reasons for heightened levels of HIV infection and poorer treatment outcomes among FSWs compared with the general population are multifaceted, resulting from complex interplays between societal, behavioural, and other factors.^6^ These include structural factors outside an individual’s control, which influence the risk environment in which FSWs live and work, such as the criminalisation of sex work, stigma and discrimination, and violence.^7^

In 2020, UNAIDS set global targets for reducing exposure to adverse structural factors among key populations, including reducing rates of violence among sex workers to less than 10% per year.^8^ However, evidence suggests rates of violence among sex workers exceeds this target in most countries^9^, with one in four sex workers globally estimated to experience sexual or physical violence annually.^10^ Studies have described how violence could increase HIV transmission risk, either directly through violent sexual exposure^11^ or indirectly through increasing condomless sex, substance use or reducing access to healthcare and HIV interventions.^12–15^ Through these effects, mathematical modelling has suggested that reducing violence could prevent up to one-third of HIV infections among FSWs and their clients in some settings.^16, 17^

Despite accumulating studies suggesting detrimental effects of violence on HIV outcomes among FSWs, no systematic review has quantitatively synthesised this evidence, and so there is uncertainty in its effects. Improving our understanding of the effects of violence among FSWs is crucial for optimising and tailoring HIV interventions for this population. We therefore undertook a global systematic review and meta-analysis to characterise the effects of violence on specific HIV outcomes (HIV testing, ART use, ART adherence, viral suppression) among FSWs.

## Methods

This systematic review and meta-analysis is reported according to the 2020 preferred reporting items for systematic reviews and meta-analyses (PRISMA) checklist^18^, following a registered study protocol PROSPERO (ID: CRD42024531539). The review was conducted in unison with another review, which aimed to estimate the effects of stigma and discrimination on the same HIV outcomes among FSWs. The completed PRISMA checklist is included in Appendix 2.

### Information sources and eligibility criteria

We searched MEDLINE, Embase, and PsycINFO for published studies from January 1^st^, 2010 to February 10^th^, 2025, without language restrictions. Eligible study designs included those that could be used to estimate associations between any form of violence and one or more selected HIV outcomes among FSWs, namely observational studies (and specifically, case-control studies, cohort studies, cross-sectional studies), and intervention studies including randomised controlled studies. We included studies in which FSWs were the main population under investigation, with a rule that greater than 70% of the included sample needed to have recently/ever engaged in sex work. Studies also required a measure of association, or provided sufficient information to allow calculation of an association, between violence and specific HIV outcomes, including HIV prevalence, HIV incidence, HIV testing uptake, ART use, ART adherence, and HIV viral suppression. For HIV testing, we included studies of recent and lifetime HIV testing among FSWs. Definitions of HIV incidence included new diagnoses among FSWs with a prior negative HIV test and biomarkers of recent infection from avidity assays. We defined ART use as current use of ART among FSWs living with HIV, or ART initiation among FSWs with a recent HIV diagnosis. We defined ART adherence as consistent use of ART (proportion of doses taken) within a specified time frame (e.g. past month). We included both self-reported and laboratory-based measures for ART use and ART adherence. HIV viral suppression was defined using the operational definition specified by the study authors (<50 copies/mL to <1000 copies/mL), with the denominator either being FSWs living with HIV or FSWs prescribed ART. If multiple studies reported the same measure of association from the same sample of FSWs, we included the most recently published study. Studies were excluded if they had a sample size below 40.

### Search strategy

Structured search terms were designed to identify articles reporting an association between violence and our HIV outcomes of interest among FSWs. To ensure we captured as many eligible studies as possible, we used detailed search terms (Appendix 3) related to female sex work and structural factors and kept the HIV search terms deliberately broad. After searching MEDLINE, Embase and PsycINFO, deduplication of titles and abstracts was conducted in Endnote using previously published methods.^19^ Title and abstract screening and full-text review were conducted in duplicate by four authors (JD, KM, SK, BN) using Rayyan AI^20^, with discrepancies resolved through discussion. Two authors (JD and BN) manually searched the reference lists of systematic reviews identified in the initial search for additional relevant studies.

### Data extraction

Data extraction was conducted by one of four authors (JD, KM, SK, BN) using Microsoft Excel. This was double-checked (JA, AT, AA, JD, BN, LB, JS) for consistency. Violence was grouped into three categories: physical violence, sexual violence and other forms of violence. Few studies reported on other forms of violence, and these measures were heterogeneous (e.g., emotional, psychological), therefore, they were grouped into a single “other violence” category. Each violence measure was categorised as recent (up to past 12 months), or lifetime. Data extracted included publication type (manuscript, conference abstract, PhD dissertation), publication year, study design, study setting, recruitment method, country, definition of sex work, time frame for sex work, fraction exposed, fraction living with HIV, sample size (overall and analytical sample), violence type and definition, and the perpetrator of violence. We extracted all associations between violence and our HIV outcomes, irrespective of the type of measure of association (i.e. odds ratio (OR), prevalence ratio (PR), risk ratio (RR), as available in each study), and whether unadjusted or adjusted for confounding factors; if both unadjusted and adjusted estimates were available, we extracted both. We manually calculated ORs and the corresponding 95% confidence intervals (95%CI) for studies where sufficient information was available, but an association was not reported. Since OR were used in most included estimates, where possible we converted unadjusted associations to ORs for pooling for all HIV outcomes other than HIV incidence. If physical violence, sexual violence and other forms of violence were not differentiated in an estimate, the estimate was extracted as mixed violence. For example, if a study reported violence as “Physically beaten or raped”, the estimate was categorised as mixed violence.

### Analysis

Analyses were carried out separately for each outcome. In our main analyses, we computed pooled estimates combining all types of violence while separating by recent and lifetime exposure. We used adjusted estimates when available, otherwise using unadjusted estimates. In supplementary analyses, we also pooled unadjusted and adjusted estimates separately. Where there were three or more studies, we also pooled estimates separately for different violence types. Given that several studies reported multiple estimates for the same outcome (for example, separate associations for physical and sexual violence, or violence for different perpetrators), we estimated the pooled effects using correlated and hierarchical effects models with robust variance estimation, which account for the variation between studies, and the correlation between effect estimates for studies with multiple effects. We conducted this analysis using the clubSandwich R package as described in Pustejovsky et al.^21^ As per Pustejovsky et al., we assumed a constant sampling correlation of p = 0.6. We assessed the robustness of this assumption by conducting sensitivity analyses using constant sampling correlations of p = 0.4 and p = 0.8. We also calculated the pooled proportion of FSWs exposed to each type of violence (separately for recent and lifetime) using multi-level random-effects meta-analysis.

Heterogeneity between studies was assessed using the I² statistic. We assessed publication asymmetry for our combined pooled effects for all combinations of violence exposures and HIV outcomes using a funnel plot and Egger’s test. Subgroup analyses for key variables included in our data extraction were used to identify possible sources of heterogeneity (Appendix 8). We conducted subgroup analyses by estimating and comparing the pooled effects for each stratum of subgroup variables, using adjusted estimates when available, otherwise using unadjusted estimates. Variables included WHO region, World Bank country income level classification (lower-income and lower-middle income, upper-middle income and upper income), year of publication (before 2017, 2017 and after), recruitment method (respondent-driven sampling, time-location sampling, convenience sampling, and probabilistic sampling), and the perpetrator of violence (clients, employers, intimate partners, family or friends, police, healthcare workers, unspecified).

Study quality was assessed for each exposure-outcome estimate pair across three domains; study representativeness (representative random sample of FSWs vs. not), study design (longitudinal vs. cross-sectional) and whether estimates were adjusted or not (adjusted vs. unadjusted). Each domain was scored zero or one; the maximum score per estimate was three. A full outline of the study quality assessment is included in Appendix 5.

## Results

Following database searches, we identified 6,954 potentially eligible studies published between January 1^st^ 2010 and February 10^th^ 2025, of which 1,786 (25.7%) were duplicates (Figure 1). After screening 5,168 titles and abstracts, 211 (4.1%) studies were eligible for full-text screening, of which 87 (41.2%) studies met the inclusion criteria. Of these, five studies were excluded due to publishing duplicate measures of association from the same sample. After manually searching the reference lists of relevant systematic reviews found during the search (N=56), nine more studies were identified (Appendix 5), resulting in a total of 91 studies that reported associations between experiences violence and our HIV outcomes. (Appendix 4).

**Figure 1.**
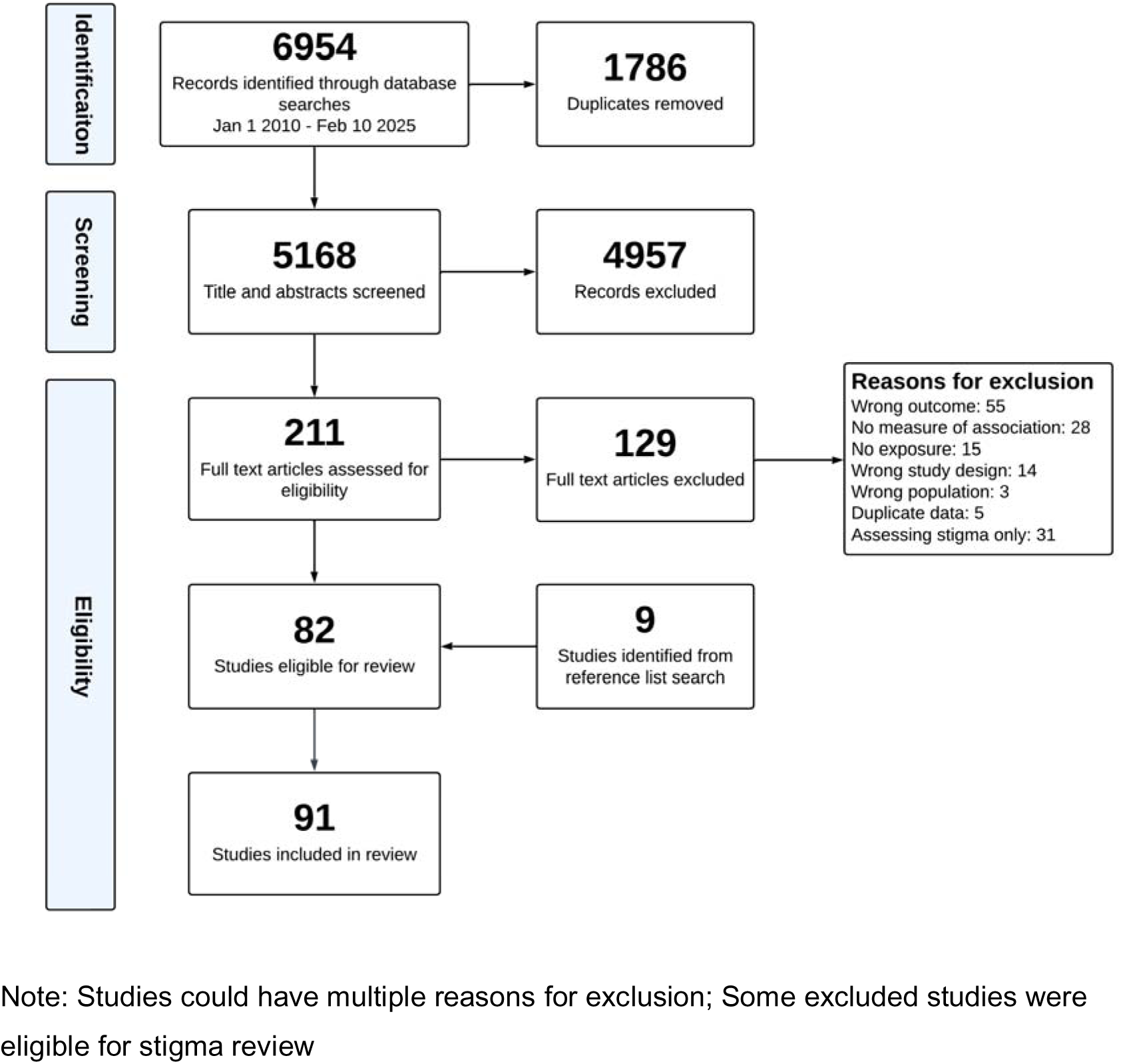
PRISMA flowchart.

Among 91 included studies, 87 were cross-sectional and four were longitudinal; 89 were journal articles, one was a conference abstract and one was a PhD dissertation (Table 1). These studies comprised 179,727 FSWs in 37 countries. Slightly more than half of the included studies were from the African region (n=50, 54.9%). Three-quarters of included studies were considered representative (n=68, 74.7%). Around half of the included estimates adjusted for confounding (n=109, 49.3%), and most estimates were cross-sectional (n=215, 97.3%). In one study, fewer than 100% of participants reported ever engaging in sex work (74%)^22^, and in all other studies 100% reported recent or lifetime sex work.

**Table 1.** Summary table of included studies.

| Characteristic | n (%) |
| --- | --- |
| Number of studies | 91 |
| Study design |  |
| Cross-sectional | 87 (96.6) |
| Longitudinal | 4 (4.4) |
| Publication status |  |
| Journal article | 89 (97.8) |
| Conference abstract | 1 (1.1) |
| PhD dissertation | 1 (1.1) |
| WHO region |  |
| African region | 50 (54.9) |
| Region of the Americas | 17 (18.7) |
| South-East Asian region | 14 (15.4) |
| European region | 6 (6.6) |
| Eastern Mediterranean region | 3 (3.3) |
| Western Pacific region | 1 (1.1) |

### Exposure to violence

Many studies reported more than one association between a measure of exposure to violence and an eligible HIV outcome, with 221 estimates available from the 91 included studies. Thirty-three studies reported estimates of association for exposure to physical violence (55 estimates), 52 reported associations for sexual violence (70 estimates), 39 reported mixed forms of violence (73 estimates), and ten reported associations for other forms of violence (23 estimates). There was heterogeneity in the measures used for each form of violence (Appendix 6).

Among studies reporting recent exposure to violence (n=55), the pooled proportion of FSWs exposed to physical, sexual, and mixed violence in past 12 months was 20.2% (95%CI:14.3%–27.7%, I^2^:98.1%; 18 studies; 21 estimates), 15.0% (95%CI:10.1%–21.6%, I^2^:98.7%; 25 studies; 29 estimates) and 21.9% (95%CI:17.0%–27.7%, I^2^:96.1%; 22 studies; 26 estimates), respectively. For studies reporting lifetime exposure (n=45), the pooled proportions of FSWs exposed to physical, sexual, and mixed violence were 32.2% (95%CI:25.7%–39.5%, I^2^:97.2%; 14 studies; 22 estimates), 24.3% (95%CI:19.1%–30.3%, I^2^:98.1%; 26 studies; 30 estimates) and 48.3% (95%CI:35.2%–61.7%, I^2^:99.4%; 12 studies; 13 estimates), respectively. In pooled proportions of other types of violence (not physical or sexual violence), 35.4% (95%CI:24.3%–48.4%, I^2^:96.7%; 7 studies; 9 estimates) of FSWs were recently exposed, and 47.4% (95%CI:35.8%–59.3%, I^2^:94.3%; 4 studies; 5 estimates) were ever exposed.

### Associations between violence and prevalent HIV infection

Our main analysis showed that FSWs with recent experiences of any type of violence have 1.33-fold (95%CI:1.17–1.51; I²:64%; 40 studies; 73 estimates) higher odds of living with HIV compared to those without recent experiences of violence (Figure 2). When stratified by different violence types, the greatest increase in the odds of living with HIV was for recent experiences of sexual violence (pOR: 1.61; 95%CI:1.27–2.04; I^2^:68%; 21 studies; 27 estimates); pooled effect estimates for mixed violence (pOR: 1.26; 95%CI:1.06–1.50; I^2^:56%; 14 studies; 19 estimates) physical violence (pOR: 1.20; 95%CI:0.98–1.47; I^2^:65%; 13 studies; 21 estimates) and other forms of violence (pOR: 1.15; 95%CI:0.77–1.71; I^2^:59%; 5 studies; 6 estimates) were more modest and for physical violence and mixed violence the 95%CI included the null.

**Figure 2.**
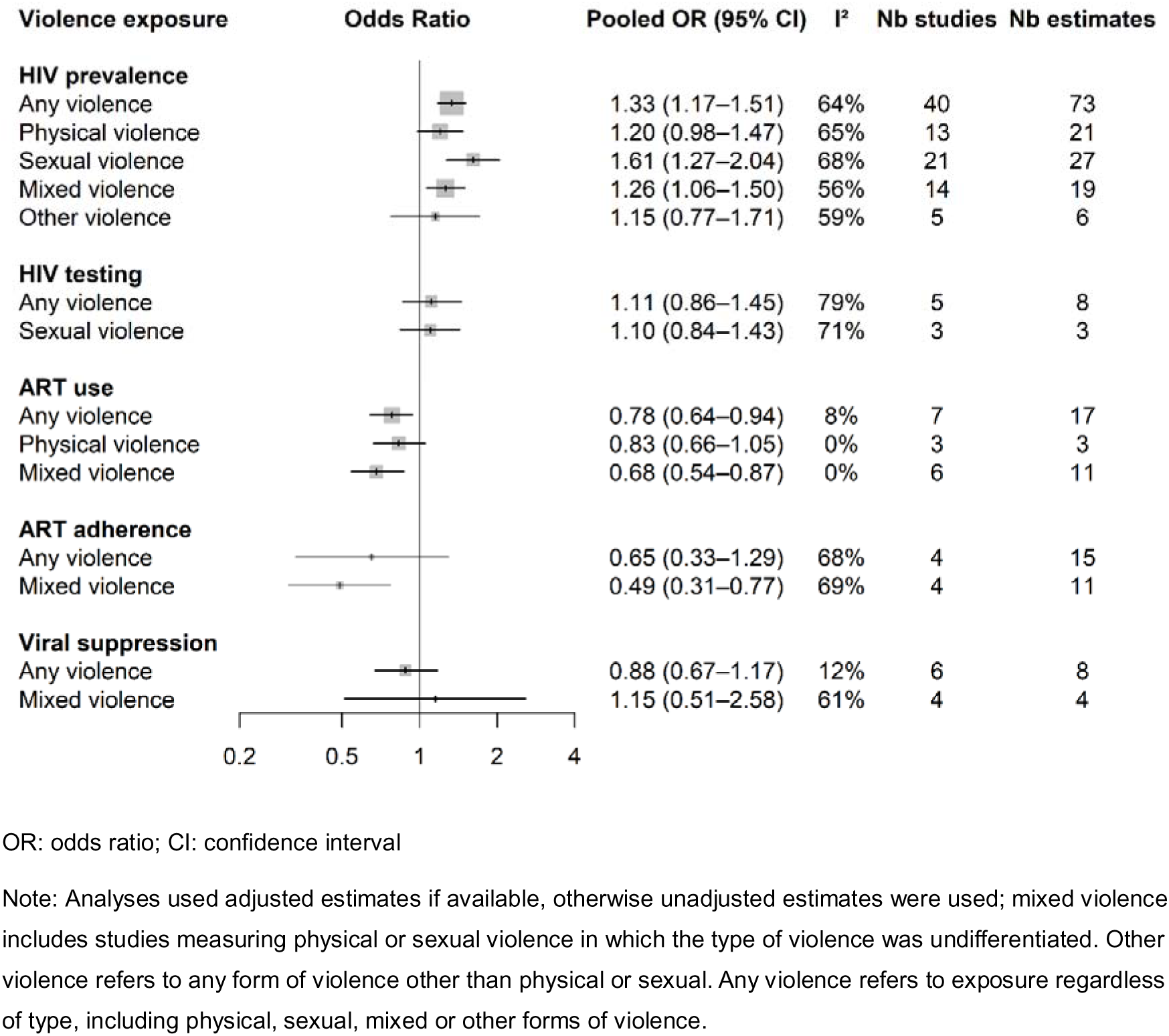
Pooled odds ratios reflecting associations between recent exposure to different forms of violence and prevalent HIV infection and the HIV cascade of care among female sex workers, stratified by HIV outcome

Analyses of lifetime exposure to violence also found strong associations between any type of violence and living with HIV among FSWs (pOR: 1.36; 95%CI:1.24–1.49; I^2^:38%; 35 studies; 67 estimates) (Figure 3). In stratified analyses, there were similar elevated odds for lifetime exposure to physical (pOR: 1.29; 95%CI:1.15–1.44; I^2^:0%; 14 studies; 21 estimates), sexual (pOR: 1.40; 95%CI:1.21–1.63; I^2^:47%; 24 studies; 28 estimates) and mixed violence (pOR: 1.32; 95%CI:1.15–1.52; I^2^:38%; 11 studies; 16 estimates). Heterogeneity varied across analyses, with some pooled estimates showing no heterogeneity and others showing moderately-high levels. There were only two estimates of the association between lifetime experiences of other violence and living with HIV, so this was not pooled.

**Figure 3.**
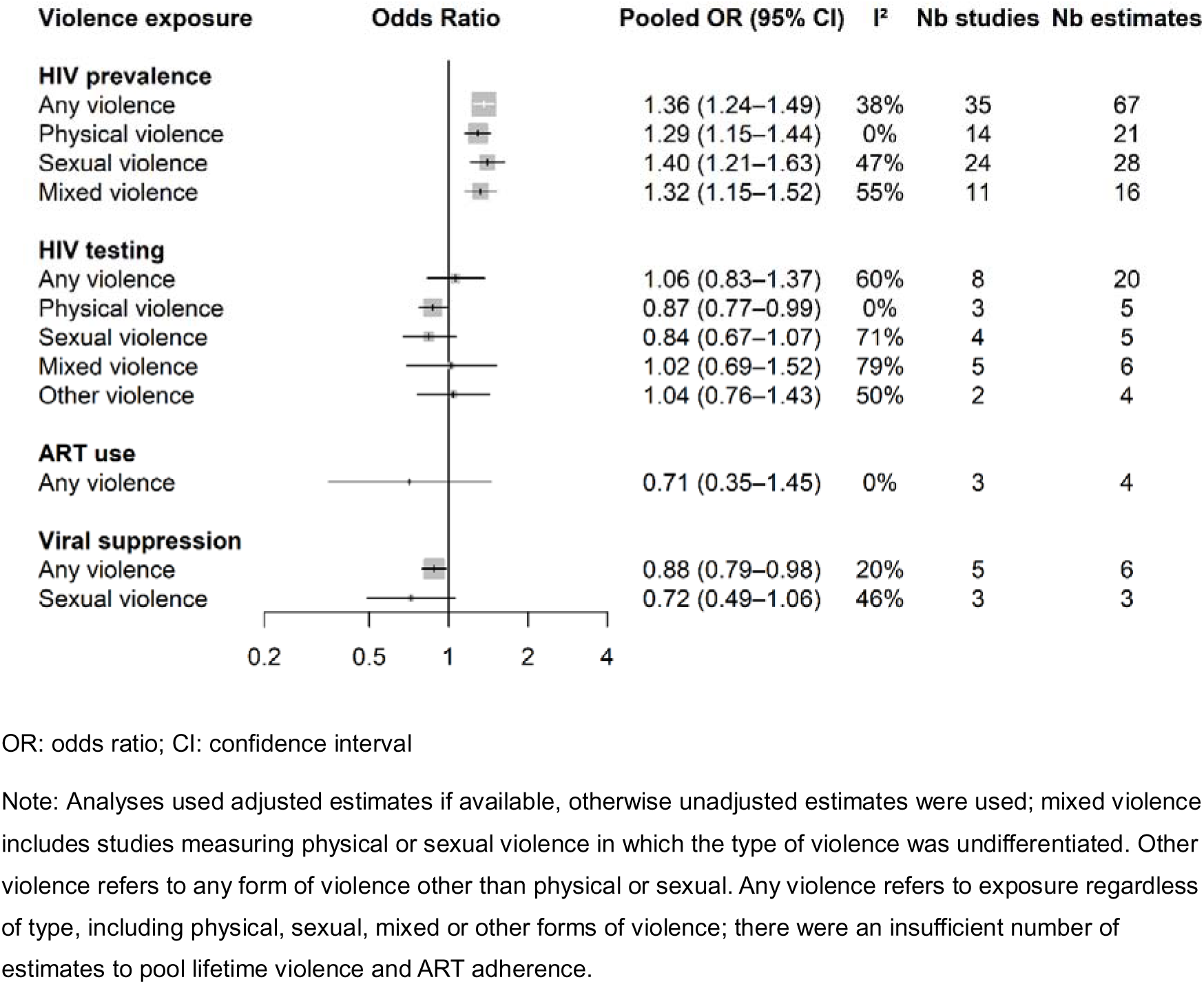
Summary of pooled associations between lifetime exposure to different forms of violence and prevalent HIV infection and the HIV cascade of care among female sex workers, stratified by HIV outcome

### Associations between violence and HIV incidence

Three studies reported an association between violence and HIV incidence, with one study measuring incidence longitudinally and two studies using a recency assay. One study reported exposure to recent sexual violence, one reported exposure to lifetime sexual violence and one reported exposure to lifetime mixed violence. Because <3 studies for either recent or lifetime violence exposure were available, these could not be pooled. In all three studies, the association between violence and HIV incidence was inconclusive.

### Associations between violence and HIV testing

We included 13 studies which reported associations with HIV testing. Four studies reported lifetime HIV testing, and nine studies reported recent HIV testing, with timeframes ranging from past three months to past two years. We found no evidence of an association between recent experiences of any form of violence and HIV testing among FSWs in pooled analyses (pOR: 1.11; 95%CI:0.86–1.45; I^2^:79%; 5 studies; 8 estimates) (Figure 2). This was consistent with lifetime experiences of all forms of violence, with no evidence of an association in pooled analyses (pOR: 1.06; 95%CI:0.83–1.37; I^2^:60%; 8 studies; 20 estimates) (Figure 3). Findings were similar when stratified by violence type, with most estimates not showing an association with HIV testing. However, there were lower odds of HIV testing among FSWs reporting lifetime experiences of physical violence (pOR: 0.87; 95%CI:0.77–0.99; I^2^:0.0%; 3 studies; 5 estimates).

### Associations between violence and ART use and adherence

Among 11 studies reporting an association between violence and ART use, one defined ART use as lifetime ART use among FSWs living with HIV, eight defined ART use as current ART use among FSWs living with HIV as the denominator, and two defined ART use as ART initiation among FSWs recently diagnosed with HIV. Recent exposure to any type of violence was associated with reduced odds of ART use (7 studies; 17 estimates; pOR: 0.78; 95%CI:0.64–0.94; I^2^:8%) (Figure 2). In subgroup analyses, UPSOV had the strongest association with reduced ART use (6 studies; 11 estimates; pOR: 0.68; 95%CI:0.54–0.87; I^2^:0%), with a weaker association between physical violence and ART use (3 studies; 3 estimates; pOR: 0.83; 95%CI:0.66–1.05; I^2^:0%), and only one estimate available for sexual violence. Lifetime exposure to any type of violence was not associated with ART use (3 studies; 4 estimates; pOR: 0.71; 95%CI:0.35–1.45; I^2^: 0%)(Figure 3).

Among five studies reporting an association between violence and ART adherence, timeframes for definitions of ART adherence ranged from four days to six months, with two studies reporting ART adherence in the time since HIV diagnosis. Recent experiences of any type of violence were not associated with ART adherence among FSWs (pOR: 0.65; 95%CI:0.33–1.29; I^2^:68%; 4 studies; 15 estimates) (Figure 2). However, in stratified analyses there were lower odds of ART adherence among FSWs who experienced recent UPSOV (pOR: 0.49; 95%CI:0.31–0.77; I^2^:69%; 4 studies; 11 estimates). No studies reported an association between lifetime experiences of violence and ART adherence.

### Associations between violence and HIV viral suppression

Among ten studies reporting an association with HIV viral suppression, seven measured viral suppression using all FSWs living with HIV as the denominator, and three used FSWs prescribed ART as the denominator. We found no association between recent exposure of any type of violence and HIV viral suppression (pOR: 0.88; 95%CI:0.67–1.17; I^2^:12%; 6 studies; 8 estimates) (Figure 2), but viral suppression was lower among FSWs who had lifetime exposure to any type of violence (pOR: 0.88; 95%CI:0.79–0.98; I^2^:20%; 5 studies; 6 estimates) (Figure 3).

### Subgroup analyses

We found little evidence that associations with prevalent HIV infection differed depending on the characteristics of the studies included in our analysis (Appendix 24). There was no evidence to suggest that the pooled associations differed between different WHO regions. We also did not detect differences in the pooled associations for different perpetrators of violence, whether studies were published before or after 2017, or for different recruitment strategies. However, there were higher pooled associations with prevalent HIV infection among studies that adjusted for confounding for both recent and lifetime exposure to violence. For lifetime exposure to violence, we found a stronger pooled effect among studies that did not use random sampling. We did not conduct subgroup analyses for the other HIV outcomes because there were too few studies.

### Publication asymmetry

For prevalent HIV infection, we observed asymmetry in the funnel plot of the associations with recent experiences of any type of violence which suggested disproportionately larger odds ratios in smaller studies, which was confirmed by the Egger’s test (P=0.001). When stratified by violence type, this asymmetry was most evident for recent sexual (P<0.001) or physical violence (P=0.015). There was no evidence of asymmetry for associations between lifetime experiences of violence and prevalent HIV infection (P=0.260). We did not detect asymmetry for estimates of the associations between lifetime violence and HIV testing (P=0.780), and there were fewer than 10 estimates for recent violence. We found evidence of asymmetry for associations between recent violence and ART use from funnel plots and Egger’s tests (P=0.013). There were fewer than 10 estimates for recent and lifetime violence and HIV incidence, ART adherence and HIV viral suppression, so we could not reliably assess for asymmetry.

### Sensitivity analyses

In sensitivity analyses using sampling correlations of p = 0.4 and p = 0.8, our findings remained robust, with pooled effect estimates and corresponding 95% confidence intervals changing marginally (Appendix 26).

## Discussion

In this systematic review and meta-analysis, we found evidence that both recent and lifetime exposure to violence are associated with higher odds of prevalent HIV infection among FSWs, with the strongest effect being seen among FSWs recently exposed to sexual violence. We also found reduced odds of ART use among FSWs who have recently experienced violence, and reduced viral suppression among FSWs who have experienced violence in their lifetime. However, we found no evidence of an association between violence and HIV testing uptake or HIV incidence, although few studies considered associations with outcomes other than prevalent HIV status, particularly incident HIV. The pooled proportion of FSWs experiencing violence in this study was high, with 20.2%, 15.0% and 21.9% of FSWs experiencing recent physical, sexual and UPSOV, respectively. These proportions align with estimates from a previous systematic review^10^, underscoring the high level of violence experienced by FSWs.

Our results align with evidence from other population groups. For instance, previous systematic reviews conducted among the general population have found strong and comparable associations between intimate partner violence and the same HIV outcomes^23^, including increased levels of HIV infection^24, 25^, reduced ART use^26^ and adherence^26^, and reduced HIV viral suppression.^26^ Similar associations have also been documented among other key populations in setting-specific studies, including people who inject drugs^27–29^, transgender women^30^ and men who have sex with men,^30, 31^ although these estimates have not been synthesised. This highlights the universal negative consequences of violence on HIV in varied population groups and so the crucial need for interventions to focus on this for improving health and wellbeing.

Our finding that all forms of violence are strongly associated with prevalent HIV infection among FSWs likely reflects multiple, interconnected pathways of increased marginalisation.^11^ Sexual violence, for example, can directly cause HIV acquisition through forced, unprotected sex. Physical and emotional violence, while not directly linked to HIV acquisition, may indirectly increase HIV risk by limiting women’s agency to negotiate safer sex, thereby making condomless sex more likely.^32, 33^ Violence is also associated with worse mental health^34^ and substance use^35, 36^, which in turn are associated with HIV risk behaviours such as reduced condom use and having more sexual partners.^10^ Reflecting these effects and potential mediating pathways, two mathematical modelling studies focusing on Kenya and Ukraine have shown that physical and sexual violence could be major drivers of HIV transmission, contributing 25-35% of new HIV infections among FSWs and their clients.^17, 33^

We also found evidence that violence is associated with worse HIV treatment outcomes, including ART use and HIV viral suppression. As with HIV transmission, multiple intersecting social and structural factors likely contribute to violence affecting HIV treatment outcomes. For some FSWs, disclosure of their HIV status or sex work can trigger violence from clients or intimate partners^37, 38^, potentially hindering or disrupting ART use through FSWs wanting to conceal use of HIV medications^39^ and experiencing psychological distress.^40^ Intimate partners who perpetrate violence may also limit the ability of their partners to attend HIV clinic appointments^39, 41^, or directly prevent them from taking ART.^39^ For FSWs living with HIV, experiences of violence can compound experiences of stigma and discrimination, eroding trust in healthcare providers and contributing to worse mental health, thus reducing their motivation to attend appointments or adhere consistently to ART.^42, 43^ Efforts to improve HIV treatment outcomes among FSWs living with HIV need to include interventions that reduce violence and provide safe working environments for FSWs.

A key challenge for designing and implementing interventions that reduce violence among FSWs is the limited evidence base.^44^ However, there are interventions that have been demonstrated to be effective in reducing violence. For instance, community empowerment and peer group membership has been shown to reduce experiences of violence among FSWs^45–49^, increase their capacity to exercise autonomy and self-protection^46, 48, 49^, and potentially reduce HIV incidence among FSWs.^50, 51^ Interventions focusing on social and structural contexts such as public health messaging, policy advocacy and education for law enforcement have also been shown to reduce experiences of violence among FSWs.^52–55^ In contrast, fewer interventions have focused on reducing behaviours that impact on the likelihood of experiencing violence, although two studies have found evidence that interventions to reduce substance use can be effective at reducing violence.^56, 57^ This evidence suggests that structural interventions should combine advocacy, community mobilisation and police education to reduce violence among FSWs. Continued investment in comprehensive, community-driven interventions is therefore required to address the complex factors contributing to violence among FSWs, which should then improve their HIV outcomes and promote their health and human rights.

Our study had limitations, many of which are reflective of the available evidence. First, for inclusion in this review, our search terms required a study’s title and/or abstract or keywords and MESH terms to mention violence (or related terms) and our specific HIV outcomes among FSWs. Consequently, we would not have identified any study where the association between violence and one of our HIV outcomes was only included in the main text of the study. Second, we found evidence of publication asymmetry for the associations between recent experiences of violence and prevalent HIV infection and ART use, suggesting the possibility of publication bias. This asymmetry may be partially explained by our title and abstract screening strategy, as studies which did not detect associations between violence and HIV outcomes are less likely to report them in their abstract. However, publication asymmetry may be explained by other factors, such as differences in study quality or design, heterogeneity in measurement methods, or statistical methods.^58^ Nonetheless, the results from the Egger’s tests suggest that our findings may be subject to publication bias. Third, there was heterogeneity in the measurement and definition of violence and some HIV outcomes across studies, which affected the comparability and synthesis of results. Whilst we conducted pooled analyses stratified by violence type, there were still variations in the definitions of violence within each strata. Fourth, most included studies were cross-sectional, making it challenging to establish temporality, and therefore causality. This is particularly relevant for associations between recent experiences of violence and prevalent HIV infection, as the timing of HIV infection is unknown, and may precede the exposure. Thus, we cannot rule out the possibility that some associations reflect increased likelihood of experiencing violence among FSWs who are living with HIV or at greater risk of HIV.

## Conclusion

Our findings underscore the substantial burden and impact of violence on various HIV outcomes among FSW. The large number of studies and strong pooled associations between violence and both prevalent HIV infection and HIV treatment outcomes highlight the importance of integrated HIV prevention and treatment approaches that address violence as a key structural determinant of health. While effective violence reduction interventions exist, experiences of violence remain common among FSWs, suggesting gaps in the implementation of these interventions. Reducing violence and improving HIV outcomes among FSWs will likely require a multi-pronged approach including sustained and increased investment in public health programmes for FSWs, and an increased focus on policy reform and sex-worker advocacy to enact broader structural changes. This will also likely result in broader improvements in the health and wellbeing of FSWs.

## Data Availability

All relevant data are within the manuscript and its Supporting Information files.

https://github.com/JoshuaDaweUoB/Structural-Barriers-FSW-Review

## Author contributions

**Joshua Dawe**: Conceptualisation, Methodology, Software, Formal Analysis, Investigation, Data Curation, Writing - Original Draft, Writing - Review and Editing, Visualisation, Project Administration, Funding Acquisition. **Khadimul Anam Mazhar**: Conceptualisation, Methodology, Funding Acquisition. **Saher Aijaz Khan**: Conceptualisation, Methodology, Funding Acquisition. **Belinda Jackson Njiro**: Investigation, Data Curation, Writing - Review and Editing. **Victoria Bendaud**: Conceptualisation, Project Administration, Funding Acquisition. **Keith Sabin**: Conceptualisation, Writing - Review and Editing, Project Administration, Funding Acquisition. **Julie Ambia**: Investigation. **Adam Trickey**: Investigation, Writing - Review and Editing. **Lucy Barrass**: Investigation, Writing - Review and Editing. **Asra Asgharzadeh**: Investigation. **Jack Stone**: Conceptualisation, Methodology, Investigation, Writing - Original Draft, Writing – Review, Supervision, Project Administration, Funding Acquisition. **Adelina Artenie**: Conceptualisation, Methodology, Writing - Original Draft, Writing – Review, Supervision, Project Administration, Funding Acquisition. **Peter Vickerman**: Conceptualisation, Methodology, Writing - Original Draft, Writing – Review, Supervision, Project Administration, Funding Acquisition.

## Data Sharing

The finalised data extraction sheet and analysis code is available at https://github.com/JoshuaDaweUoB/Structural-Barriers-FSW-Review.

