## Appendices for "Impact of violence on HIV outcomes among female sex workers: A global systematic review and meta-analysis"

**Table of Contents**

Appendix 1. Study Protocol	3

Appendix 2. PRISMA checklist	8

Appendix 3. Search Strategy	10

Appendix 4. Studies excluded because of duplicate data	12

Appendix 5. Studies identified through reference list searching	13

Appendix 6. Variables included in data extraction	14

Appendix 7. Study quality domains	15

Appendix 8. Definitions of forms of violence	16

Appendix 9. Definitions of HIV outcomes	16

Appendix 10. Variables extracted for subgroup analyses	17

Appendix 11. Characteristics of included studies (n=99)	18

Appendix 12. Estimates of the association between sexual violence and HIV outcomes	27

Appendix 13. Estimates of the association between physical violence and HIV outcomes	32

Appendix 14. Estimates of the association between physical and/or sexual violence and HIV outcomes	36

Appendix 15. Estimates of the association between other forms of violence and HIV outcomes	41

Appendix 16. Study quality scores of included studies	43

Appendix 17. Forest plots of all forms of violence estimates and prevalent HIV infection	46

Appendix 18. Forest plots of physical violence estimates and prevalent HIV infection	51

Appendix 19. Forest plots of sexual violence estimates and prevalent HIV infection	53

Appendix 20. Forest plots of mixed violence estimates and prevalent HIV infection	56

Appendix 21. Forest plots of other forms of violence estimates and prevalent HIV infection	58

Appendix 23. Forest plots of violence and HIV testing	59

Appendix 24. Forest plots of violence and ART outcomes	61

Appendix 25. Forest plots of violence and HIV viral suppression	64

Appendix 26. Subgroup analyses	66

Appendix 27. Publication asymmetry	68

Appendix 28. Sensitivity analyses of RHO	73

Reference list	74

### Appendix 1. Study Protocol

#### The impact of societal enablers and barriers on HIV outcomes among the key populations of people who inject drugs, men who have sex with men, transgender women and female sex workers: a protocol for a systematic review and meta-analysis

##### Citation

Joshua Dawe, Khadimul Mazhar, Saher Aijaz Khan, Adelina Artenie, Jack Stone, Matthew Hickman, Peter Vickerman. The impact of societal enablers and barriers on HIV outcomes among the key populations of people who inject drugs, men who have sex with men, transgender women and female sex workers: a protocol for a systematic review and meta-analysis. PROSPERO 2024 CRD42024531539 Available from: https://www.crd.york.ac.uk/prospero/display_record.php?ID=CRD42024531539

##### Review question

Our overall aim is to summarise the impact of societal enablers and barriers on HIV outcomes among four key populations: people who inject drugs (PWID), men who have sex with men (MSM), transgender women (TGW) and female sex workers (FSW). Our review aims to focus on four HIV outcomes: (1) HIV prevalence, (2) HIV incidence, (3) ART coverage and (4) viral suppression. Three societal enablers and barriers will be reviewed: (1) stigma, (2) violence, and (3) legal and policy environments. For each key population, we have assessed the quality of existing global systematic reviews or large-scale multi-country studies to identify evidence gaps for each societal enabler.

Specifically, we have identified the following gaps in the evidence base for each key population:

1. People who inject drugs (PWID): Stigma and violence
2. Men who have sex with men (MSM): Stigma and violence
3. Sex workers (SW): Legal and Policy environment and stigma
4. Transgender people (TG): Legal and Policy environment, stigma and violence

Our specific aims (SA) are to:

- SA1: synthesise evidence from research which estimates the association between societal enablers and barriers and HIV outcomes among each key population;
- SA2: Explore sources of heterogeneity in pooled associations from SA1.

##### Searches

Searches will be conducted in MEDLINE, Embase and PsycINFO.

The search string will be built using derivations and synonyms to describe each key population, societal enablers and barriers, and epidemiological studies. We will not apply language restrictions to the initial search. The search will be limited to studies published in 2010 and after.

##### Types of study to be included

Studies that could be used to estimate the association between societal enablers and barriers and HIV outcomes among each key population empirically will be included, including observational studies (case-control/cohort/cross-sectional) and intervention trials.

##### Inclusion criteria:

- Studies which report on the selected key populations (PWID/FSW/MSM/TG).
- Studies which report one of the societal enablers (stigma, violence, legal and policy environment);
- Studies which report data on HIV outcomes of interest.

##### Exclusion criteria:

- Case studies, systematic reviews, qualitative studies, animal studies, commentaries, editorials, and modelling studies.
- Studies with samples sizes under n=40.
- Studies where a key population is not the primary population of study (i.e. PWID/FSW/MSM/TG) will be excluded. Specifically, we will exclude studies where less than 70% of the sample includes a key population, or those which do not present data for a key population specifically.
- Studies which do not include a HIV outcome of interest

##### Condition or domain being studied

Studies that could be used to estimate the association between societal enablers and barriers and HIV outcomes among key populations at-risk of HIV.

##### Participants/population

We will focus on four key populations:

1. People who inject drugs.
2. Men who have sex with men.
3. Transgender people.
4. Female sex workers.

##### Intervention(s), exposure(s)

We will be exploring the impact of the following societal enablers and barriers on HIV outcomes:

- Stigma and discrimination
- Violence
- Legal and policy environment

##### Comparator(s)/control

Comparisons: these will be defined according to the strata listed in the exposure section.

##### Main outcome(s)

We will be studying four main HIV outcomes (HIV prevalence, HIV incidence, ART coverage and viral suppression).

##### Measures of effect

We are interested in measure of effects (i.e., odds ratio, prevalence ratio, risk ratio, hazard ratio) which estimate the association between societal enablers and barriers and the main HIV outcomes, for each population. See aims SA1–SA2.

##### Additional outcome(s)

None

##### Data extraction (selection and coding)

Records identified through database searches (MEDLINE, Embase and PsycINFO) will be assessed for duplicates. Title and abstract screening will be conducted in Rayyan AI software.

During title and abstract screening, three authors (JD, KM and SK) will double-screen titles and abstracts and all disagreements will be resolved by mutual agreement after discussion. For any unresolved disagreements, a third author (AA) will be included.

All three authors will double screen all the studies included for full-text screening. We will identify studies that include any of the four exposures of interest. The screening and selection processes will be documented, in line with PRISMA guidelines.

Study data will be double-extracted to ensure consistency. The data to be extracted will include:

1. Study characteristics (e.g., lead author, journal name, year of publication, start/end dates, design, city/country,
1. sampling strategy, recruitment site, inclusion/exclusion criteria, attrition (where appropriate));
2. Descriptive characteristics of study sample (e.g., age, proportion young, proportion female, proportion with exposure
3. of interest) and the proportion of study samples experiencing societal enablers and barriers;
4. Characteristics of the exposure (e.g., societal enablers and barriers definition, time frame of measurement) and measures of association between structural risk factors and the outcomes included in SA1–SA2.

##### Risk of bias (quality) assessment

Risk of bias assessment will be carried out independently by researchers and will be double screened/assessed to ensure consistency throughout, with disagreements resolved by discussion. We will use an adapted version of the Newcastle-Ottawa Scale (NOS) to assess risk of bias.

##### Strategy for data synthesis

A minimum of three estimates will be required to generate a pooled effect between societal enablers and barriers and HIV outcomes. We will provide descriptive summaries of effect estimates (magnitude and range of effects) where a pooled effect cannot be calculated. Cross-sectional and case-control studies will be pooled together, where appropriate we will pool cross-sectional and longitudinal effects together.

##### Descriptive data of each included study will be displayed in a summary table.

For each societal enabler and barrier, we will use random-effects meta-analyses to pool effect estimates comparing differences in outcomes between individuals exposed to a structural factor and individuals not exposed to a structural risk factor. Heterogeneity will be tested using the χ² test and quantified using the I²-statistic.

Where possible, we will perform sub-group and meta-regression analyses to investigate potential sources of heterogeneity. Publication bias will be explored using funnel plots and Egger's test.

##### Analysis of subgroups or subsets

We plan to report findings according to global geographic and economic regions.

##### Contact details for further information

Joshua Dawe

##### Organisational affiliation of the review

University of Bristol

https://www.bristol.ac.uk/medical-school/

##### Review team members and their organisational affiliations

Mr Joshua Dawe. Population Health Sciences, Bristol Medical School, University of Bristol

Mr Khadimul Mazhar. Population Health Sciences, Bristol Medical School, University of Bristol

Ms Saher Aijaz Khan. Population Health Sciences, Bristol Medical School, University of Bristol

Dr Adelina Artenie. Population Health Sciences, Bristol Medical School, University of Bristol

Dr Jack Stone. Population Health Sciences, Bristol Medical School, University of Bristol

Professor Matthew Hickman. Population Health Sciences, Bristol Medical School, University of Bristol

Professor Peter Vickerman. Population Health Sciences, Bristol Medical School, University of Bristol

##### Type and method of review

Epidemiologic, Meta-analysis, Systematic review

##### Anticipated or actual start date

26 March 2024

##### Anticipated completion date

31 May 2024

##### Funding sources/sponsors

The Joint United Nations Programme on HIV/AIDS (UNAIDS)

##### Conflicts of interest

Language

English

##### Country

England

##### Stage of review

Review Ongoing

##### Subject index terms status

Subject indexing assigned by CRD

##### Subject index terms

Drug Users; Evidence Gaps; Female; HIV Infections; Homosexuality, Male; Humans; Incidence; Male; Meta-Analysis as Topic; Policy; Prevalence; Review Literature as Topic; Sex Workers; Sexual and Gender Minorities; Substance Abuse, Intravenous; Systematic Reviews as Topic; Transgender Persons; Violence

##### Date of registration in PROSPERO

04 April 2024

##### Date of first submission

02 April 2024

##### Stage of review at time of this submission

| Stage | Started | Completed |
| --- | --- | --- |
| Preliminary searches | Yes | Yes |
| Piloting of the study selection process | Yes | Yes |
| Formal screening of search results against eligibility criteria | Yes | No |
| Data extraction | No | No |
| Risk of bias (quality) assessment | No | No |
| Data analysis | No | No |

### Appendix 2. PRISMA checklist

| **Section and Topic** | **Item #** | **Checklist item** | **Location where item is reported** |
| --- | --- | --- | --- |
| **TITLE** |  |  |  |
| Title | 1 | Identify the report as a systematic review. | 1 |
| **ABSTRACT** |  |  |  |
| Abstract | 2 | See the PRISMA 2020 for Abstracts checklist. | 2 |
| **INTRODUCTION** |  |  |  |
| Rationale | 3 | Describe the rationale for the review in the context of existing knowledge. | 3 |
| Objectives | 4 | Provide an explicit statement of the objective(s) or question(s) the review addresses. | 6 |
| **METHODS** |  |  |  |
| Eligibility criteria | 5 | Specify the inclusion and exclusion criteria for the review and how studies were grouped for the syntheses. | 7-9 |
| Information sources | 6 | Specify all databases, registers, websites, organisations, reference lists and other sources searched or consulted to identify studies. Specify the date when each source was last searched or consulted. | 7-8 |
| Search strategy | 7 | Present the full search strategies for all databases, registers and websites, including any filters and limits used. | S10-S11 |
| Selection process | 8 | Specify the methods used to decide whether a study met the inclusion criteria of the review, including how many reviewers screened each record and each report retrieved, whether they worked independently, and if applicable, details of automation tools used in the process. | 7-8 |
| Data collection process | 9 | Specify the methods used to collect data from reports, including how many reviewers collected data from each report, whether they worked independently, any processes for obtaining or confirming data from study investigators, and if applicable, details of automation tools used in the process. | 8 |
| Data items | 10a | List and define all outcomes for which data were sought. Specify whether all results that were compatible with each outcome domain in each study were sought (e.g. for all measures, time points, analyses), and if not, the methods used to decide which results to collect. | 7-9 |
|  | 10b | List and define all other variables for which data were sought (e.g. participant and intervention characteristics, funding sources). Describe any assumptions made about any missing or unclear information. | 7-9 |
| Study risk of bias assessment | 11 | Specify the methods used to assess risk of bias in the included studies, including details of the tool(s) used, how many reviewers assessed each study and whether they worked independently, and if applicable, details of automation tools used in the process. | 9 |
| Effect measures | 12 | Specify for each outcome the effect measure(s) (e.g. risk ratio, mean difference) used in the synthesis or presentation of results. | 8-9 |
| Synthesis methods | 13a | Describe the processes used to decide which studies were eligible for each synthesis (e.g. tabulating the study intervention characteristics and comparing against the planned groups for each synthesis (item #5)). | 8-9 |
|  | 13b | Describe any methods required to prepare the data for presentation or synthesis, such as handling of missing summary statistics, or data conversions. | 8-9 |
|  | 13c | Describe any methods used to tabulate or visually display results of individual studies and syntheses. | 8-9 |
|  | 13d | Describe any methods used to synthesize results and provide a rationale for the choice(s). If meta-analysis was performed, describe the model(s), method(s) to identify the presence and extent of statistical heterogeneity, and software package(s) used. | 8-9 |
|  | 13e | Describe any methods used to explore possible causes of heterogeneity among study results (e.g. subgroup analysis, meta-regression). | 9 |
|  | 13f | Describe any sensitivity analyses conducted to assess robustness of the synthesized results. | 8-9 |
| Reporting bias assessment | 14 | Describe any methods used to assess risk of bias due to missing results in a synthesis (arising from reporting biases). | 9 |
| Certainty assessment | 15 | Describe any methods used to assess certainty (or confidence) in the body of evidence for an outcome. | 8-9 |
| **RESULTS** |  |  |  |
| Study selection | 16a | Describe the results of the search and selection process, from the number of records identified in the search to the number of studies included in the review, ideally using a flow diagram. | 10-11 |
|  | 16b | Cite studies that might appear to meet the inclusion criteria, but which were excluded, and explain why they were excluded. | 11 |
| Study characteristics | 17 | Cite each included study and present its characteristics. | Sxx-Sxx |
| Risk of bias in studies | 18 | Present assessments of risk of bias for each included study. | Sxx-Sxx |
| Results of individual studies | 19 | For all outcomes, present, for each study: (a) summary statistics for each group (where appropriate) and (b) an effect estimate and its precision (e.g. confidence/credible interval), ideally using structured tables or plots. | Sxx-Sxx |
| Results of syntheses | 20a | For each synthesis, briefly summarise the characteristics and risk of bias among contributing studies. | Xx |
|  | 20b | Present results of all statistical syntheses conducted. If meta-analysis was done, present for each the summary estimate and its precision (e.g. confidence/credible interval) and measures of statistical heterogeneity. If comparing groups, describe the direction of the effect. | 12-14; Sxx-Sxx |
|  | 20c | Present results of all investigations of possible causes of heterogeneity among study results. | 15; Sxx-Sxx |
|  | 20d | Present results of all sensitivity analyses conducted to assess the robustness of the synthesized results. | 15; Sxx-Sxx |
| Reporting biases | 21 | Present assessments of risk of bias due to missing results (arising from reporting biases) for each synthesis assessed. | 15; Sxx-Sxx |
| Certainty of evidence | 22 | Present assessments of certainty (or confidence) in the body of evidence for each outcome assessed. | 12-15 |
| **DISCUSSION** |  |  |  |
| Discussion | 23a | Provide a general interpretation of the results in the context of other evidence. | 18-20 |
|  | 23b | Discuss any limitations of the evidence included in the review. | 20 |
|  | 23c | Discuss any limitations of the review processes used. | 20 |
|  | 23d | Discuss implications of the results for practice, policy, and future research. | 18-19 |
| **OTHER INFORMATION** | |  |  |
| Registration and protocol | 24a | Provide registration information for the review, including register name and registration number, or state that the review was not registered. | 7 |
|  | 24b | Indicate where the review protocol can be accessed, or state that a protocol was not prepared. | 7 |
|  | 24c | Describe and explain any amendments to information provided at registration or in the protocol. | NA |
| Support | 25 | Describe sources of financial or non-financial support for the review, and the role of the funders or sponsors in the review. | 3 |
| Competing interests | 26 | Declare any competing interests of review authors. | 3 |
| Availability of data, code, and other materials | 27 | Report which of the following are publicly available and where they can be found: template data collection forms; data extracted from included studies; data used for all analyses; analytic code; any other materials used in the review. | 20 |

### Appendix 3. Search Strategy

Database search strategy to identify relevant studies

1. exp social stigma/
2. exp Stigma/
3. exp drug discrimination/
4. exp perceived discrimination/
5. exp prejudice/
6. exp Stereotyping/
7. exp social discrimination/
8. exp perceptive discrimination/
9. (stigma or discrimin* or "social stigma" or (social adj3 stigma) or stigm* or prejudice or stigma or discrimination or bias or intolerance or harass*).mp.
10. 1 or 2 or 3 or 4 or 5 or 6 or 7 or 8 or 9
11. exp obstetric violence/
12. exp workplace violence/
13. exp family violence/
14. exp partner violence/
15. exp sexual violence/
16. exp exposure to violence/
17. exp police violence/
18. exp physical violence/
19. exp domestic violence/
20. exp Gender-Based Violence/
21. exp gender based violence/
22. exp dating violence/
23. exp violence/
24. exp gun violence/
25. exp Ethnic Violence/
26. exp Intimate Partner Violence/
27. violence.mp.
28. 11 or 12 or 13 or 14 or 15 or 16 or 17 or 18 or 19 or 20 or 21 or 22 or 23 or 24 or 25 or 26 or 27
29. exp criminal law/
30. exp policy/
31. exp government regulation/
32. exp legal services/
33. exp liability, legal/
34. exp law enforcement/
35. exp legislation, drug/
36. exp social control, informal/
37. exp "health care quality, access, and evaluation"/
38. exp Police/
39. exp Law enforcement/
40. exp Legislation/
41. exp Crime/
42. exp Prisons/
43. (criminalis* or criminaliz* or prohibit* or decriminalis* or decriminaliz* or legalisation or legalization or legislat* or law* or legal* or illegal* or policing or police* or enforce* or criminal justice or prison* or penalt* or fine* or raid* or police operation* or arrest* or detention* or detain* or sanction* or dispersal order* or zoning restriction* or zon* or convict* or police record* or criminal record*).mp.
44. (anti-social behaviour* or (police adj3 harrass*) or (police adj3 repress*) or (police adj3 warning) or (police adj3 evict*) or (police adj3 engage$) or (police adj3 caution) or civil order* or administrative offense or (police adj3 coercion) or confiscat* or crime prevention or brothel closure* or confiscat* or loiter* or solicit* or support order or rehabilit* or (vagrancy adj3 act) or (municipal adj3 order) or by-law or police repress* or licens* or regulation or mandatory test* or registration or forced test* or condoms as evidence).mp.
45. 29 or 30 or 31 or 32 or 33 or 34 or 35 or 36 or 37 or 38 or 39 or 40 or 41 or 42 or 43 or 44
46. exp HIV Seroprevalence/
47. exp HIV-2/
48. exp HIV Infections/
49. exp HIV-1/
50. exp HIV Seropositivity/
51. exp HIV Testing/
52. exp HIV/
53. exp HIV Antibodies/
54. (HIV or "acquired immunodeficiency syndrome" or "AIDS" or "aids" or "hiv").mp.
55. 46 or 47 or 48 or 49 or 50 or 51 or 52 or 53 or 54
56. exp Sex Work/
57. ("sex work*" or prostitut* or "street walker*" or escort* or rent boy* or "sell sex" or "sold sex" or "selling sex" or "exchanged sex" or "exchange sex" or "exchanged sex" or "sex trade" or "commercial sex" or "sex industry" or "transactional sex" or "sexual favour" or "bar hostess" or "red light district" or "escort*").mp.
58. 56 or 57
59. exp Data Collection/ or Longitudinal Studies/ or longitudinal stud*.mp. or regression analysis/ or linear models/ or logistic models/ or proportional hazards models/ or spatial regression/ or survival analysis/ or epidemiologic studies/ or case-control studies/ or case control stud*.mp. or retrospective studies/ or cohort studies/ or follow-up studies/ or longitudinal studies/ or prospective studies/ or controlled before-after studies/ or cross-sectional studies/ or cross?sectional stud*.mp. or historically controlled study/ or interrupted time series analysis/ or seroepidemiologic studies/ or seroprevalence stud*.mp. or hiv seroprevalence/ or feasibility studies/ or multicenter studies as topic/ or pilot projects/ or sampling studies/ or sentinel surveillance/ or seroprevalence stud*.mp. or bio behavio?ral stud*.mp. or bio?behavio?ral stud*.mp. or health survey.mp. or exp Health Surveys/ or sampling studies.mp. or exp Sampling Studies/
60. 10 or 28 or 45
61. 55 and 58
62. 60 and 61
63. 59 and 62
64. limit 63 to yr="2010 -Current"
65. remove duplicates from 64

### Appendix 4. Studies excluded because of duplicate data

| **Lead author (year)** | **Title** | **Journal** |
| --- | --- | --- |
| Braunstein (2012) | Risk factor detection as a metric of STARHS performance for HIV incidence surveillance among female sex workers in Kigali, Rwanda | The open AIDS journal |
| Kerrigan (2019) | Project Shikamana: Community empowerment-based Combination HIV Prevention Significantly Impacts HIV Incidence and Care Continuum Outcomes among Female Sex Workers in Iringa, Tanzania | JAIDS Journal of Acquired Immune Deficiency Syndromes |
| Pickles (2024) | Exploratory analysis of the potential impact of violence on HIV among female sex workers in Mombasa, Kenya: a mathematical modelling study | BMC Medicine |
| Tounkara (2013) | Relationship between violence and HIV infection among female sex workers in Benin | Sexually Transmitted Infections |
| Mountain (2018) | Experience of violence is associated with HIV prevention uptake and HIV infection among young women who self-identify as sex workers in Mombasa, Kenya | HIV Research for Prevention 2018 |

### Appendix 5. Studies identified through reference list searching

| **Lead author (year)** | **Title** | **Journal** |
| --- | --- | --- |
| Amogne (2019) | Prevalence and correlates of physical violence and rape among female sex workers in Ethiopia: a Cross-sectional study with Respondent-driven sampling from 11 major towns | BMJ Open |
| Comins (2024) | ART coverage and viral suppression among female sex workers living with HIV in eThekwini, South Africa: Baseline findings from the Siyaphambili study | PLOS Global Public Health |
| Lima (2017) | Factors associated with violence against female sex workers in ten Brazilian cities | Cadernos de Saúde Pública |
| Mountain (2018) | Associations between violence, HIV prevention and HIV infection among young women who self-identify as sex workers in Mombasa, Kenya | PhD Dissertation |
| Prangnell (2018) | Workplace violence among female sex workers who use drugs in Vancouver, Canada: does client-targeted policing increase safety? | Journal of public health policy |
| Vu (2017) | High Burden of HIV, Syphilis and HSVNA2 and Factors Associated with HIV Infection Among Female Sex Workers in Tanzania: Implications for Early Treatment of HIV and Pre-exposure Prophylaxis (PrEP) | AIDS and Behavior |
| West (2022) | Typologies and Correlates of Police Violence Against Female Sex Workers Who Inject Drugs at the Mexico-United States Border: Limits of De Jure Decriminalization in Advancing Health and Human Rights | Journal of interpersonal violence |
| Wilson (2016b) | Prevalence and correlates of intimate partner violence in HIV-positive women engaged in transactional sex in Mombasa, Kenya | International journal of STD & AIDS |
| Wirth (2013) | How Does Sex Trafficking Increase the Risk of HIV Infection? An Observational Study From Southern India | American Journal of Epidemiology |

### Appendix 6. Variables included in data extraction

Data was extracted on:

- Lead author, review title or unique identifier and date
- Publication type
- Study design
- Recruitment strategy
- Study location
- Study setting
- Study period (years in which recruitment occurred)
- Study sample size
- Study inclusion and exclusion criteria (including definition of PWID, e.g. injecting in the last 30 days)
- Definition of exposure
- Time frame of exposure
- Perpetrator of exposure
- Number and proportion of exposed participants
- Definition of outcome
- Unadjusted and adjusted effect size (e.g. incidence rate ratio (IRR); odds ratio (OR); hazard ratio (HR); and precision (e.g. 95% confidence interval (CI))
- Covariates included in adjusted models

### Appendix 7. Study quality domains

| **Domain** | **Description** |
| --- | --- |
| Domain 1: Sample representativeness of the target population | This domain assessed the extent to which the study sample was likely to approximate the target population. One point was assigned if the study used recruitment approaches intended to enhance representativeness among KPs, including RDS, time–location or venue-based sampling, or recruitment through multiple, diverse sources (e.g., NSPs, shelters, street outreach and OAT clinics for the PWID review). Probability-based sampling, while rarely used, was also considered indicative of high representativeness. To receive one point, studies could not be restricted to narrowly defined subgroups (e.g., specific age groups, gender identities, or sexual orientations), unless such restrictions were intrinsic to the study population (e.g., all women in studies of FSW). Studies that did not meet these criteria were assigned zero points. |
| Domain 2: Temporality of exposure and outcome | This domain assessed whether stigma or violence exposure clearly preceded the HIV-related outcome of interest. One point was assigned to longitudinal studies in which exposure was measured prior to outcome assessment. Longitudinal studies that assessed exposure and outcome over the same time period, as well as cross-sectional studies, were assigned zero points. |
| Domain Three: Control for confounding | This domain assessed whether estimates were adjusted for potential confounders. One point was assigned if estimates were adjusted; unadjusted estimates were assigned zero points. |

### Appendix 8. Definitions of forms of violence

| Societal barrier | Description |
| --- | --- |
| Physical violence | Acts of physical violence perpetrated against study participants, and experiences in which participants reported feeling physically threatened or endangered. |
| Sexual violence | Any sexual act, attempt to obtain a sexual act, or other act directed against a study participant sexuality using coercion, by any person regardless of their relationship, in any setting. It includes rape, defined as the physically forced or otherwise coerced penetration of the vulva or anus with a penis, other body part or object. |
| Physical and/or sexual violence | Acts of violence perpetrated against study participants, where the nature of the violence is not specified, or sexual violence and physical violence have been combined. |
| Other violence | Other forms of violence that do not constitute physical violence or sexual violence (for example, emotional violence). |

### Appendix 9. Definitions of HIV outcomes

| HIV Outcome | Numerator | Denominator | Description |
| --- | --- | --- | --- |
| HIV infection | FSWs living with HIV | All FSWs | Measures the proportion of all FSWs in the community who are living with HIV. |
| HIV incidence | Incident HIV infections | Person-time at-risk of HIV or recency assay | Longitudinally measured using tests subsequent to an initial negative HIV test. Recency assays use biomarkers to detect new and longstanding HIV infections. |
| ART initiation | FSWs prescribed ART | FSWs newly diagnosed with HIV | FSWs who were recently diagnosed with HIV who received a prescription for ART and initiated treatment |
| ART use | FSWs living with HIV on ART | FSWs living with diagnosed HIV | FSWs who are living with HIV and are currently prescribed ART. |
| ART adherence | FSWs adhering to ART regimen | FSWs living with diagnosed HIV with ART prescription | The proportion of ART doses taken on time within a given time period. |
| HIV viral suppression | Virally suppressed FSWs living with HIV | FSWs living with diagnosed HIV or FSWs living with diagnosed HIV with ART prescription | A measure of HIV viral load, including both dichotomous indicators of viral suppression (viral load above or below a particular threshold), and continuous measures of viral load. |

### Appendix 10. Variables extracted for subgroup analyses

| Variable | Description |
| --- | --- |
| WHO region | The WHO regions are divided into six geographic areas: Africa, Americas, South-East Asia, Europe, Eastern Mediterranean, and Western Pacific. |
| Country income levels | Dichotomised into two categories (lower-income and lower-middle income, upper-middle income and upper income) |
| Year of publication | Dichotomised into two categories (published between 2010–2016 and published between 2017–2025). |
| Perpetrator | Perpetrator of violence. Includes clients, employers, intimate partners, family or friends, police, and healthcare workers. |
| Recruitment method | Strategy used to identify, contact, and enrol participants. Includes respondent-driven sampling, time-space sampling, convenience sampling, and probabilistic sampling. |
| Study quality | Study quality was assessed on three criteria: representativeness, study design and confounder adjustment |

### Appendix 11. Characteristics of included studies (n=99)

Geographic and design characteristics of included studies

| **Author (year)** | **Publication type** | **Study design** | **Recruitment method** | **Country** | **WHO region** |
| --- | --- | --- | --- | --- | --- |
| Abdella (2022) (1) | Ethiopia | Manuscript | Cross-sectional | Respondent-driven | African Region |
| Aho (2013) (2) | Guinea | Manuscript | Cross-sectional | Convenience | African Region |
| Alaei (2021) (3) | Tajikistan | Manuscript | Cross-sectional | Respondent-driven | European Region |
| Alary (2014) (4) | India | Manuscript | Cross-sectional | Respondent-driven, cluster and time-space sampling | South-East Asia Region |
| Alemu (2022) (5) | Ethiopia | Manuscript | Cross-sectional | Probabilistic | African Region |
| Amogne (2019) (6) | Ethiopia | Manuscript | Cross-sectional | Respondent-driven | African Region |
| Arimide (2022) (7) | Ethiopia | Manuscript | Cross-sectional | Respondent-driven | African Region |
| Arumugam (2022) (8) | India | Manuscript | Cross-sectional | Probabilistic | South-East Asia Region |
| Ayamah (2023) (9) | Ghana | Manuscript | Cross-sectional | Time-space sampling | African Region |
| Beattie (2010) (10) | India | Manuscript | Cross-sectional | Time-space sampling | South-East Asia Region |
| Beattie (2015) (11) | India | Manuscript | Cross-sectional | Time-space sampling | South-East Asia Region |
| Beksinska (2018) (12) | India | Manuscript | Cross-sectional | Time-space sampling | South-East Asia Region |
| Berger (2018) (13) | Eswatini | Manuscript | Cross-sectional | Respondent-driven | African Region |
| Bhardwaj (2023) (14) | South Africa | Manuscript | Cross-sectional | Convenience | African Region |
| Bowring (2019) (15) | Cameroon | Manuscript | Cross-sectional | Respondent-driven | African Region |
| Braunstein (2011) (16) | Rwanda | Manuscript | Cross-sectional | Convenience | African Region |
| Budhwani (2017) (17) | Jamaica | Manuscript | Cross-sectional | Convenience | Region of the Americas |
| Budhwani (2021) (18) | Dominican Republic | Manuscript | Cross-sectional | Convenience | Region of the Americas |
| Bugssa (2015) (19) | Ethiopia | Manuscript | Cross-sectional | Probabilistic | African Region |
| Chabata (2019) (20) | Zimbabwe | Manuscript | Cross-sectional | Respondent-driven | African Region |
| Coetzee (2017) (21) | South Africa | Manuscript | Cross-sectional | Respondent-driven | African Region |
| Comins (2024) (22) | South Africa | Manuscript | Cross-sectional | Convenience | African Region |
| Damacena (2011) (23) | Brazil | Manuscript | Cross-sectional | Respondent-driven | Region of the Americas |
| Davey (2020) (24) | Zimbabwe | Manuscript | Cross-sectional | Respondent-driven | African Region |
| Decker (2012) (25) | Russia | Manuscript | Cross-sectional | Respondent-driven | European Region |
| Decker (2014) (26) | Russia | Manuscript | Cross-sectional | Respondent-driven | European Region |
| Decker (2016) (27) | Cameroon | Manuscript | Cross-sectional | Respondent-driven | African Region |
| Doshi (2018) (28) | Uganda | Manuscript | Cross-sectional | Respondent-driven | African Region |
| Duff (2016) (29) | Canada | Manuscript | Prospective cohort | Time-space sampling | Region of the Americas |
| Emmanuel (2021) (30) | Pakistan | Manuscript | Cross-sectional | Probabilistic | Eastern Mediterranean Region |
| Faini (2022) (31) | Tanzania | Manuscript | Cross-sectional | Respondent-driven | African Region |
| Goldenberg (2013) (32) | Mexico | Manuscript | Cross-sectional | Respondent-driven | Region of the Americas |
| Goldenberg (2016a) (33) | Uganda | Manuscript | Cross-sectional | Time-space sampling | African Region |
| Goldenberg (2016b) (34) | Canada | Manuscript | Prospective cohort | Time-space sampling | Region of the Americas |
| Goldenberg (2019) (35) | Uganda | Manuscript | Cross-sectional | Time-space sampling | African Region |
| Guure (2023) (36) | Ghana | Manuscript | Cross-sectional | Time-space sampling | African Region |
| Hendrickson (2018) (37) | Tanzania | Manuscript | Cross-sectional | Time-space sampling | African Region |
| Hensen (2019) (38) | Zimbabwe | Manuscript | Cross-sectional | Respondent-driven | African Region |
| Hladik (2017) (39) | Uganda | Manuscript | Cross-sectional | Respondent-driven | African Region |
| Iakunchykova (2017) (40) | Ukraine | Manuscript | Cross-sectional | Time-space sampling | European Region |
| Izadi (2023) (41) | Iran | Manuscript | Cross-sectional | Respondent-driven | Eastern Mediterranean Region |
| Jennings Mayo-Wilson (2023) (42) | Uganda | Manuscript | Cross-sectional | Time-space sampling | African Region |
| Jones (2023) (43) | Zimbabwe | Manuscript | Retrospective cohort | Convenience | African Region |
| Kakisingi (2020) (44) | Democratic Republic of the Congo | Manuscript | Cross-sectional | Convenience | African Region |
| Kassanjee (2022) (45) | South Africa | Manuscript | Cross-sectional | Convenience | African Region |
| Kelly-Hanku (2020) (46) | Papua New Guinea | Manuscript | Cross-sectional | Respondent-driven | Western Pacific Region |
| Kerrigan (2017) (47) | Tanzania | Manuscript | Cross-sectional | Time-space sampling | African Region |
| Kumar (2014) (48) | India | Manuscript | Cross-sectional | Respondent-driven | South-East Asia Region |
| Leddy (2018) (49) | Tanzania | Manuscript | Cross-sectional | Time-space sampling | African Region |
| Lima (2017) (50) | Brazil | Manuscript | Cross-sectional | Respondent-driven | Region of the Americas |
| Lyons (2017) (51) | Côte d’Ivoire | Manuscript | Cross-sectional | Respondent-driven | African Region |
| Lyons (2020) (52) | Multi-country | Manuscript | Cross-sectional | Respondent-driven | African Region |
| MacLin (2023) (53) | Dominican Republic | Manuscript | Cross-sectional | Convenience | Region of the Americas |
| Mendoza (2017) (54) | Dominican Republic | Manuscript | Cross-sectional | Snowball and peer-led | Region of the Americas |
| Mizinduko (2020) (55) | Tanzania | Manuscript | Cross-sectional | Respondent-driven | African Region |
| Mountain (2018) (56) | Kenya | PhD dissertation | Cross-sectional | Time-space sampling and cluster sampling | African Region |
| Mukherjee (2022) (57) | Kazakhstan | Manuscript | Cross-sectional | Convenience | European Region |
| Mukuku (2024) (58) | Democratic Republic of the Congo | Manuscript | Cross-sectional | Respondent-driven | African Region |
| Mulholland (2022) (59) | multi-country | Manuscript | Cross-sectional | Probabilistic | African Region |
| Mutagoma (2017) (60) | Rwanda | Manuscript | Cross-sectional | Time-space sampling | African Region |
| Mutagoma (2019) (61) | Rwanda | Manuscript | Cross-sectional | Time-space sampling | African Region |
| Nabayinda (2023) (62) | Uganda | Manuscript | Cross-sectional | Time-space sampling | African Region |
| Namale (2019) (63) | Uganda | Manuscript | Cross-sectional | Convenience | African Region |
| Oldenburg (2018) (64) | Zambia | Manuscript | Cross-sectional | Respondent-driven | African Region |
| Ouma (2021) (65) | Uganda | Manuscript | Cross-sectional | Convenience | African Region |
| Pando (2013) (66) | Argentina | Manuscript | Cross-sectional | Convenience | Region of the Americas |
| Prangnell (2018) (67) | Canada | Manuscript | Cross-sectional | Convenience | Region of the Americas |
| Ramesh (2012) (68) | India | Manuscript | Cross-sectional | Time-space sampling and cluster sampling | South-East Asia Region |
| Rice (2022) (69) | Zimbabwe | Manuscript | Cross-sectional | Respondent-driven | African Region |
| Schwartz (2016) (70) | South Africa | Manuscript | Cross-sectional | Respondent-driven | African Region |
| Sherman (2019) (71) | USA | Manuscript | Cross-sectional | Time-space sampling | Region of the Americas |
| Shokoohi (2017) (72) | Iran | Manuscript | Cross-sectional | Convenience | Eastern Mediterranean Region |
| Shrestha (2017) (73) | Nepal | Manuscript | Cross-sectional | Probabilistic | South-East Asia Region |
| Sileo (2018) (74) | Uganda | Manuscript | Cross-sectional | Respondent-driven | African Region |
| Strathdee (2011) (75) | Mexico | Manuscript | Cross-sectional | Respondent-driven | Region of the Americas |
| Szwarcwald (2017) (76) | Brazil | Manuscript | Cross-sectional | Respondent-driven | Region of the Americas |
| Tokar (2013) (77) | Ukraine | Conference abstract | Cross-sectional | Respondent-driven and time-space sampling | European Region |
| Tounkara (2014) (78) | Benin | Manuscript | Cross-sectional | Probabilistic | African Region |
| Tuot (2020) (79) | Cambodia | Manuscript | Cross-sectional | Respondent-driven | South-East Asia Region |
| Ulibarri (2011) (80) | Mexico | Manuscript | Cross-sectional | Respondent-driven | Region of the Americas |
| Vandenhoudt (2013) (81) | Kenya | Manuscript | Cross-sectional | Respondent-driven | African Region |
| Vu (2017) (82) | Tanzania | Manuscript | Cross-sectional | Respondent-driven | African Region |
| Wand (2015) (83) | Papua New Guinea | Manuscript | Cross-sectional | Respondent-driven | South-East Asia Region |
| Wang (2021) (84) | Malaysia | Manuscript | Cross-sectional | Respondent-driven | South-East Asia Region |
| West (2022) (85) | Mexico | Manuscript | Cross-sectional | Convenience | Region of the Americas |
| Wilson (2016a) (86) | Kenya | Manuscript | Prospective cohort | Convenience | African Region |
| Wilson (2016b) (87) | Kenya | Manuscript | Cross-sectional | Convenience | African Region |
| Wirth (2013) (88) | India | Manuscript | Cross-sectional | Convenience | South-East Asia Region |
| Xie (2023) (89) | China | Manuscript | Cross-sectional | Convenience | South-East Asia Region |
| Zalla (2019) (90) | Haiti | Manuscript | Cross-sectional | Probabilistic | Region of the Americas |
| Zhang (2012) (91) | China | Manuscript | Cross-sectional | Convenience | South-East Asia Region |

Sex work and HIV characteristics of included studies

| **Author (year)** | **Country** | **Overall sample size** | **Analytic sample size** | **Number HIV+** | **Percent HIV+** |
| --- | --- | --- | --- | --- | --- |
| Abdella (2022) (1) | Ethiopia | 6085 | 6085 | 1138 | 19% |
| Aho (2013) (2) | Guinea | 223 | 221 | 78 | 35% |
| Alaei (2021) (3) | Tajikistan | 2174 | 2174 | 64 | 3% |
| Alary (2014) (4) | India | 23176 | 21707 | 3385 | 16% |
| Alemu (2022) (5) | Ethiopia | 381 | 381 | 76 | 20% |
| Amogne (2019) (6) | Ethiopia | 4900 | 4900 | 1173 | 24% |
| Arimide (2022) (7) | Ethiopia | 4900 | 239 | 239 | 100% |
| Arumugam (2022) (8) | India | 27007 | 27007 | 548 | 2% |
| Ayamah (2023) (9) | Ghana | 4279 | 3493 | 222 | 6% |
| Beattie (2010) (10) | India | 3852 | 3852 | 634 | 16% |
| Beattie (2015) (11) | India | 5792 | 3909 | 495 | 13% |
| Beksinska (2018) (12) | India | 1111 | 1111 | 91 | 8% |
| Berger (2018) (13) | Eswatini | 325 | 325 | 222 | 70% |
| Bhardwaj (2023) (14) | South Africa | 1384 | 1384 | 1384 | 100% |
| Bowring (2019) (15) | Cameroon | 2255 | 2255 | 550 | 24% |
| Braunstein (2011) (16) | Rwanda | 800 | 629 | 192 | 24% |
| Budhwani (2017) (17) | Jamaica | 459 | 459 | 6 | 1% |
| Budhwani (2021) (18) | Dominican Republic | 307 | 307 | 110 | 36% |
| Bugssa (2015) (19) | Ethiopia | 319 | 319 | 38 | 12% |
| Chabata (2019) (20) | Zimbabwe | 503 | 502 | 173 | 34% |
| Coetzee (2017) (21) | South Africa | 508 | 508 | 280 | 55% |
| Comins (2024) (22) | South Africa | 1373 | 1373 | 1373 | 100% |
| Damacena (2011) (23) | Brazil | 2523 | 2523 | 121 | 5% |
| Davey (2020) (24) | Zimbabwe | 2839 | 2839 | 1671 | 59% |
| Decker (2012) (25) | Russia | 147 | 143 | 7 | 5% |
| Decker (2014) (26) | Russia | 754 | 754 | 29 | 4% |
| Decker (2016) (27) | Cameroon | 1817 | 81 | 81 | 100% |
| Doshi (2018) (28) | Uganda | 1497 | 1497 | 485 | 31% |
| Duff (2016) (29) | Canada | 72 | 72 | 72 | 100% |
| Emmanuel (2021) (30) | Pakistan | 5660 | 5660 | 121 | 2% |
| Faini (2022) (31) | Tanzania | 773 | 773 | 59 | 8% |
| Goldenberg (2013) (32) | Mexico | 214 | 214 | 9 | 4% |
| Goldenberg (2016a) (33) | Uganda | 400 | 400 | 135 | 34% |
| Goldenberg (2016b) (34) | Canada | 74 | 74 | 74 | 100% |
| Goldenberg (2019) (35) | Uganda | 400 | 89 | 89 | 100% |
| Guure (2023) (36) | Ghana | 5990 | 5679 | 264 | 5% |
| Hendrickson (2018) (37) | Tanzania | 496 | 496 | 203 | 41% |
| Hensen (2019) (38) | Zimbabwe | 2387 | 2387 | 543 | 24% |
| Hladik (2017) (39) | Uganda | 942 | 942 | 323 | 33% |
| Iakunchykova (2017) (40) | Ukraine | 4806 | 4806 | 270 | 6% |
| Izadi (2023) (41) | Iran | 1515 | 1515 | 24 | 2% |
| Jennings Mayo-Wilson (2023) (42) | Uganda | 542 | 542 | 192 | 35% |
| Jones (2023) (43) | Zimbabwe | 6665 | 6665 | NR | NR |
| Kakisingi (2020) (44) | Democratic Republic of the Congo | 1555 | 1555 | 127 | 8% |
| Kassanjee (2022) (45) | South Africa | 3005 | 2999 | 1862 | 62% |
| Kelly-Hanku (2020) (46) | Papua New Guinea | 2091 | 848 | 83 | 10% |
| Kerrigan (2017) (47) | Tanzania | 496 | 62 | 62 | 100% |
| Kumar (2014) (48) | India | 4074 | 3873 | 551 | 14% |
| Leddy (2018) (49) | Tanzania | 496 | 496 | 203 | 41% |
| Lima (2017) (50) | Brazil | 2523 | 2523 | NR | NR |
| Lyons (2017) (51) | Côte d’Ivoire | 466 | 453 | 50 | 11% |
| Lyons (2020) (52) | Multi-country | 7259 | 7259 | 2070 | 29% |
| MacLin (2023) (53) | Dominican Republic | 311 | 211 | 211 | 100% |
| Mendoza (2017) (54) | Dominican Republic | 268 | 268 | 268 | 100% |
| Mizinduko (2020) (55) | Tanzania | 958 | 958 | 169 | 18% |
| Mountain (2018) (56) | Kenya | 408 | 346 | 34 | 10% |
| Mukherjee (2022) (57) | Kazakhstan | 255 | 255 | 64 | 25% |
| Mukuku (2024) (58) | Democratic Republic of the Congo | 572 | 572 | 19 | 3% |
| Mulholland (2022) (59) | multi-country | 786 | 715 | 85 | 12% |
| Mutagoma (2017) (60) | Rwanda | 1338 | 1112 | 565 | 51% |
| Mutagoma (2019) (61) | Rwanda | 1978 | 1978 | 819 | 42% |
| Nabayinda (2023) (62) | Uganda | 542 | 542 | 220 | 41% |
| Namale (2019) (63) | Uganda | 603 | 432 | 432 | 100% |
| Oldenburg (2018) (64) | Zambia | 964 | 898 | 234 | 24% |
| Ouma (2021) (65) | Uganda | 300 | 300 | 126 | 42% |
| Pando (2013) (66) | Argentina | 1255 | 870 | NR | NR |
| Prangnell (2018) (67) | Canada | 259 | 259 | 89 | 34% |
| Ramesh (2012) (68) | India | 2042 | 2042 | 371 | 18% |
| Rice (2022) (69) | Zimbabwe | 605 | 605 | 122 | 20% |
| Schwartz (2016) (70) | South Africa | 410 | 163 | 163 | 100% |
| Sherman (2019) (71) | USA | 262 | 249 | 13 | 5% |
| Shokoohi (2017) (72) | Iran | 1337 | 1295 | 0 | 0% |
| Shrestha (2017) (73) | Nepal | 610 | 610 | 6 | 1% |
| Sileo (2018) (74) | Uganda | 115 | 115 | NR | NR |
| Strathdee (2011) (75) | Mexico | 620 | 620 | 33 | 5% |
| Szwarcwald (2017) (76) | Brazil | 4245 | 4245 | 225 | 5% |
| Tokar (2013) (77) | Ukraine | 5023 | 5023 | NR | NR |
| Tounkara (2014) (78) | Benin | 981 | 981 | 200 | 20% |
| Tuot (2020) (79) | Cambodia | 3149 | 3149 | 72 | 3% |
| Ulibarri (2011) (80) | Mexico | 924 | 924 | 55 | 6% |
| Vandenhoudt (2013) (81) | Kenya | 481 | 481 | 277 | 58% |
| Vu (2017) (82) | Tanzania | 1914 | 1906 | 535 | 28% |
| Wand (2015) (83) | Papua New Guinea | 523 | 203 | 14 | 7% |
| Wang (2021) (84) | Malaysia | 299 | 283 | 17 | 6% |
| West (2022) (85) | Mexico | 584 | 584 | 0 | 0% |
| Wilson (2016a) (86) | Kenya | 195 | 195 | 195 | 100% |
| Wilson (2016b) (87) | Kenya | 357 | 356 | 357 | 100% |
| Wirth (2013) (88) | India | 1814 | 1814 | 310 | 18% |
| Xie (2023) (89) | China | 480 | 480 | NR | NR |
| Zalla (2019) (90) | Haiti | 990 | 958 | 75 | 8% |
| Zhang (2012) (91) | China | 1022 | 937 | NR | NR |

### Appendix 12. Estimates of the association between sexual violence and HIV outcomes

| **Author (year)** | **Violence definition** | **Violence time frame** | **Exposed, n (%)** | **Perpetrator** | **HIV outcome** | **Effect size (95% CI)** |
| --- | --- | --- | --- | --- | --- | --- |
| Alemu (2022) | Sexual assault | Ever | 44 (12%) | Unspecified | HIV prevalence | OR: 1.60 (0.78–3.28); aOR: 1.59 (0.76–3.33) |
| Amogne (2019) | Rape | Ever | 742 (15%) | Client or sexual partner | HIV prevalence | OR: 0.88 (0.73–1.07) |
| Arimide (2022) | Forced into selling sex | Ever | 26 (11%) | Unspecified | Viral suppression | OR: 0.33 (0.14–0.76); aOR: 0.36 (0.14–0.93) |
| Arumugam (2022) | Forced sexual intercourse | Past 12 months | 4847 (18%) | Unspecified | HIV prevalence | OR: 0.96 (0.77–1.20) |
| Ayamah (2023) | Sexual violence | Past 12 months | 1059 (25%) | Unspecified | HIV prevalence | OR: 0.94 (0.69–1.29) |
| Ayamah (2023) | Sexual violence | Past 12 months | 1059 (25%) | Unspecified | HIV testing | OR: 0.91 (0.77–1.07) |
| Bowring (2019) | Forced to have sex against will | Past 6 months | 202 (10%) | Unspecified | HIV prevalence | OR: 1.06 (0.69–1.61) |
| Braunstein (2011) | History of forced sex | Ever | 202 (25%) | Unspecified | HIV incidence | OR: 2.26 (0.92–5.58); aOR: 2.30 (0.90–5.80) |
| Braunstein (2011) | History of forced sex | Ever | 202 (25%) | Unspecified | HIV prevalence | OR: 2.21 (1.55–3.13); aOR: 2.20 (1.50–3.20) |
| Budhwani (2017) | Sexually abused | Ever | 82 (24%) | Unspecified | HIV prevalence | OR: 1.07 (0.11–10.43) |
| Bugssa (2015) | Sexually abused | Ever | 44 (14%) | Unspecified | HIV prevalence | OR: 3.10 (1.40–6.70); aOR: 2.20 (1.40–6.30) |
| Chabata (2019) | Forced to have sexual intercourse | Ever | 44 (9%) | Unspecified | HIV prevalence | OR: 1.32 (0.57–3.06) |
| Coetzee (2017) | Sexual violence by non-intimate partner | Ever | 277 (55%) | Unspecified | HIV prevalence | OR: 1.64 (1.15–2.33) |
| Comins (2024) | Forced to have sex | Ever | 515 (38%) | Unspecified | Viral suppression | OR: 0.71 (0.59–0.85); aOR: 0.87 (0.71–1.08) |
| Damacena (2011) | Physically forced sex | Ever | NR | Unspecified | HIV prevalence | OR: 1.74 (1.00–3.03) |
| Decker (2014) | Forced or coerced vaginal sex | Past 6 months | 86 (11%) | Client | HIV prevalence | OR: 3.79 (1.67–8.61); aOR: 3.77 (1.73–8.22) |
| Decker (2014) | Sex to avoid incarceration or arrest | Past 6 months | 23 (3%) | Police | HIV prevalence | OR: 4.07 (1.14–14.56); aOR: 3.45 (0.92–12.93) |
| Decker (2014) | Forced or coerced anal sex | Past 6 months | 47 (6%) | Client | HIV prevalence | OR: 5.45 (2.20–13.51); aOR: 4.80 (1.89–12.19) |
| Decker (2012) | Sex for leniency from police | Past 3 months | 52 (37%) | Police | HIV prevalence | OR: 2.42 (0.52–11.25) |
| Decker (2012) | Sexual violence | Past 12 months | 5 (4%) | Pimp | HIV prevalence | OR: 1.84 (0.09–36.95) |
| Doshi (2018) | Forced into sex work | Ever | 222 (65%) | Unspecified | Viral suppression | OR: 0.67 (0.38–1.20) |
| Doshi (2018) | Forced into sex work | Ever | 71 (33%) | Unspecified | Current ART | OR: 0.98 (0.40–2.43) |
| Faini (2022) | Rape | Past 3 months | 142 (18%) | Unspecified | HIV prevalence | OR: 2.19 (1.21–3.95) |
| Goldenberg (2013) | Involuntary sex exchange | Past month | 31 (14%) | Client | HIV prevalence | OR: 1.73 (0.34–8.76) |
| Guure (2023) | Forced sex | Ever | 986 (15%) | Unspecified | HIV prevalence | OR: 1.26 (0.91–1.74); aOR: 1.38 (1.02–1.89) |
| Hensen (2019) | Experienced sexual violence | Past 12 months | 390 (16%) | Unspecified | HIV prevalence | OR: 1.20 (0.86–1.68) |
| Hladik (2017) | Raped | Past month | 243 (26%) | Client | HIV prevalence | OR: 1.03 (0.76–1.41) |
| Hladik (2017) | Raped | Past month | 308 (33%) | Unspecified | HIV prevalence | OR: 1.10 (0.82–1.46) |
| Izadi (2023) | Violent sex | Ever | 600 (40%) | Unspecified | HIV prevalence | OR: 1.81 (0.80–4.06) |
| Kakisingi (2020) | Sexual violence due to selling sex | Past 3 months | 32 (2%) | Unspecified | HIV prevalence | OR: 1.17 (0.35–3.89); aOR: 0.74 (0.21–2.62) |
| Kassanjee (2022) | Sexual violence | Past 12 months | 1447 (48%) | Client | HIV incidence | OR: 1.42 (0.04–52.04) |
| Kelly-Hanku (2020) | Sexual violence | Past 12 months | 109 (16%) | Unspecified | HIV prevalence | OR: 2.80 (1.30–6.10) |
| Lyons (2017) | Forced sex | Ever | 195 (43%) | Unspecified | HIV prevalence | OR: 0.99 (0.54–1.80) |
| Lyons (2017) | Forced sex | Ever | 200 (43%) | Unspecified | HIV testing | OR: 1.27 (0.78–2.06) |
| Lyons (2020) | Forced sex | Ever | 2207 (30%) | Unspecified | HIV prevalence | OR: 1.29 (1.16–1.44); aOR: 1.32 (1.13–1.54) |
| Mizinduko (2020) | Forced sex | Past 12 months | 312 (33%) | Unspecified | HIV prevalence | OR: 1.45 (0.98–2.15); aOR: 1.94 (1.34–2.82) |
| Mountain (2018) | Forced sex | Ever | 119 (30%) | Unspecified | HIV prevalence | OR: 1.56 (0.89–2.72); aOR: 1.44 (0.81–2.56) |
| Mountain (2018) | Forced sex | Ever | 119 (30%) | Unspecified | HIV testing | OR: 0.81 (0.66–1.00); aOR: 0.82 (0.66–1.01) |
| Mukuku (2024) | Forced sex | Past 12 months | 29 (5%) | Unspecified | HIV prevalence | OR: 10.60 (3.70–30.50); aOR: 12.20 (3.20–46.40) |
| Mulholland (2022) | Forced sex | Ever | 158 (20%) | Unspecified | HIV prevalence | OR: 1.13 (0.64–2.00) |
| Mutagoma (2019) | Sexual violence | Ever | 361 (18%) | Unspecified | HIV prevalence | OR: 1.40 (1.28–1.57) |
| Oldenburg (2018) | Sexual intimate partner violence | Past 12 months | 30 (47%) | Intimate partner | ART initiation | aOR: 0.42 (0.22–0.77) |
| Pando (2013) | Sexual abuse during lifetime | Ever | 182 (20%) | Unspecified | HIV prevalence | OR: 2.40 (0.90–6.60); aOR: 1.50 (0.90–2.50) |
| Pando (2013) | Forced first sex or sexual abuse in lifetime | Ever | 219 (24%) | Unspecified | HIV prevalence | OR: 2.10 (0.80–5.80); aOR: 1.40 (0.80–2.40) |
| Rice (2022) | Sexual violence/abuse | Past 12 months | 124 (20%) | Unspecified | HIV prevalence | OR: 1.40 (0.80–2.45) |
| Sherman (2019) | Childhood sexual abuse (CGW) | Ever | NR | Family or friend | HIV prevalence | OR: 0.57 (0.18–1.80) |
| Sherman (2019) | Childhood sexual abuse (TGW) | Ever | NR | Family or friend | HIV prevalence | OR: 3.39 (2.73–4.21); aOR: 4.56 (1.20–17.32) |
| Shokoohi (2017) | Sexual violence | Ever | 512 (40%) | Unspecified | HIV testing | OR: 1.09 (0.97–1.22); aOR: 1.02 (0.91–1.14) |
| Shrestha (2017) | Forced sex | Past 12 months | 125 (20%) | Unspecified | HIV testing | OR: 1.99 (1.32–3.01); aOR: 1.10 (1.00–1.30) |
| Sileo (2018) | Sexual violence | Ever | 71 (62%) | Intimate partner | HIV prevalence | OR: 3.33 (1.11–10.03); aOR: 3.94 (1.22–12.66) |
| Sileo (2018) | Sexual violence | Past 12 months | 62 (54%) | Intimate partner | HIV prevalence | OR: 1.67 (0.66–4.21) |
| Strathdee (2011) | Sexually abused as a child | Ever | 205 (33%) | Family or friend | HIV prevalence | OR: 0.77 (0.35–1.69) |
| Strathdee (2011) | Sexually abused/raped | Past month | 34 (6%) | Intimate partner | HIV prevalence | OR: 1.13 (0.26–4.95) |
| Strathdee (2011) | Sexually abused/raped | Past month | 138 (23%) | Client | HIV prevalence | OR: 1.10 (0.48–2.49) |
| Szwarcwald (2017) | Sexual violence | Ever | NR | Unspecified | HIV prevalence | OR: 1.54 (1.05–2.26) |
| Tounkara (2014) | Forced sex | Past month | 132 (14%) | Unspecified | HIV prevalence | OR: 1.56 (1.03–2.37); aOR: 1.42 (1.02–1.98) |
| Tuot (2020) | Gang rape | Past 3 months | 40 (1%) | Unspecified | HIV prevalence | OR: 4.48 (1.72–11.70) |
| Tuot (2020) | Forced sex | Past 3 months | 42 (1%) | Unspecified | HIV prevalence | OR: 5.28 (2.17–12.84) |
| Ulibarri (2011) | Sexual abuse | Past 6 months | 88 (10%) | Client | HIV prevalence | OR: 3.50 (1.66–7.36); aOR: 3.00 (1.30–6.92) |
| Vandenhoudt (2013) | Raped as FSW | Ever | 193 (40%) | Unspecified | HIV prevalence | OR: 0.91 (0.63–1.32); aOR: 0.90 (0.60–1.30) |
| Vu (2017) | Sexually abused | Past 12 months | 485 (25%) | Client | HIV prevalence | OR: 1.10 (0.80–1.50) |
| Wand (2015) | Raped | Past 12 months | 74 (14%) | Unspecified | HIV prevalence | OR: 3.48 (1.08–11.21); aOR: 3.42 (1.06–11.06) |
| Wang (2021) | Childhood sexual assault (CWSW) | Ever | 77 (27%) | Unspecified | HIV prevalence | OR: 1.12 (0.38–3.28) |
| Wang (2021) | Adulthood sexual assault (TGWSW) | Ever | 44 (24%) | Unspecified | HIV prevalence | OR: 3.26 (1.17–9.05); aOR: 4.06 (0.68–24.19) |
| Wang (2021) | Childhood sexual assault (TGWSW) | Ever | 89 (48%) | Unspecified | HIV prevalence | OR: 3.98 (1.25–12.17); aOR: 0.92 (0.12–6.75) |
| West (2022) | Material violence and sexual violence | Past 6 months | 92 (16%) | Police | HIV testing | OR: 1.68 (1.02–2.74) |
| Wirth (2013) | Forced prostitution | Ever | 107 (6%) | Unspecified | HIV prevalence | OR: 2.74 (1.41–5.31); aOR: 2.30 (1.08–4.90) |
| Zalla (2019) | Forced to have sex | Ever | 384 (40%) | Unspecified | HIV prevalence | OR: 2.07 (0.32–13.45) |
| Zhang (2012) | Unwanted sexual intercourse | Ever | 135 (14%) | Client | HIV testing | OR: 0.65 (0.45–0.95) |
| Zhang (2012) | Unwanted sexual intercourse | Ever | 120 (16%) | Intimate partner | HIV testing | OR: 0.59 (0.39–0.88) |

### Appendix 13. Estimates of the association between physical violence and HIV outcomes

| **Author (year)** | **Violence definition** | **Violence time frame** | **Exposed, n (%)** | **Perpetrator** | **HIV outcome** | **Effect size (95% CI)** |
| --- | --- | --- | --- | --- | --- | --- |
| Abdella (2022) | Physical violence | Past 12 months | 1356 (22%) | Unspecified | HIV prevalence | OR: 1.06 (0.91–1.24); aOR: 1.00 (0.80–1.10) |
| Amogne (2019) | Physical beating | Past 12 months | 855 (18%) | Client or sexual partner | HIV prevalence | OR: 1.04 (0.88–1.24) |
| Arimide (2022) | Physically beaten | Past 12 months | 30 (13%) | Unspecified | Viral suppression | OR: 0.88 (0.37–2.13) |
| Arumugam (2022) | Physically beaten | Past 12 months | 6500 (24%) | Unspecified | HIV prevalence | OR: 1.01 (0.83–1.23) |
| Ayamah (2023) | Physical violence | Past 12 months | 433 (10%) | Unspecified | HIV prevalence | OR: 1.00 (0.64–1.54) |
| Ayamah (2023) | Physical violence | Past 12 months | 433 (10%) | Unspecified | HIV testing | OR: 0.91 (0.72–1.14) |
| Bowring (2019) | Physical harassment/hurt due to sex work | Past 6 months | 232 (12%) | Unspecified | HIV prevalence | OR: 0.65 (0.41–1.03) |
| Chabata (2019) | Physical violence | Ever | 146 (29%) | Client | HIV prevalence | OR: 1.08 (0.54–1.77) |
| Chabata (2019) | Physical violence | Ever | 233 (46%) | Intimate partner | HIV prevalence | OR: 0.97 (0.58–1.63) |
| Comins (2024) | Physical violence | Ever | 740 (54%) | Unspecified | Viral suppression | OR: 0.79 (0.66–0.93); aOR: 0.99 (0.82–1.20) |
| Decker (2014) | Physical violence | Ever | 74 (18%) | Pimp | HIV prevalence | OR: 9.97 (2.92–34.07); aOR: 6.32 (1.85–21.63) |
| Decker (2014) | Physical violence | Past 6 months | 77 (10%) | Client | HIV prevalence | OR: 2.98 (1.23–7.22); aOR: 2.52 (1.41–4.51) |
| Decker (2012) | Threats of physical violence | Past 12 months | 14 (10%) | Pimp | HIV prevalence | OR: 16.36 (2.47–108.62) |
| Decker (2012) | Physical violence | Past 12 months | 107 (76%) | Client | HIV prevalence | OR: 0.78 (0.15–4.24) |
| Decker (2012) | Physical violence | Past 12 months | 12 (8%) | Pimp | HIV prevalence | OR: 0.68 (0.04–12.62) |
| Goldenberg (2019) | Intimate partner violence | Past 6 months | 47 (53%) | Intimate partner | Current ART | OR: 0.80 (0.35–1.84) |
| Hensen (2019) | Physical violence | Ever | 972 (39%) | Intimate partner | HIV prevalence | OR: 1.44 (1.12–1.86); aOR: 1.37 (1.05–1.78) |
| Kelly-Hanku (2020) | Physical violence | Past 12 months | 146 (17%) | Unspecified | HIV prevalence | OR: 2.30 (1.20–4.20); aOR: 2.50 (1.20–4.90) |
| Lima (2017) | Physical violence | Past 12 months | 295 (12%) | Clients | HIV prevalence | OR: 1.79 (1.04–3.08) |
| Lima (2017) | Physical violence | Past 12 months | 419 (17%) | Family member | HIV prevalence | OR: 1.41 (0.86–2.29) |
| Lima (2017) | Physical violence | Past 12 months | 636 (25%) | Intimate partner | HIV prevalence | OR: 1.36 (0.94–1.97) |
| Lima (2017) | Physical violence | Past 12 months | 961 (38%) | Any perpetrator | HIV prevalence | OR: 1.26 (0.97–1.64) |
| Lima (2017) | Physical violence | Past 12 months | 199 (8%) | Police | HIV prevalence | OR: 1.82 (1.05–3.16) |
| Lyons (2017) | Physically hurt | Ever | 240 (53%) | Unspecified | HIV prevalence | OR: 1.83 (1.00–3.41) |
| Lyons (2017) | Physically hurt | Ever | 247 (53%) | Unspecified | HIV testing | OR: 1.02 (0.54–1.65) |
| Lyons (2020) | Physical violence | Ever | 2359 (33%) | Unspecified | HIV prevalence | OR: 1.58 (1.42–1.76); aOR: 1.23 (1.02–1.49) |
| Mountain (2018) | Physical hurt by sex partner | Ever | 123 (30%) | Sexual partner | HIV prevalence | OR: 2.04 (1.27–4.27); aOR: 1.89 (1.15–4.10) |
| Mountain (2018) | Police assault or arrest | Ever | 183 (45%) | Police | HIV prevalence | OR: 1.61 (0.81–4.20); aOR: 1.61 (0.80–4.21) |
| Mountain (2018) | Physical hurt by sex partner | Ever | 123 (30%) | Sexual partner | HIV testing | OR: 0.83 (0.68–1.01); aOR: 0.84 (0.68–1.03) |
| Mountain (2018) | Police assault or arrest | Ever | 183 (45%) | Police | HIV testing | OR: 0.85 (0.69–1.04); aOR: 0.84 (0.68–1.04) |
| Mulholland (2022) | Physical violence | Ever | 250 (32%) | Intimate partner | HIV prevalence | OR: 1.03 (0.62–1.71) |
| Mutagoma (2019) | Physical violence | Ever | 702 (36%) | Unspecified | HIV prevalence | OR: 1.50 (1.11–2.05) |
| Namale (2019) | Physical violence from sexual partner(s) | Past 3 months | 148 (34%) | Intimate partner | Viral suppression | OR: 0.80 (0.40–1.60) |
| Oldenburg (2018) | Physical intimate partner violence | Past 12 months | 25 (50%) | Intimate partner | ART initiation | aOR: 0.65 (0.35–1.21) |
| Pando (2013) | Beaten because of sex work | Ever | 267 (22%) | Unspecified | HIV prevalence | OR: 1.90 (0.80–4.50); aOR: 1.20 (0.80–1.90) |
| Rice (2022) | Physical violence/abuse | Past 12 months | 133 (22%) | Client | HIV prevalence | OR: 2.53 (1.48–4.32) |
| Rice (2022) | Physical violence/abuse | Past 12 months | 151 (25%) | Intimate partner | HIV prevalence | OR: 1.51 (0.89–2.58); aOR: 1.97 (1.05–3.67) |
| Rice (2022) | Physical violence | Past 12 months | 50 (8%) | Police | HIV prevalence | OR: 1.23 (0.52–2.92) |
| Schwartz (2016) | Physical abuse/violence | Past 12 months | NR | Unspecified | Current ART | OR: 0.84 (0.64–1.10); aOR: 0.87 (0.67–1.13) |
| Sherman (2019) | Childhood physical abuse | Ever | NR | Family or friend | HIV prevalence | OR: 1.17 (0.39–3.53) |
| Shrestha (2017) | Physical assault | Past 12 months | 81 (13%) | Unspecified | HIV testing | OR: 1.56 (0.98–2.50); aOR: 0.80 (0.60–1.00) |
| Sileo (2018) | Physical violence | Ever | 59 (51%) | Intimate partner | HIV prevalence | OR: 1.16 (0.47–2.87) |
| Sileo (2018) | Physical violence | Past 12 months | 46 (40%) | Intimate partner | HIV prevalence | OR: 1.05 (0.43–2.56) |
| Strathdee (2011) | Physically abused as a child | Ever | 150 (25%) | Family or friend | HIV prevalence | OR: 0.85 (0.36–2.02) |
| Szwarcwald (2017) | Physical violence | Ever | NR | Unspecified | HIV prevalence | OR: 1.08 (0.73–1.61) |
| Tounkara (2014) | Physical violence | Past month | 168 (17%) | Unspecified | HIV prevalence | OR: 1.32 (0.89–1.96); aOR: 1.45 (1.05–2.00) |
| Vu (2017) | Physically abused | Past 12 months | 656 (34%) | Client | HIV prevalence | OR: 0.80 (0.60–1.10) |
| Wang (2021) | Adulthood physical assault (CWSW) | Ever | 99 (35%) | Unspecified | HIV prevalence | OR: 2.22 (0.83–5.96) |
| Wang (2021) | Childhood physical assault (CWSW) | Ever | 101 (36%) | Unspecified | HIV prevalence | OR: 1.68 (0.63–4.50) |
| Wang (2021) | Physical assault because of sex work (CWSW) | Ever | 26 (9%) | Unspecified | HIV prevalence | OR: 4.80 (1.54–14.93); aOR: 2.59 (0.67–10.19) |
| Wang (2021) | Physical assault because of sex work (TGWSW) | Ever | 37 (20%) | Unspecified | HIV prevalence | OR: 1.27 (0.39–4.14) |
| Wang (2021) | Adulthood physical assault (TGWSW) | Ever | 61 (33%) | Unspecified | HIV prevalence | OR: 0.61 (0.19–1.94) |
| Wang (2021) | Childhood physical assault (TGWSW) | Ever | 74 (40%) | Unspecified | HIV prevalence | OR: 1.80 (0.66–4.90) |
| Zhang (2012) | Physically hurt | Ever | 126 (17%) | Client | HIV testing | OR: 1.13 (0.77–1.65) |
| Zhang (2012) | Physically hurt | Ever | 148 (20%) | Intimate partner | HIV testing | OR: 0.80 (0.56–1.15) |

### Appendix 14. Estimates of the association between physical and/or sexual violence and HIV outcomes

| **Author (year)** | **Violence definition** | **Violence time frame** | **Exposed, n (%)** | **Perpetrator** | **HIV outcome** | **Effect size (95% CI)** |
| --- | --- | --- | --- | --- | --- | --- |
| Aho (2013) | Violence episode | Past 3 months | 53 (24%) | Client | HIV prevalence | OR: 1.17 (0.62–2.22) |
| Alary (2014) | Experienced violence | Ever | 2190 (10%) | Unspecified | HIV prevalence | aOR: 1.35 (1.19–1.53) |
| Beattie (2015) | Beaten or forced to have sex | Past 12 months | 416 (11%) | Unspecified | HIV prevalence | OR: 1.87 (1.40–2.49); aOR: 1.59 (1.18–2.15) |
| Beattie (2015) | Beaten or forced to have sex | Past 12 months | 416 (11%) | Unspecified | HIV testing | OR: 1.02 (0.78–1.33); aOR: 1.09 (0.81–1.48) |
| Beattie (2010) | Beaten or forced to have sex | Past 12 months | 413 (11%) | Unspecified | HIV prevalence | OR: 1.10 (0.80–1.49); aOR: 0.96 (0.70–1.32) |
| Beksinska (2018) | Domestic violence | Past 12 months | 216 (19%) | Intimate partner | HIV prevalence | OR: 0.28 (0.11–0.73); aOR: 0.40 (0.15–1.09) |
| Beksinska (2018) | Domestic and workplace/community violence | Past 12 months | 69 (6%) | Unspecified | HIV prevalence | OR: 1.11 (0.40–3.38); aOR: 1.32 (0.41–4.29) |
| Beksinska (2018) | Workplace or community violence | Past 12 months | 80 (7%) | Unspecified | HIV prevalence | OR: 1.75 (0.88–3.50); aOR: 1.16 (0.55–2.44) |
| Beksinska (2018) | Any violence | Past 12 months | 365 (33%) | Unspecified | HIV prevalence | OR: 0.73 (0.42–1.27); aOR: 0.82 (0.44–1.53) |
| Berger (2018) | Beaten or forced to have sex | Ever | 156.816 (48%) | Unspecified | HIV prevalence | OR: 1.09 (0.67–1.78) |
| Berger (2018) | Beaten or forced to have sex | Ever | 133 (41%) | Unspecified | HIV testing | OR: 1.15 (0.70–1.91) |
| Bhardwaj (2023) | Experienced physical or sexual violence and moderate to severe drug use risk | Ever | 638 (46%) | Unspecified | Viral suppression | aOR: 0.88 (0.79–0.99) |
| Budhwani (2021) | Beaten or raped in past year | Past 12 months | 64 (22%) | Unspecified | HIV prevalence | aOR: 3.15 (1.19–8.34) |
| Coetzee (2017) | Intimate partner physical/sexual violence | Ever | 288 (57%) | Intimate partner | HIV prevalence | OR: 0.95 (0.66–1.35) |
| Davey (2020) | Physically hurt or raped | Past month | 637 (23%) | Unspecified | Viral suppression | OR: 0.87 (0.59–1.28); aOR: 1.00 (0.93–1.07) |
| Davey (2020) | Physically hurt or raped | Past month | 637 (23%) | Any | Current ART | OR: 0.80 (0.55–1.15); aOR: 0.96 (0.90–1.03) |
| Davey (2020) | Physically hurt | Past month | 637 (23%) | Unspecified | HIV testing | OR: 1.07 (0.80–1.43); aOR: 0.99 (0.87–1.12) |
| Decker (2016) | Beaten up, physically hurt or forced to have sex | Ever | 1098 (60%) | Unspecified | Current ART | OR: 0.59 (0.20–1.72); aOR: 0.49 (0.13–1.82) |
| Decker (2016) | Beaten or forced to have sex | Ever | 1098 (60%) | Unspecified | HIV testing | OR: 0.83 (0.66–1.05); aOR: 0.92 (0.71–1.19) |
| Duff (2016) | Physical/sexual violence | Past 6 months | 13 (18%) | Client | Viral suppression | OR: 0.50 (0.20–1.23) |
| Emmanuel (2021) | Faced violence | Past 12 months | NR | Unspecified | HIV prevalence | OR: 1.10 (0.70–1.50) |
| Goldenberg (2019) | Sexual/physical abuse | Past 6 months | 39 (44%) | Client | Current ART | OR: 0.66 (0.28–1.53) |
| Goldenberg (2016) | Physical/sexual violence | Past 6 months | 10 (14%) | Client | ART adherence | OR: 0.71 (0.17–3.03) |
| Goldenberg (2016) | Verbal, physical, or sexual violence | Past 6 months | 37 (9%) | Police | HIV prevalence | OR: 1.07 (0.53–2.18) |
| Goldenberg (2016) | Verbal, physical, or sexual violence | Past 6 months | 66 (16%) | Intimate partner | HIV prevalence | OR: 0.83 (0.47–1.47) |
| Goldenberg (2016) | Physical or sexual violence | Past 6 months | 314 (78%) | Client | HIV prevalence | OR: 0.82 (0.50–1.35) |
| Hendrickson (2018) | Physical or sexual GBV | Past 6 months | 197 (40%) | Unspecified | HIV prevalence | OR: 1.15 (0.82–1.61); aOR: 1.37 (0.92–2.04) |
| Hladik (2017) | Violence due to selling sex | Ever | 433 (46%) | Unspecified | HIV prevalence | OR: 1.34 (1.02–1.75) |
| Iakunchykova (2017) | Any violence during sex work | Ever | NR | Unspecified | HIV prevalence | OR: 1.91 (1.48–2.47); aOR: 1.45 (1.09–1.93) |
| Jones (2023) | NA | Ever | 1292 (20%) | Any perpretator | HIV incidence | OR: 1.02 (0.81–1.27); aOR: 1.08 (0.87–1.35) |
| Kerrigan (2017) | Gender-based violence | Ever | 39 (68%) | Unspecified | Viral suppression | OR: 0.59 (0.20–1.79) |
| Kerrigan (2017) | Gender-based violence | Ever | 252 (51%) | Unspecified | HIV prevalence | OR: 1.18 (0.80–1.73) |
| Kumar (2014) | Violence or forced sex | Past 12 months | 960 (25%) | Unspecified | HIV prevalence | OR: 1.74 (1.42–2.14); aOR: 1.76 (1.43–2.16) |
| Leddy (2018) | Physical or sexual GBV | Past 6 months | 197 (40%) | Unspecified | HIV prevalence | OR: 1.65 (0.95–2.86) |
| MacLin (2023) | SW-related violence and harassment | Past 6 months | 18 (18%) | Unspecified | Viral suppression | OR: 1.43 (0.43–4.76); aOR: 1.56 (0.46–5.23) |
| MacLin (2023) | SW-related violence and harassment | Past 6 months | 18 (18%) | Unspecified | ART adherence | OR: 3.12 (0.36–25.00); aOR: 2.44 (0.27–20.00) |
| MacLin (2023) | SW-related violence and harassment | Past 6 months | 18 (18%) | Unspecified | ART adherence | OR: 0.91 (0.29–2.88); aOR: 0.82 (0.26–2.64) |
| MacLin (2023) | SW-related violence and harassment | Past 6 months | 18 (18%) | Unspecified | Current ART | OR: 1.25 (0.30–5.27); aOR: 1.27 (0.30–5.48) |
| Mendoza (2017) | Violence | Past 6 months | 33 (12%) | Intimate partner | ART adherence | OR: 0.27 (0.12–0.64); aOR: 0.19 (0.07–0.52) |
| Mendoza (2017) | Violence | Past 6 months | 49 (18%) | Sexual partner | ART adherence | OR: 0.29 (0.14–0.60); aOR: 0.27 (0.11–0.63) |
| Mendoza (2017) | Violence | Past 6 months | 23 (8%) | Client | ART adherence | OR: 0.37 (0.13–1.09); aOR: 0.47 (0.14–1.52) |
| Mendoza (2017) | Violence | Past 6 months | 33 (12%) | Intimate partner | ART coverage | OR: 0.48 (0.21–1.11); aOR: 0.45 (0.17–1.18) |
| Mendoza (2017) | Violence | Past 6 months | 33 (12%) | Intimate partner | ART coverage | OR: 0.83 (0.35–1.96); aOR: 0.94 (0.38–2.38) |
| Mendoza (2017) | Violence | Past 6 months | 49 (18%) | Sexual partner | ART coverage | OR: 0.31 (0.15–0.65); aOR: 0.31 (0.14–0.71) |
| Mendoza (2017) | Violence | Past 6 months | 49 (18%) | Sexual partner | ART coverage | OR: 0.80 (0.39–1.67); aOR: 0.90 (0.41–2.00) |
| Mendoza (2017) | Violence | Past 6 months | 23 (8%) | Client | ART coverage | OR: 0.18 (0.05–0.67); aOR: 0.18 (0.05–0.67) |
| Mendoza (2017) | Violence | Past 6 months | 23 (8%) | Client | ART coverage | OR: 0.47 (0.19–1.18); aOR: 0.47 (0.18–1.27) |
| Mendoza (2017) | Violence | Past 6 months | 33 (12%) | Intimate partner | Current ART | OR: 0.25 (0.08–0.80); aOR: 0.25 (0.06–1.00) |
| Mendoza (2017) | Violence | Past 6 months | 49 (18%) | Sexual partner | Current ART | OR: 0.32 (0.11–0.93); aOR: 0.41 (0.12–1.41) |
| Mendoza (2017) | Violence | Past 6 months | 23 (8%) | Client | Current ART | OR: 0.86 (0.11–6.97); aOR: 0.40 (0.04–3.90) |
| Mountain (2018) | Forced sex, physical violence and police assault or arrest | Ever | 44 (11%) | Unspecified | HIV prevalence | OR: 2.51 (1.11–5.64); aOR: 2.65 (1.19–5.90) |
| Mountain (2018) | Forced sex, physical violence and police assault or arrest | Ever | 44 (11%) | Unspecified | HIV testing | OR: 0.56 (0.37–0.84); aOR: 0.56 (0.37–0.85) |
| Mutagoma (2017) | Violence/harassment | Ever | 905 (51%) | Unspecified | HIV prevalence | OR: 1.00 (0.80–1.20) |
| Nabayinda (2023) | Intimate-partner violence | Past 3 months | 248 (46%) | Intimate partner | HIV prevalence | OR: 0.96 (0.68–1.35) |
| Oldenburg (2018) | Physical or sexual intimate partner violence | Past 12 months | 142 (61%) | Intimate partner | ART initiation | aOR: 0.40 (0.22–0.72) |
| Ouma (2021) | Gender-based violence | Ever | 183 (61%) | Client | HIV prevalence | OR: 1.72 (1.07–2.79) |
| Pando (2013) | Three violent experiences (vs. none) | Ever | 151 (11%) | Unspecified | HIV prevalence | OR: 5.50 (0.50–60.40); aOR: 1.20 (0.30–4.20) |
| Pando (2013) | Two violent experiences (vs. none) | Ever | 330 (21%) | Unspecified | HIV prevalence | OR: 5.90 (0.70–52.20); aOR: 1.30 (0.50–3.40) |
| Pando (2013) | One violent experience (vs. none) | Ever | 607 (32%) | Unspecified | HIV prevalence | OR: 3.80 (0.40–33.80); aOR: 0.80 (0.30–2.30) |
| Pando (2013) | Four or more violent experiences (vs. none) | Ever | 56 (6%) | Unspecified | HIV prevalence | OR: 14.10 (1.50–133.00); aOR: 2.90 (0.90–9.20) |
| Prangnell (2018) | Physical, sexual, or verbal violence | Past 6 months | 32 (12%) | Client | HIV prevalence | OR: 0.62 (0.27–1.46) |
| Ramesh (2012) | Physically beaten or forced to have sex | Past 12 months | 352 (17%) | Unspecified | HIV prevalence | OR: 1.72 (1.31–2.26); aOR: 1.58 (1.20–2.09) |
| Tokar (2013) | Personal experience of violence | NA | NR | Police | HIV prevalence | OR: 1.40 (1.00–1.80) |
| Tokar (2013) | Personal experience of violence | NA | NR | Intimate partner | HIV prevalence | OR: 1.80 (1.30–2.60) |
| Tokar (2013) | Personal experience of violence | NA | NR | Unspecified | HIV prevalence | OR: 1.80 (1.50–2.10) |
| Wilson (2016) | Intimate-partner violence | Past 12 months | 45 (23%) | Intimate partner | Viral suppression | OR: 4.42 (1.36–14.38); aOR: 4.76 (1.19–20.00) |
| Wilson (2016) | Intimate-partner violence | Past 12 months | 45 (23%) | Intimate partner | ART adherence | OR: 0.51 (0.18–1.45); aOR: 0.63 (0.21–1.92) |
| Wilson (2016) | Intimate-partner violence | Past 12 months | 45 (23%) | Intimate partner | ART adherence | OR: 1.18 (0.91–1.52); aOR: 1.19 (0.88–1.64) |
| Wilson (2016) | Intimate-partner violence | Past 12 months | 52 (15%) | Intimate partner | Current ART | OR: 0.86 (0.58–1.53) |
| Xie (2023) | Violence | Ever | 65 (14%) | Client | HIV testing | OR: 2.83 (1.65–4.84) |
| Zalla (2019) | Intimate-partner violence | Past 12 months | 305 (32%) | Intimate partner | HIV prevalence | OR: 3.72 (0.36–38.21) |
| Zhang (2012) | Any type of violence | Ever | 422 (45%) | Client | HIV testing | OR: 1.05 (0.81–1.36); aOR: 0.98 (0.75–1.28) |
| Zhang (2012) | Any type of violence | Ever | 430 (58%) | Intimate partner | HIV testing | OR: 0.80 (0.59–1.07); aOR: 0.80 (0.59–1.09) |

### Appendix 15. Estimates of the association between other forms of violence and HIV outcomes

| **Author (year)** | **Violence definition** | **Violence time frame** | **Exposed, n (%)** | **Perpetrator** | **HIV outcome** | **Effect size (95% CI)** |
| --- | --- | --- | --- | --- | --- | --- |
| Alaei (2021) | Experience of violence, stigma or discrimination | Past 12 months | 1317 (55%) | Unspecified | HIV prevalence | OR: 0.61 (0.37–1.00) |
| Decker (2012) | Presented with more clients than agreed | Ever | 63 (44%) | Unspecified | HIV prevalence | OR: 0.95 (0.20–4.41) |
| Goldenberg (2016) | Abducted into the Lord's Resistance Army | Past 6 months | 129 (32%) | Army | HIV prevalence | OR: 1.61 (1.04–2.49); aOR: 1.62 (1.00–2.63) |
| Jennings Mayo-Wilson (2023) | Reported economic abuse (high vs. low) | Ever | 175 (32%) | Unspecified | ART initiation | OR: 0.26 (0.03–2.52); aOR: 0.21 (0.02–2.22) |
| Jennings Mayo-Wilson (2023) | Reported economic abuse (medium vs. low) | Ever | 178 (33%) | Unspecified | ART initiation | OR: 0.37 (0.03–4.20); aOR: 0.44 (0.03–5.78) |
| Jennings Mayo-Wilson (2023) | Reported economic abuse (high vs. low) | Ever | 175 (32%) | Unspecified | HIV testing | OR: 1.34 (0.61–2.93); aOR: 1.29 (0.58–2.90) |
| Jennings Mayo-Wilson (2023) | Reported economic abuse (medium vs. low) | Ever | 178 (33%) | Unspecified | HIV testing | OR: 1.66 (0.73–3.75); aOR: 1.74 (0.76–4.01) |
| MacLin (2023) | SW-related police harassment | Past 6 months | 26 (12%) | Police | Viral suppression | OR: 0.85 (0.33–2.15); aOR: 0.82 (0.32–2.15) |
| MacLin (2023) | SW-related police harassment | Past 6 months | 37 (37%) | Police | Viral suppression | OR: 0.81 (0.33–1.98); aOR: 0.79 (0.32–1.95) |
| MacLin (2023) | SW-related police harassment | Past 6 months | 26 (12%) | Police | ART adherence | OR: 0.50 (0.21–1.18); aOR: 0.46 (0.19–1.14) |
| MacLin (2023) | SW-related police harassment | Past 6 months | 26 (12%) | Police | ART adherence | OR: 0.78 (0.33–1.83); aOR: 0.71 (0.29–1.72) |
| MacLin (2023) | SW-related police harassment | Past 6 months | 37 (37%) | Police | ART adherence | OR: 1.18 (0.34–4.08); aOR: 1.39 (0.38–3.85) |
| MacLin (2023) | SW-related police harassment | Past 6 months | 37 (37%) | Police | ART adherence | OR: 2.66 (1.08–6.51); aOR: 2.78 (1.12–6.88) |
| MacLin (2023) | SW-related police harassment | Past 6 months | 26 (12%) | Police | Current ART | OR: 0.98 (0.11–8.33); aOR: 0.99 (0.11–8.59) |
| MacLin (2023) | SW-related police harassment | Past 6 months | 37 (37%) | Police | Current ART | OR: 2.06 (0.58–7.74); aOR: 2.05 (0.58–7.31) |
| Mukherjee (2022) | Extortion & Discrimination (vs. low victimisation) | Past 3 months | 39 (15%) | Police | HIV prevalence | OR: 0.67 (0.27–1.66); aOR: 0.66 (0.26–1.68) |
| Mukherjee (2022) | Poly-Victimization (vs. low victimisation) | Past 3 months | 86 (34%) | Police | HIV prevalence | OR: 1.26 (0.68–2.32); aOR: 1.26 (0.68–2.37) |
| Sileo (2018) | Any emotional violence | Ever | 77 (67%) | Unspecified | HIV prevalence | OR: 2.86 (0.95–8.64) |
| Sileo (2018) | Any emotional intimate partner violence | Past 12 months | 67 (58%) | Intimate partner | HIV prevalence | OR: 1.81 (0.70–4.65) |
| Tounkara (2014) | Psychological violence | Past month | 326 (34%) | Unspecified | HIV prevalence | OR: 1.51 (1.10–2.08); aOR: 1.41 (1.08–1.85) |
| West (2022) | Material violence, verbal and emotional harassment | Past 6 months | 279 (48%) | Police | HIV testing | OR: 2.42 (1.68–3.49) |
| Zhang (2012) | Belittled, humiliated or threatened | Ever | 372 (40%) | Client | HIV testing | OR: 1.12 (0.87–1.46) |
| Zhang (2012) | Belittled, humiliated or threatened | Ever | 411 (56%) | Intimate partner | HIV testing | OR: 0.77 (0.58–1.03) |

### Appendix 16. Study quality scores of included studies

| **Study** | **Domain 1: Sample representativeness** | **Domain 2: Temporality** | **Domain Three: Control for confounding** | **Total score /3** |
| --- | --- | --- | --- | --- |
| Abdella (2022) (1) | Random sampling | Cross-sectional | Adjusted | 2 |
| Aho (2013) (2) | Not random sampling | Cross-sectional | Unadjusted | 0 |
| Alaei (2021) (3) | Random sampling | Cross-sectional | Unadjusted | 1 |
| Alary (2014) (4) | Random sampling | Cross-sectional | Adjusted | 2 |
| Alemu (2022) (5) | Random sampling | Cross-sectional | Adjusted | 2 |
| Amogne (2019) (6) | Random sampling | Cross-sectional | Unadjusted | 1 |
| Arimide (2022) (7) | Random sampling | Cross-sectional | Adjusted | 2 |
| Arumugam (2022) (8) | Random sampling | Cross-sectional | Unadjusted | 1 |
| Ayamah (2023) (9) | Random sampling | Cross-sectional | Unadjusted | 1 |
| Beattie (2010) (10) | Random sampling | Cross-sectional | Adjusted | 2 |
| Beattie (2015) (11) | Random sampling | Cross-sectional | Adjusted | 2 |
| Beksinska (2018) (12) | Random sampling | Cross-sectional | Adjusted | 2 |
| Berger (2018) (13) | Random sampling | Cross-sectional | Unadjusted | 1 |
| Bhardwaj (2023) (14) | Not random sampling | Cross-sectional | Adjusted | 1 |
| Bowring (2019) (15) | Random sampling | Cross-sectional | Unadjusted | 1 |
| Braunstein (2011) (16) | Not random sampling | Cross-sectional | Adjusted | 1 |
| Budhwani (2017) (17) | Not random sampling | Cross-sectional | Unadjusted | 0 |
| Budhwani (2021) (18) | Not random sampling | Cross-sectional | Adjusted | 1 |
| Bugssa (2015) (19) | Random sampling | Cross-sectional | Adjusted | 2 |
| Chabata (2019) (20) | Random sampling | Cross-sectional | Unadjusted | 1 |
| Coetzee (2017) (21) | Random sampling | Cross-sectional | Unadjusted | 1 |
| Comins (2024) (22) | Not random sampling | Cross-sectional | Adjusted | 1 |
| Damacena (2011) (23) | Random sampling | Cross-sectional | Unadjusted | 1 |
| Davey (2020) (24) | Random sampling | Cross-sectional | Unadjusted | 1 |
| Decker (2012) (25) | Random sampling | Cross-sectional | Unadjusted | 1 |
| Decker (2014) (26) | Random sampling | Cross-sectional | Adjusted | 2 |
| Decker (2016) (27) | Random sampling | Cross-sectional | Adjusted | 2 |
| Doshi (2018) (28) | Random sampling | Cross-sectional | Unadjusted | 1 |
| Duff (2016) (29) | Random sampling | Longitudinal | Unadjusted | 2 |
| Emmanuel (2021) (30) | Random sampling | Cross-sectional | Unadjusted | 1 |
| Faini (2022) (31) | Random sampling | Cross-sectional | Unadjusted | 1 |
| Goldenberg (2013) (32) | Random sampling | Cross-sectional | Unadjusted | 1 |
| Goldenberg (2016a) (33) | Random sampling | Cross-sectional | Unadjusted | 1 |
| Goldenberg (2016b) (34) | Random sampling | Longitudinal | Unadjusted | 2 |
| Goldenberg (2019) (35) | Random sampling | Cross-sectional | Unadjusted | 1 |
| Guure (2023) (36) | Random sampling | Cross-sectional | Adjusted | 2 |
| Hendrickson (2018) (37) | Random sampling | Cross-sectional | Adjusted | 2 |
| Hensen (2019) (38) | Random sampling | Cross-sectional | Adjusted | 2 |
| Hladik (2017) (39) | Random sampling | Cross-sectional | Unadjusted | 1 |
| Iakunchykova (2017) (40) | Random sampling | Cross-sectional | Adjusted | 2 |
| Izadi (2023) (41) | Random sampling | Cross-sectional | Unadjusted | 1 |
| Jennings Mayo-Wilson (2023) (42) | Random sampling | Cross-sectional | Adjusted | 2 |
| Jones (2023) (43) | Not random sampling | Longitudinal | Adjusted | 2 |
| Kakisingi (2020) (44) | Not random sampling | Cross-sectional | Adjusted | 1 |
| Kassanjee (2022) (45) | Not random sampling | Cross-sectional | Unadjusted | 0 |
| Kelly-Hanku (2020) (46) | Random sampling | Cross-sectional | Adjusted | 2 |
| Kerrigan (2017) (47) | Random sampling | Cross-sectional | Unadjusted | 1 |
| Kumar (2014) (48) | Random sampling | Cross-sectional | Adjusted | 2 |
| Leddy (2018) (49) | Random sampling | Cross-sectional | Unadjusted | 1 |
| Lima (2017) (50) | Random sampling | Cross-sectional | Unadjusted | 1 |
| Lyons (2017) (51) | Random sampling | Cross-sectional | Unadjusted | 1 |
| Lyons (2020) (52) | Random sampling | Cross-sectional | Adjusted | 2 |
| MacLin (2023) (53) | Not random sampling | Cross-sectional | Adjusted | 1 |
| Mendoza (2017) (54) | Not random sampling | Cross-sectional | Adjusted | 1 |
| Mizinduko (2020) (55) | Random sampling | Cross-sectional | Adjusted | 2 |
| Mountain (2018) (56) | Random sampling | Cross-sectional | Adjusted | 2 |
| Mukherjee (2022) (57) | Not random sampling | Cross-sectional | Adjusted | 1 |
| Mukuku (2024) (58) | Random sampling | Cross-sectional | Adjusted | 2 |
| Mulholland (2022) (59) | Random sampling | Cross-sectional | Unadjusted | 1 |
| Mutagoma (2017) (60) | Random sampling | Cross-sectional | Unadjusted | 1 |
| Mutagoma (2019) (61) | Random sampling | Cross-sectional | Unadjusted | 1 |
| Nabayinda (2023) (62) | Random sampling | Cross-sectional | Unadjusted | 1 |
| Namale (2019) (63) | Not random sampling | Cross-sectional | Unadjusted | 0 |
| Oldenburg (2018) (64) | Random sampling | Cross-sectional | Adjusted | 2 |
| Ouma (2021) (65) | Not random sampling | Cross-sectional | Adjusted | 1 |
| Pando (2013) (66) | Not random sampling | Cross-sectional | Adjusted | 1 |
| Prangnell (2018) (67) | Not random sampling | Cross-sectional | Unadjusted | 0 |
| Ramesh (2012) (68) | Random sampling | Cross-sectional | Adjusted | 2 |
| Rice (2022) (69) | Random sampling | Cross-sectional | Unadjusted | 1 |
| Schwartz (2016) (70) | Random sampling | Cross-sectional | Adjusted | 2 |
| Sherman (2019) (71) | Random sampling | Cross-sectional | Unadjusted | 1 |
| Shokoohi (2017) (72) | Not random sampling | Cross-sectional | Adjusted | 1 |
| Shrestha (2017) (73) | Random sampling | Cross-sectional | Adjusted | 2 |
| Sileo (2018) (74) | Random sampling | Cross-sectional | Unadjusted | 1 |
| Strathdee (2011) (75) | Random sampling | Cross-sectional | Unadjusted | 1 |
| Szwarcwald (2017) (76) | Random sampling | Cross-sectional | Unadjusted | 1 |
| Tokar (2013) (77) | Random sampling | Cross-sectional | Unadjusted | 1 |
| Tounkara (2014) (78) | Random sampling | Cross-sectional | Adjusted | 2 |
| Tuot (2020) (79) | Random sampling | Cross-sectional | Unadjusted | 1 |
| Ulibarri (2011) (80) | Random sampling | Cross-sectional | Adjusted | 2 |
| Vandenhoudt (2013) (81) | Random sampling | Cross-sectional | Adjusted | 2 |
| Vu (2017) (82) | Random sampling | Cross-sectional | Unadjusted | 1 |
| Wand (2015) (83) | Random sampling | Cross-sectional | Adjusted | 2 |
| Wang (2021) (84) | Random sampling | Cross-sectional | Unadjusted | 1 |
| West (2022) (85) | Not random sampling | Cross-sectional | Unadjusted | 0 |
| Wilson (2016a) (86) | Not random sampling | Longitudinal | Adjusted | 2 |
| Wilson (2016b) (87) | Not random sampling | Cross-sectional | Unadjusted | 0 |
| Wirth (2013) (88) | Not random sampling | Cross-sectional | Adjusted | 1 |
| Xie (2023) (89) | Not random sampling | Cross-sectional | Unadjusted | 0 |
| Zalla (2019) (90) | Random sampling | Cross-sectional | Unadjusted | 1 |
| Zhang (2012) (91) | Not random sampling | Cross-sectional | Adjusted | 1 |

### Appendix 17. Forest plots of all forms of violence estimates and prevalent HIV infection

#### Pooled unadjusted estimates of recent exposure to all forms of violence and prevalent HIV infection among female sex workers

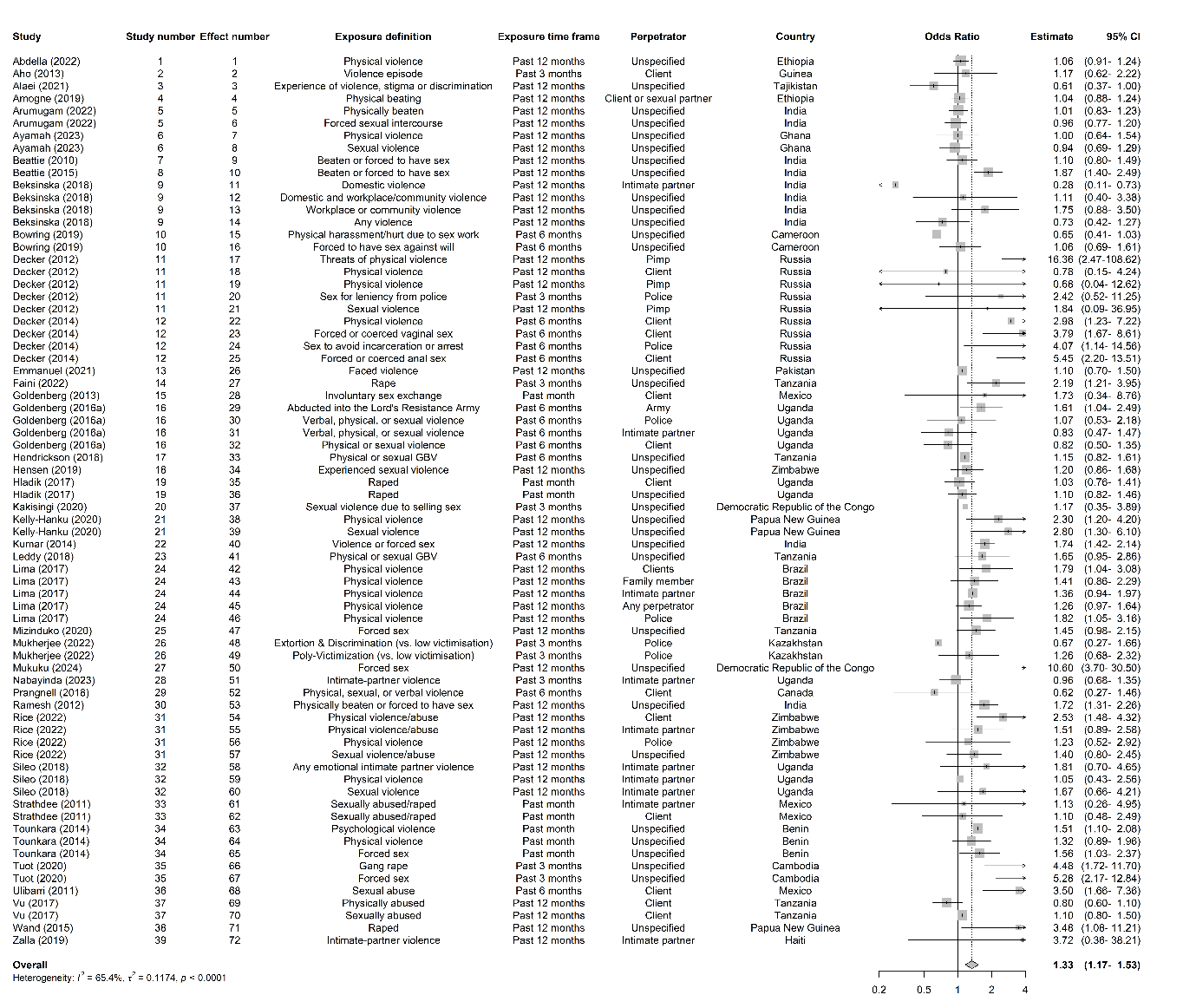

#### Pooled unadjusted estimates of lifetime exposure to all forms of violence and prevalent HIV infection among female sex workers

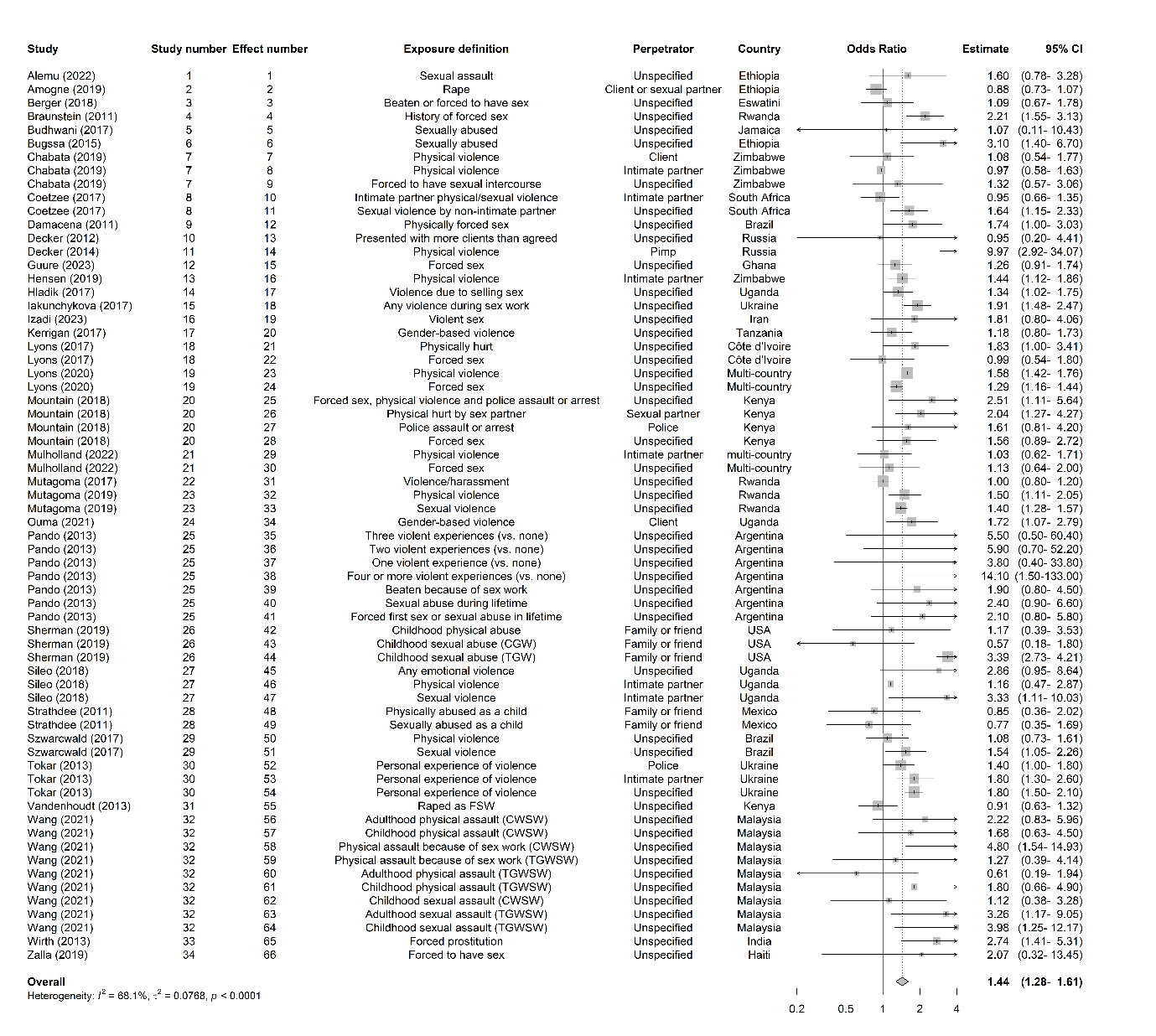

#### Pooled adjusted estimates of recent exposure to all forms of violence and prevalent HIV infection among female sex workers

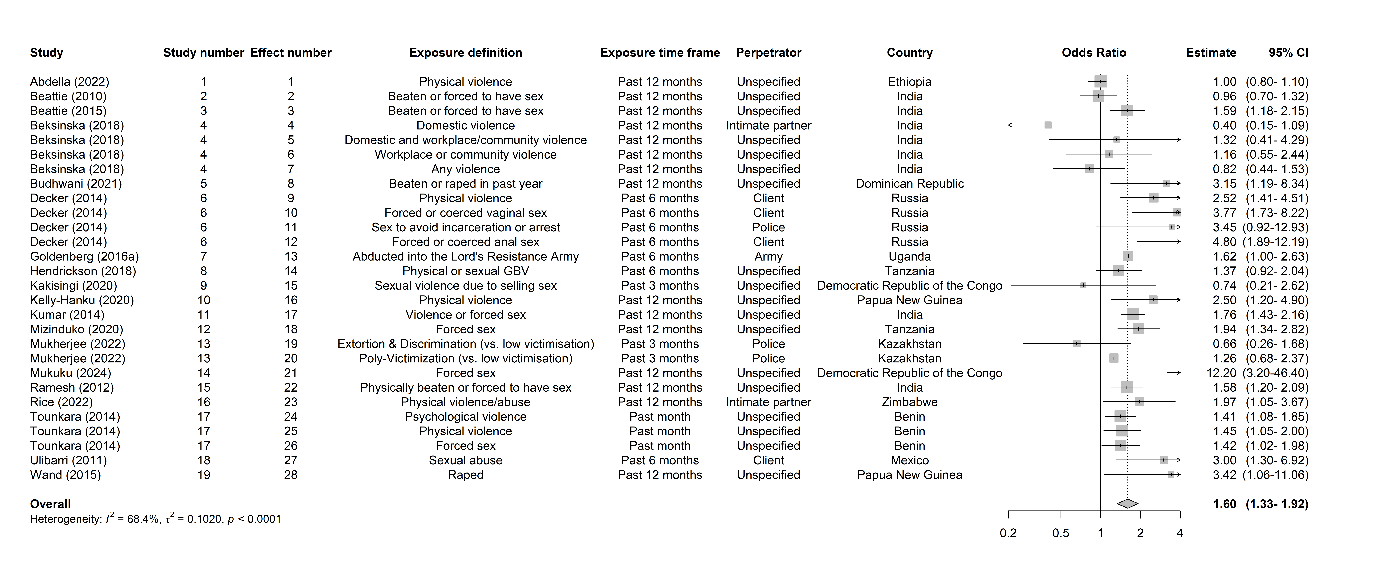

#### Pooled adjusted estimates of lifetime exposure to all forms of violence and prevalent HIV infection among female sex workers

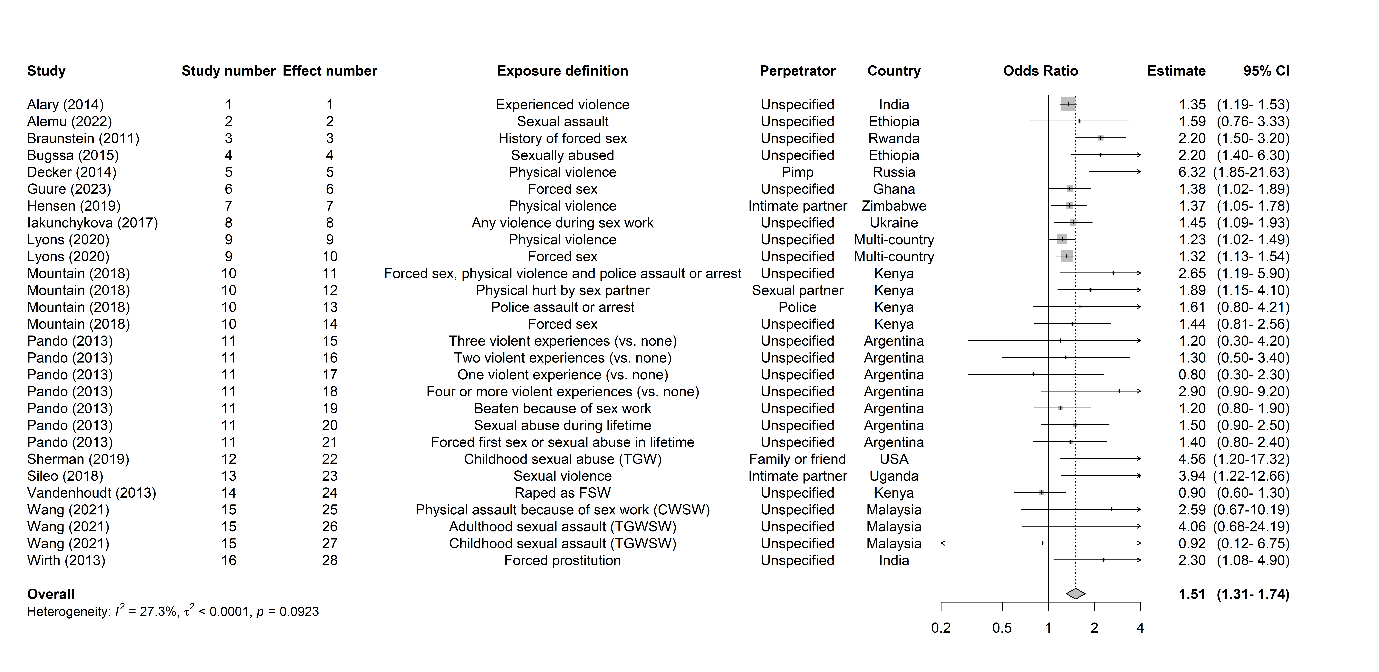

#### Pooled combined unadjusted and adjusted estimates of recent exposure to all forms of violence and prevalent HIV infection among female sex workers

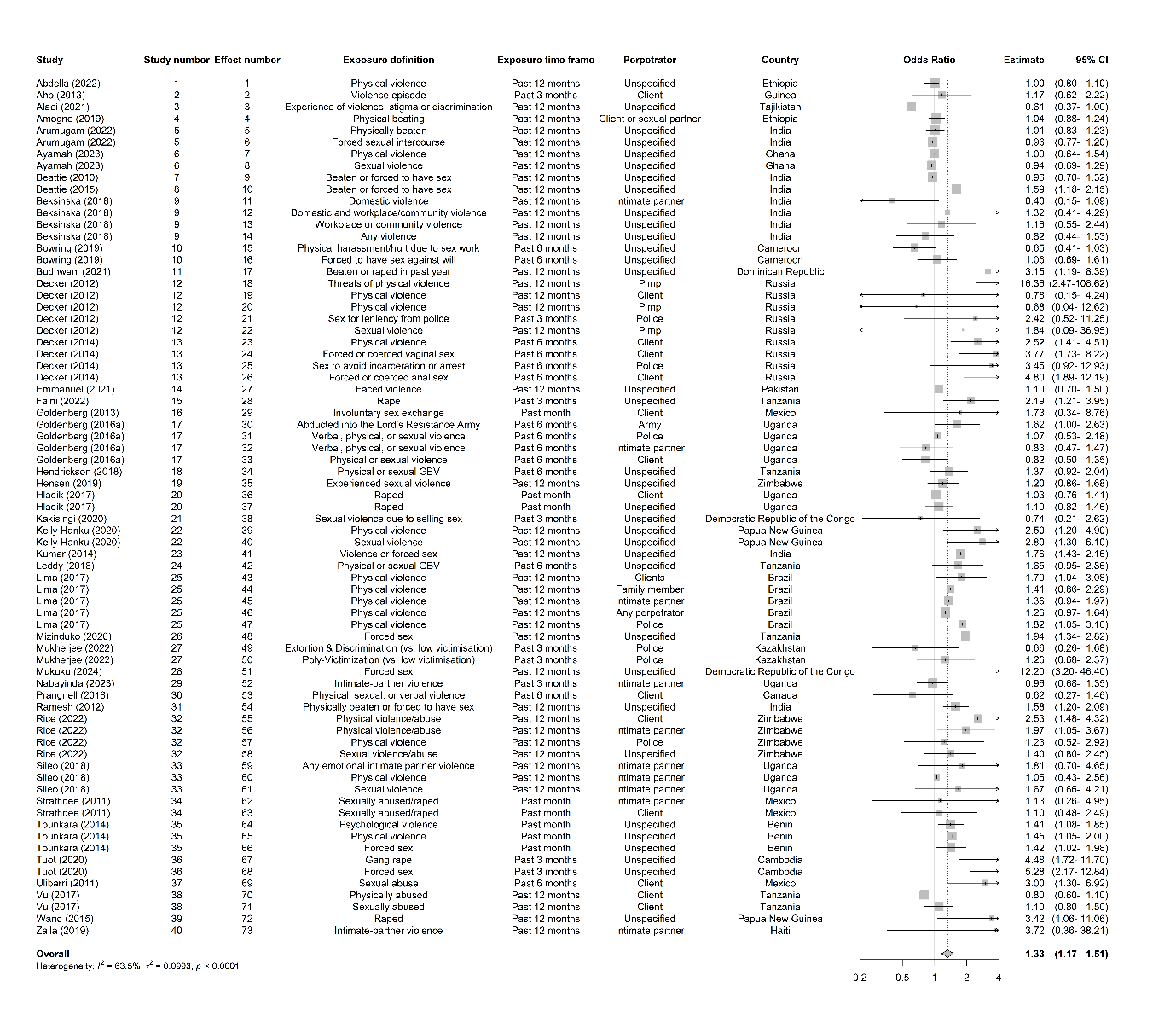

#### Pooled combined unadjusted and adjusted estimates of lifetime exposure to all forms of violence and prevalent HIV infection among female sex workers

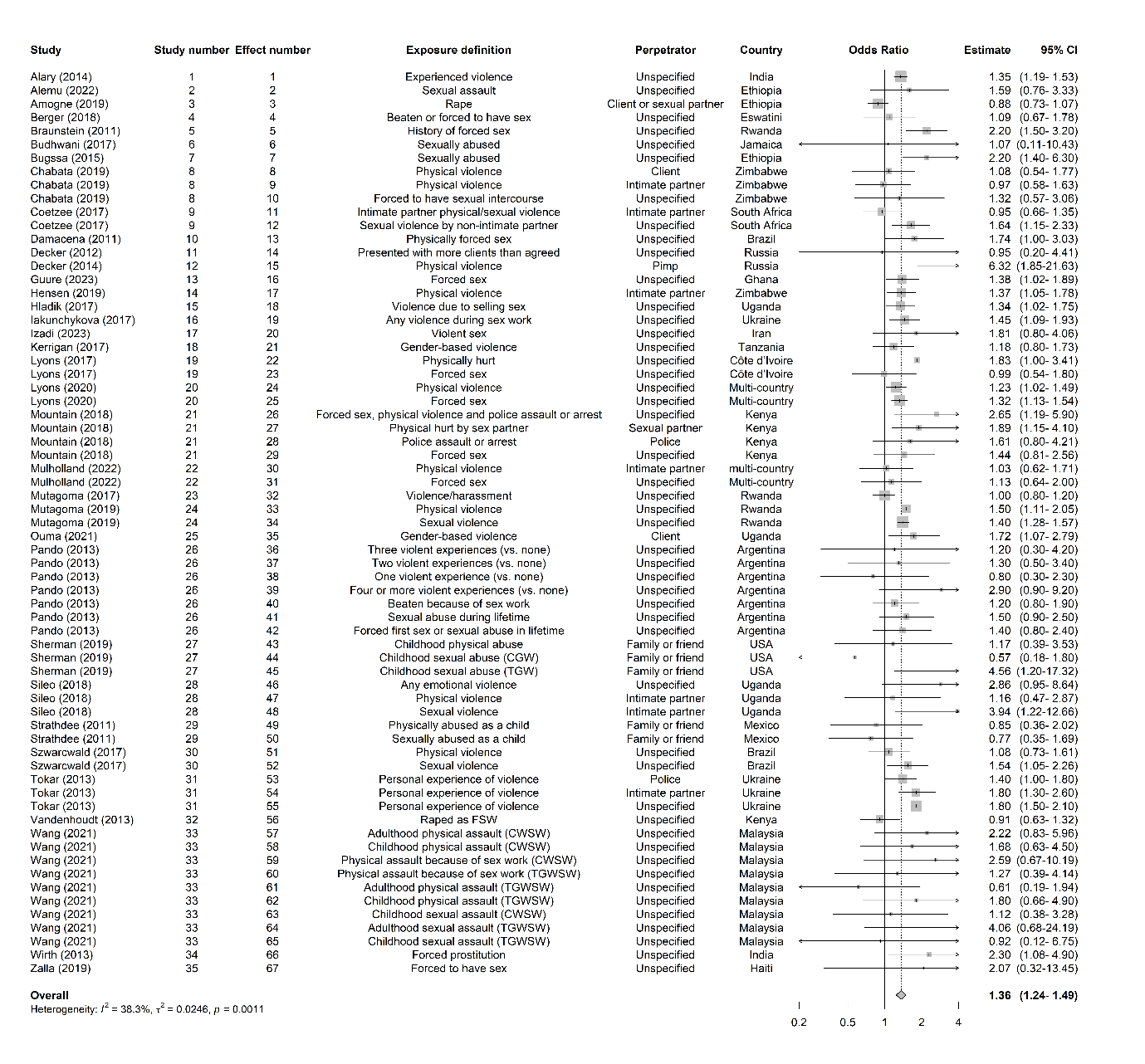

### Appendix 18. Forest plots of physical violence estimates and prevalent HIV infection

#### Pooled unadjusted estimates of recent exposure to physical violence and prevalent HIV infection among female sex workers

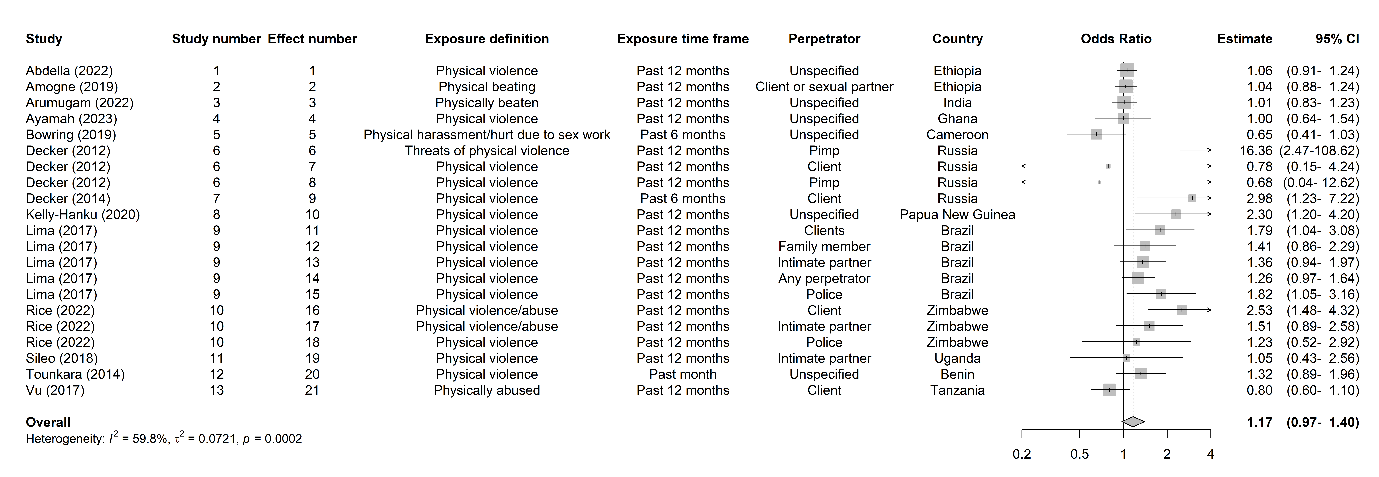

#### Pooled unadjusted estimates of lifetime exposure to physical violence and prevalent HIV infection among female sex workers

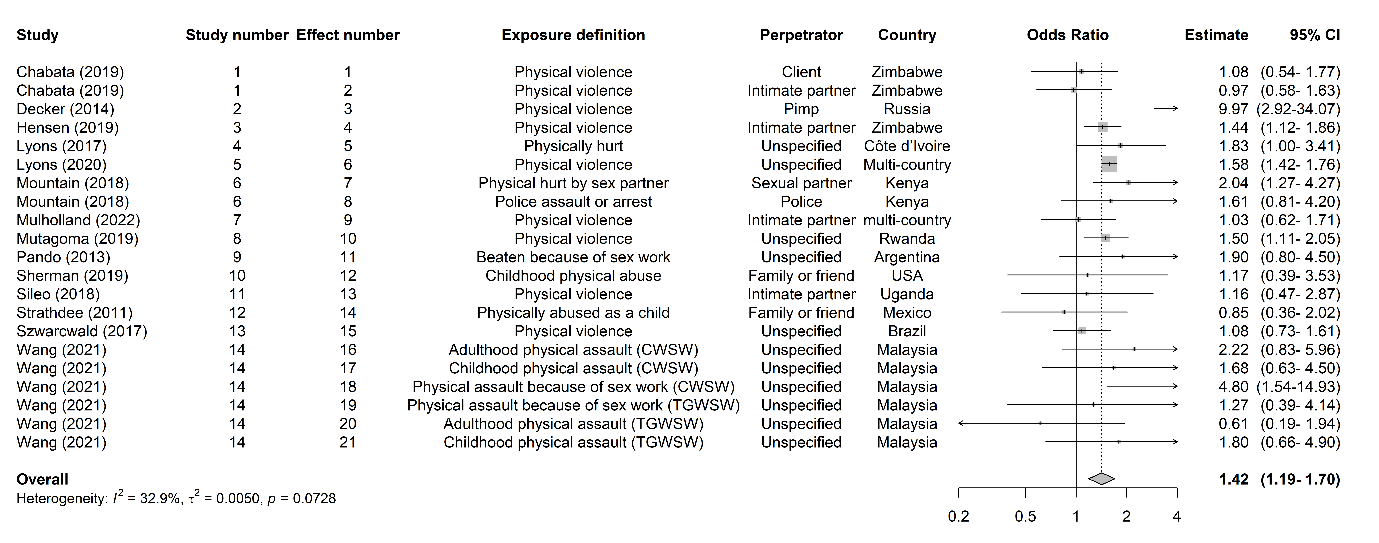

**Pooled adjusted estimates of recent exposure to physical violence and prevalent HIV infection among female sex workers**

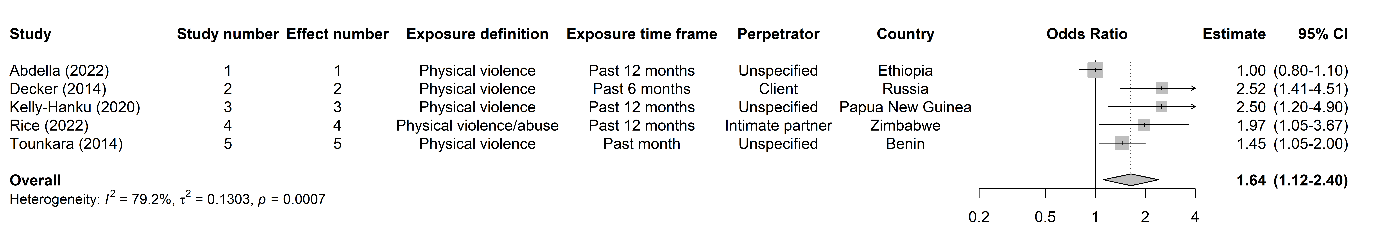

#### Pooled adjusted estimates of lifetime exposure to physical violence and prevalent HIV infection among female sex workers

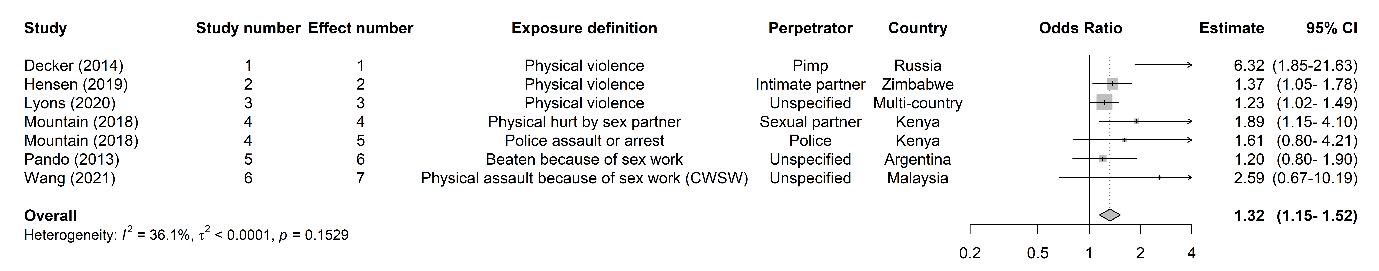

#### Pooled combined unadjusted and adjusted estimates of recent exposure to physical violence and prevalent HIV infection among female sex workers

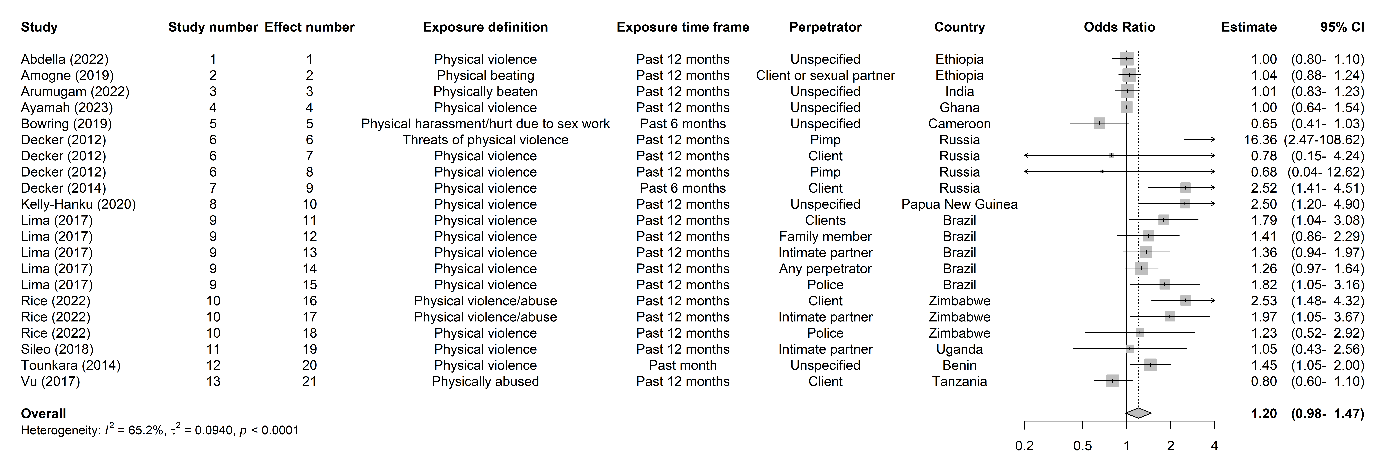

#### Pooled combined unadjusted and adjusted estimates of lifetime exposure to physical violence and prevalent HIV infection among female sex workers

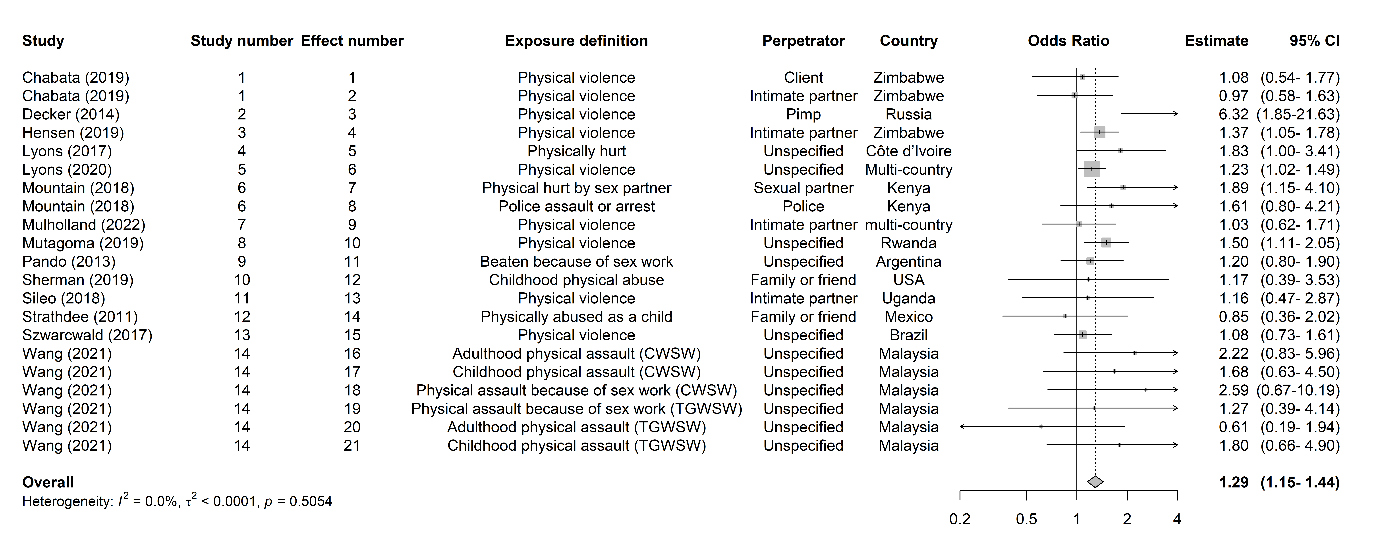

### Appendix 19. Forest plots of sexual violence estimates and prevalent HIV infection

#### Pooled unadjusted estimates of recent exposure to sexual violence and prevalent HIV infection among female sex workers

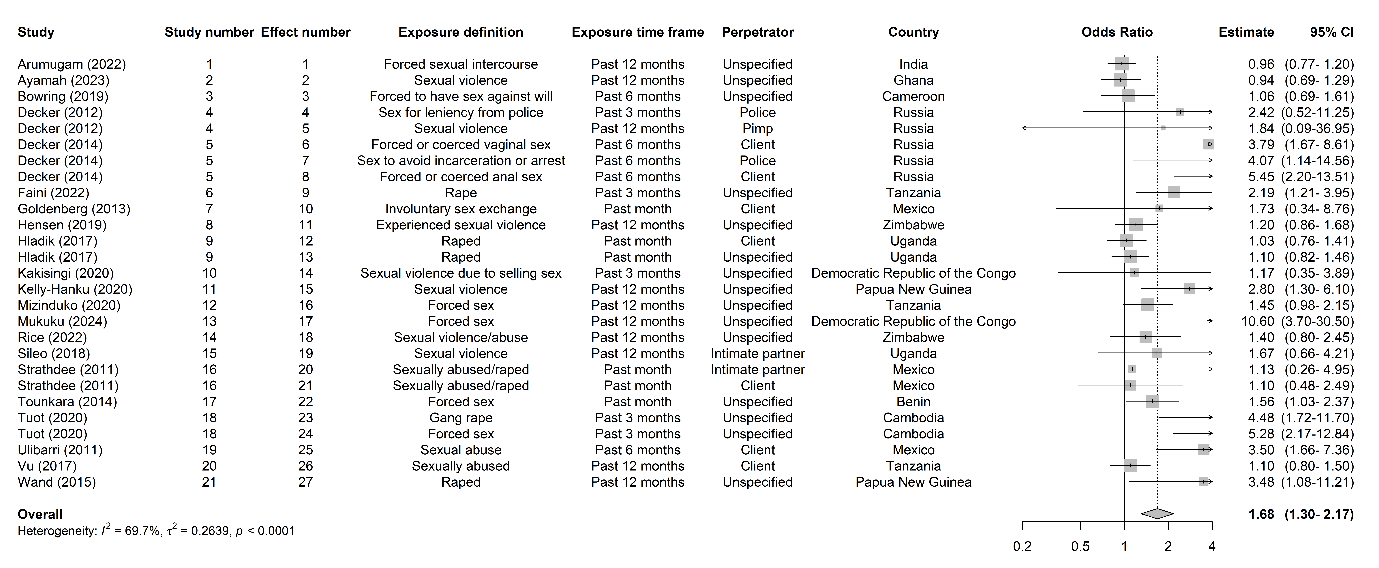

#### Pooled unadjusted estimates of lifetime exposure to sexual violence and prevalent HIV infection among female sex workers

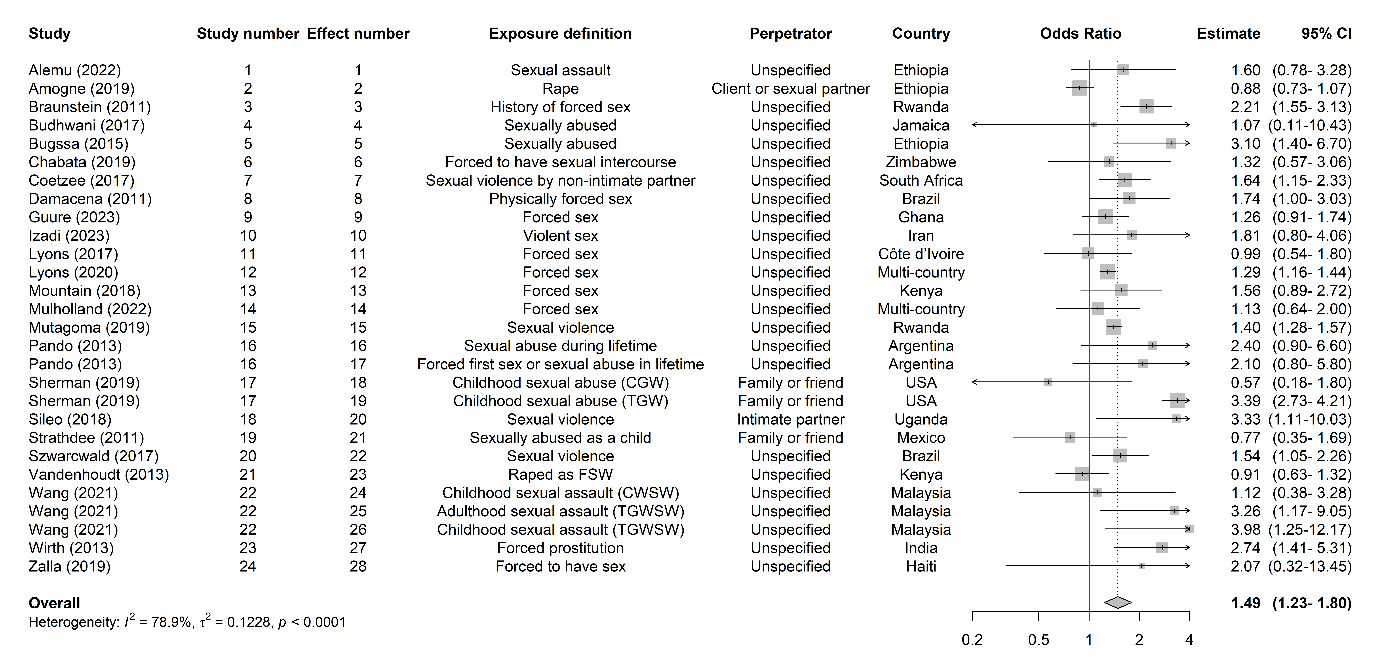

#### Pooled adjusted estimates of recent exposure to sexual violence and prevalent HIV infection among female sex workers

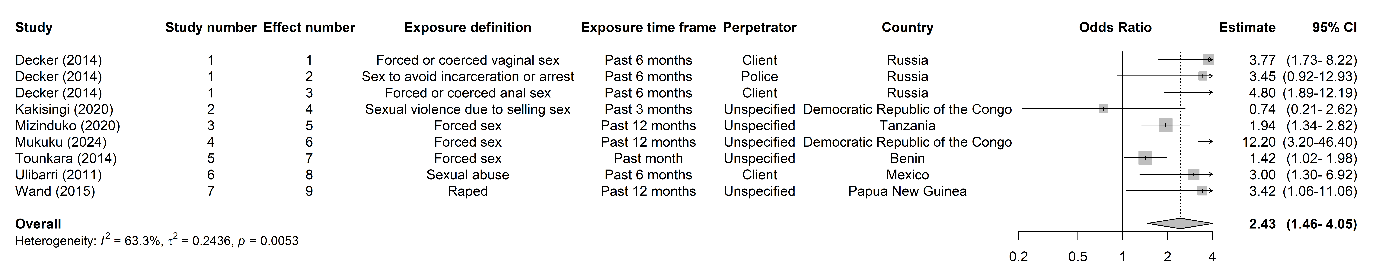

#### Pooled adjusted estimates of lifetime exposure to sexual violence and prevalent HIV infection among female sex workers

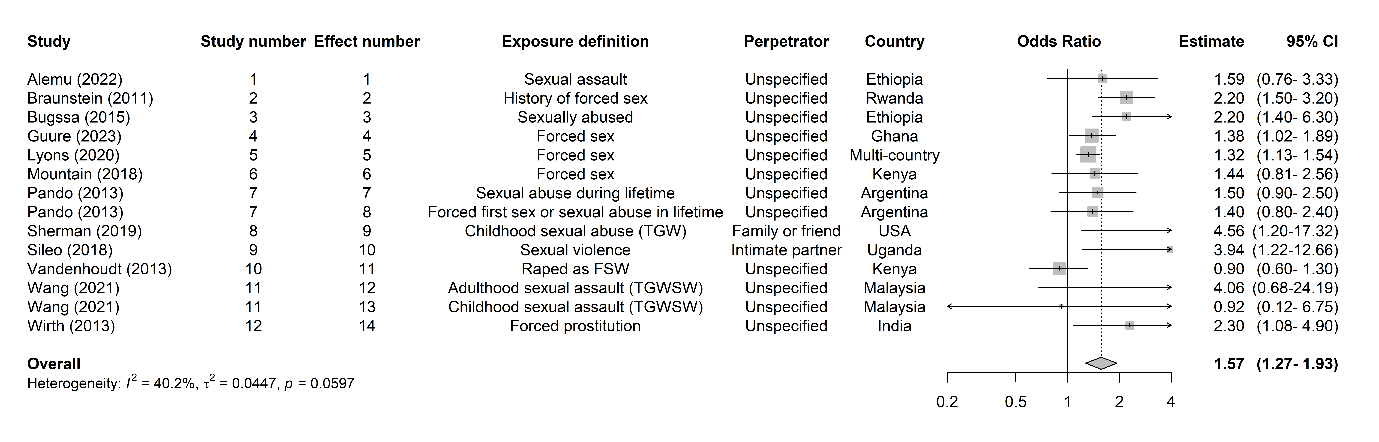

#### Pooled combined unadjusted and adjusted estimates of recent exposure to sexual violence and prevalent HIV infection among female sex workers

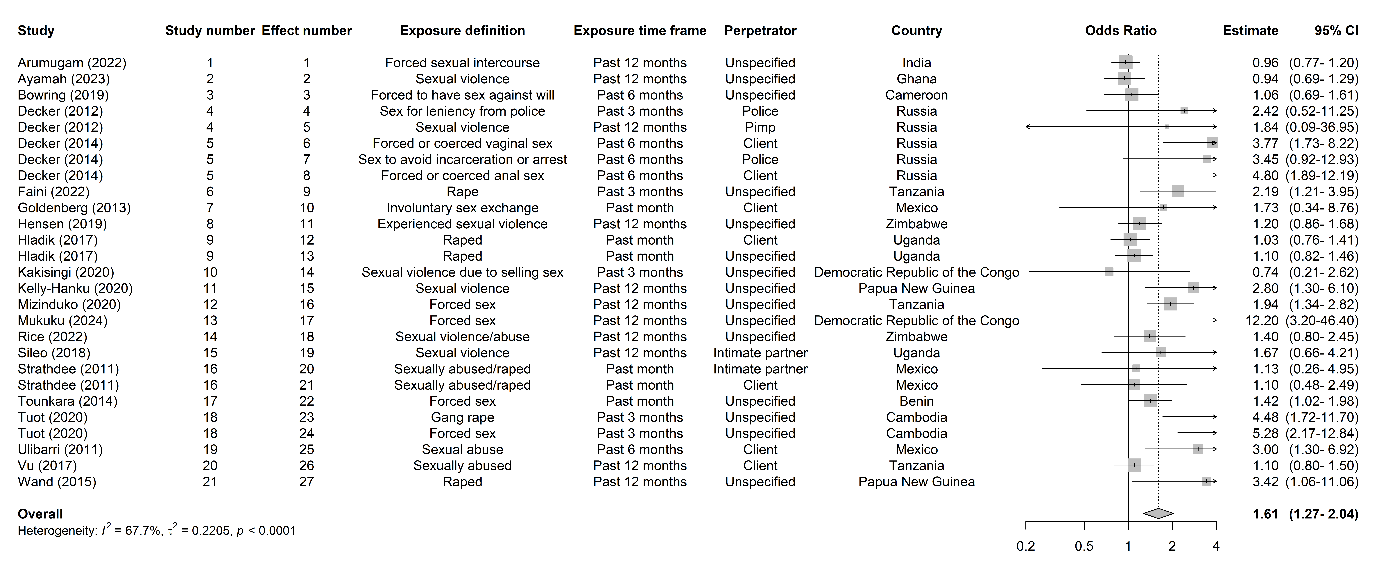

#### Pooled combined unadjusted and adjusted estimates of lifetime exposure to sexual violence and prevalent HIV infection among female sex workers

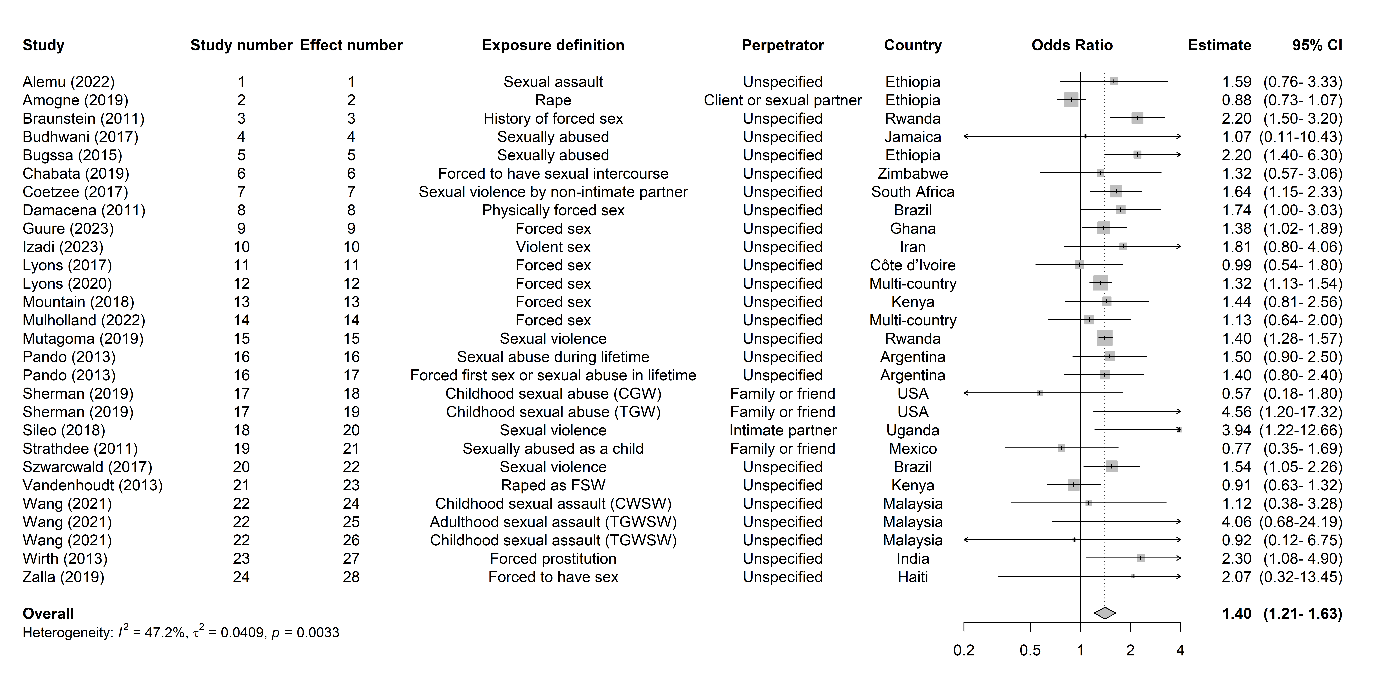

### Appendix 20. Forest plots of mixed violence estimates and prevalent HIV infection

#### Pooled unadjusted estimates of recent exposure to mixed violence and prevalent HIV infection among female sex workers

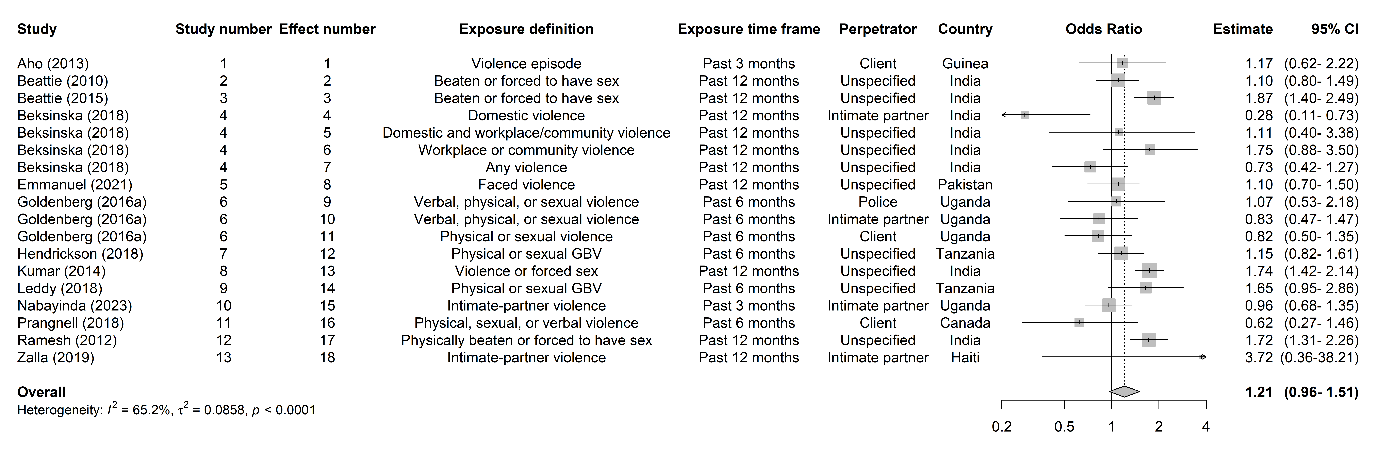

#### Pooled unadjusted estimates of lifetime exposure to mixed violence and prevalent HIV infection among female sex workers

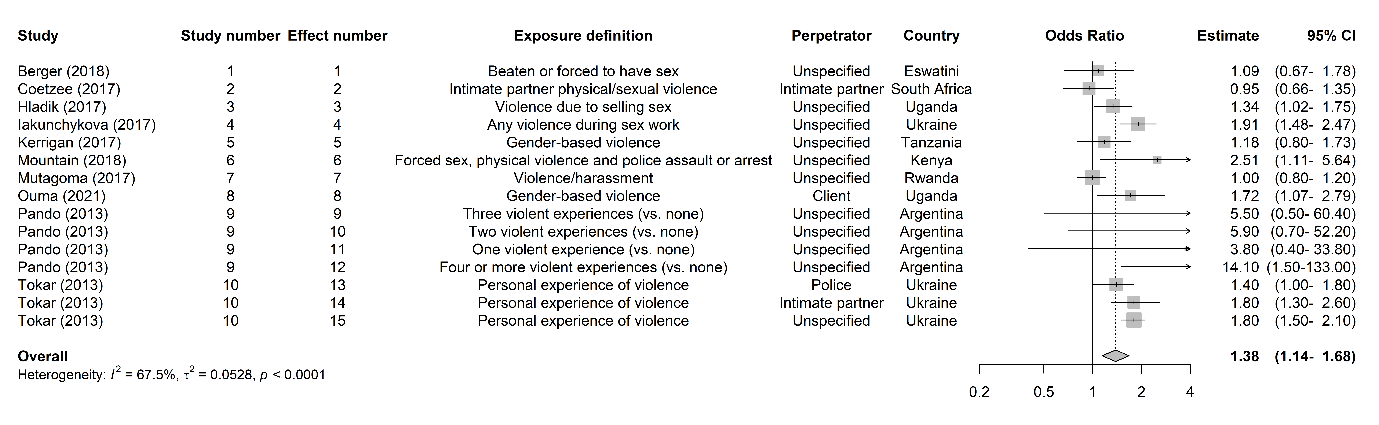

#### Pooled adjusted estimates of recent exposure to mixed violence and prevalent HIV infection among female sex workers

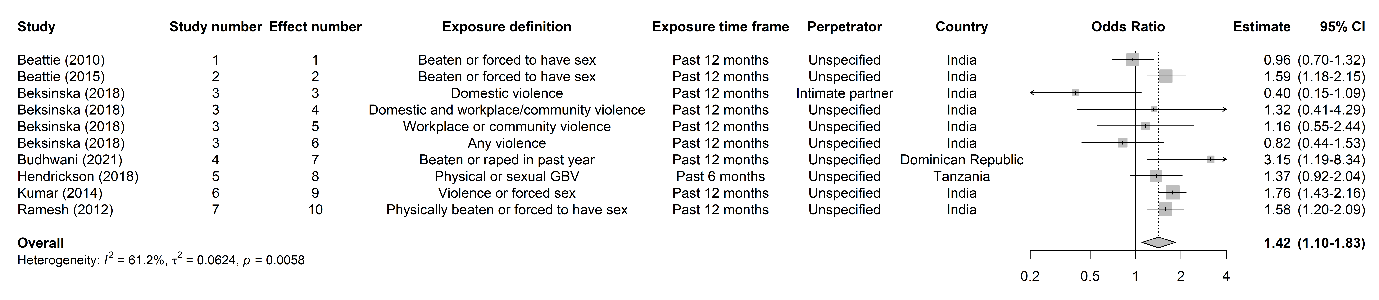

#### Pooled adjusted estimates of lifetime exposure to mixed violence and prevalent HIV infection among female sex workers

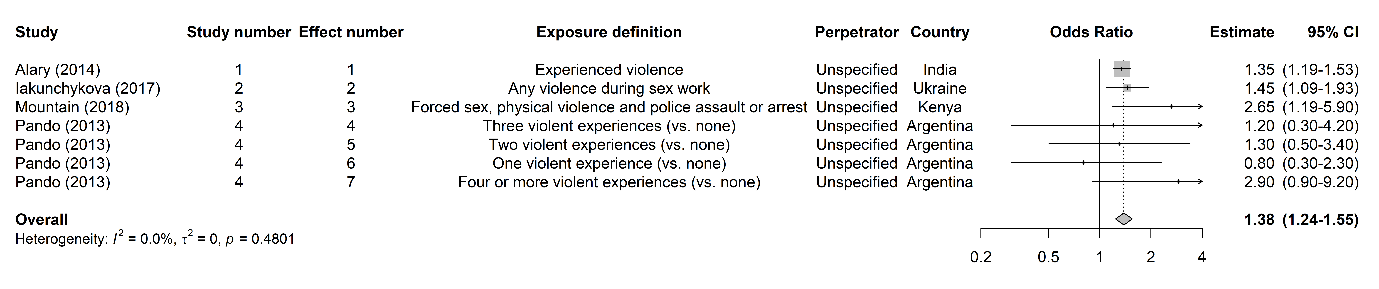

#### Pooled combined unadjusted and adjusted estimates of recent exposure to mixed violence and prevalent HIV infection among female sex workers

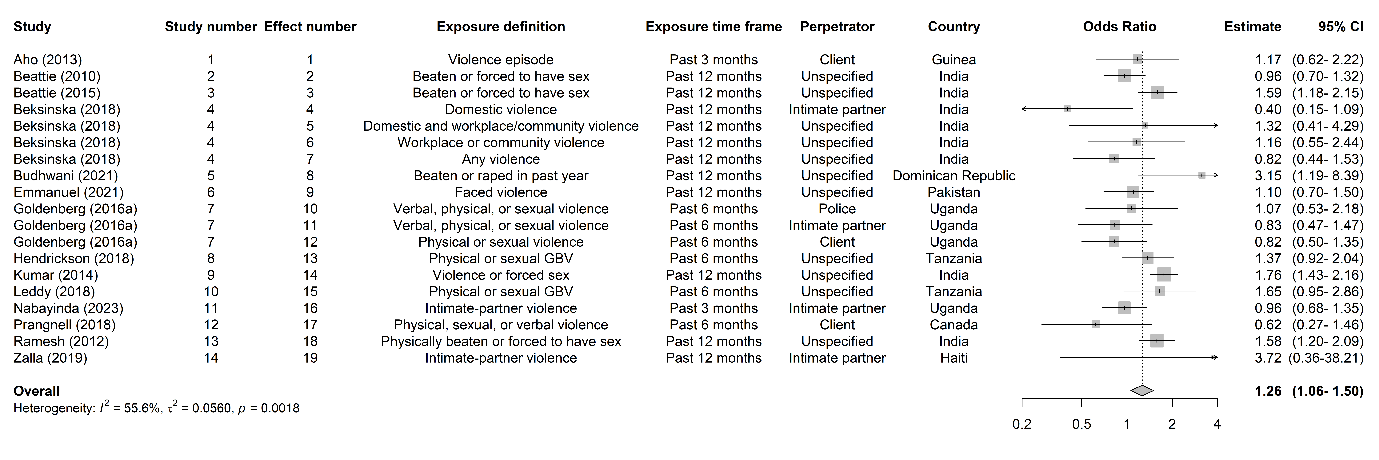

#### Pooled combined unadjusted and adjusted estimates of lifetime exposure to mixed violence and prevalent HIV infection among female sex workers

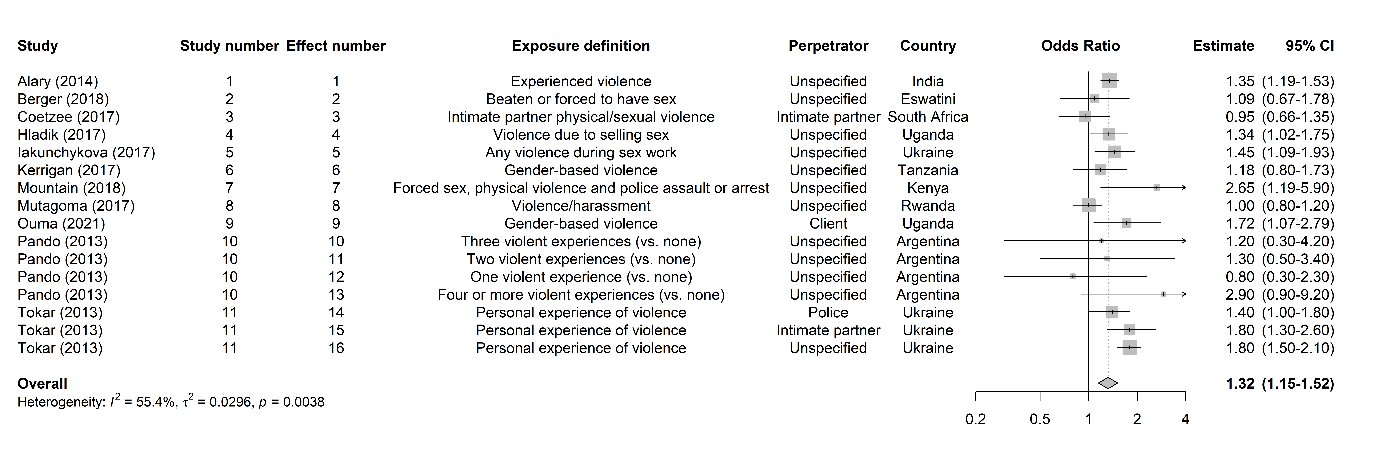

### Appendix 21. Forest plots of other forms of violence estimates and prevalent HIV infection

#### Pooled unadjusted estimates of recent exposure to other forms of violence and prevalent HIV infection among female sex workers

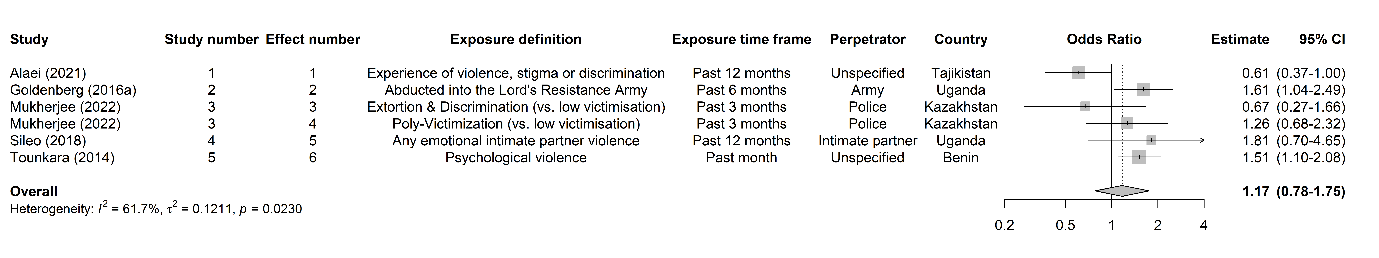

#### Pooled adjusted estimates of recent exposure to other forms of violence and prevalent HIV infection among female sex workers

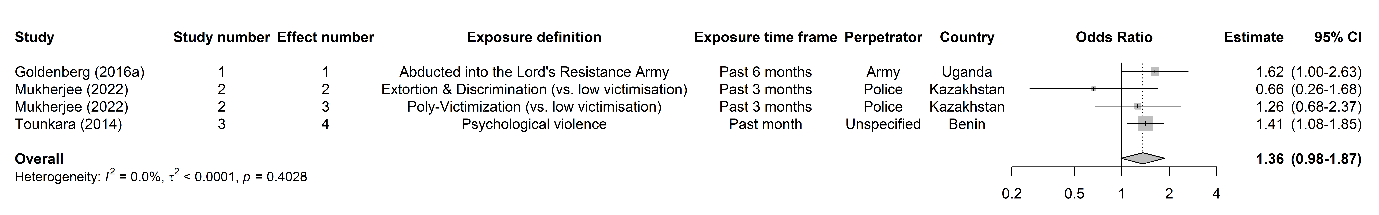

#### Pooled combined unadjusted and adjusted estimates of recent exposure to other forms of violence and prevalent HIV infection among female sex workers

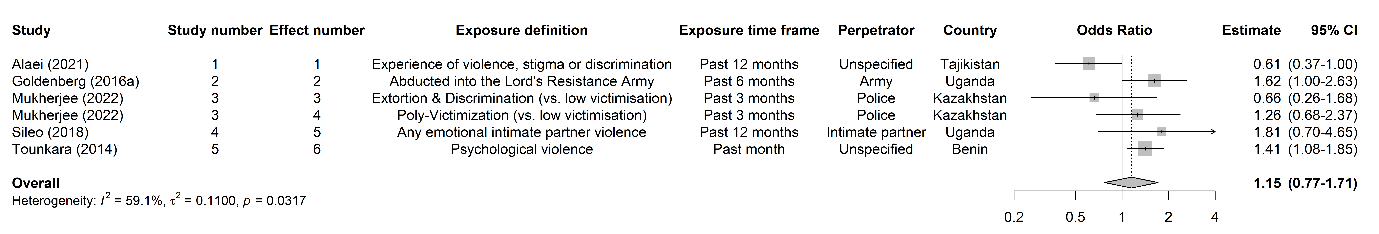

### Appendix 23. Forest plots of violence and HIV testing

#### Pooled unadjusted estimates of recent exposure to all forms of violence and HIV testing among female sex workers

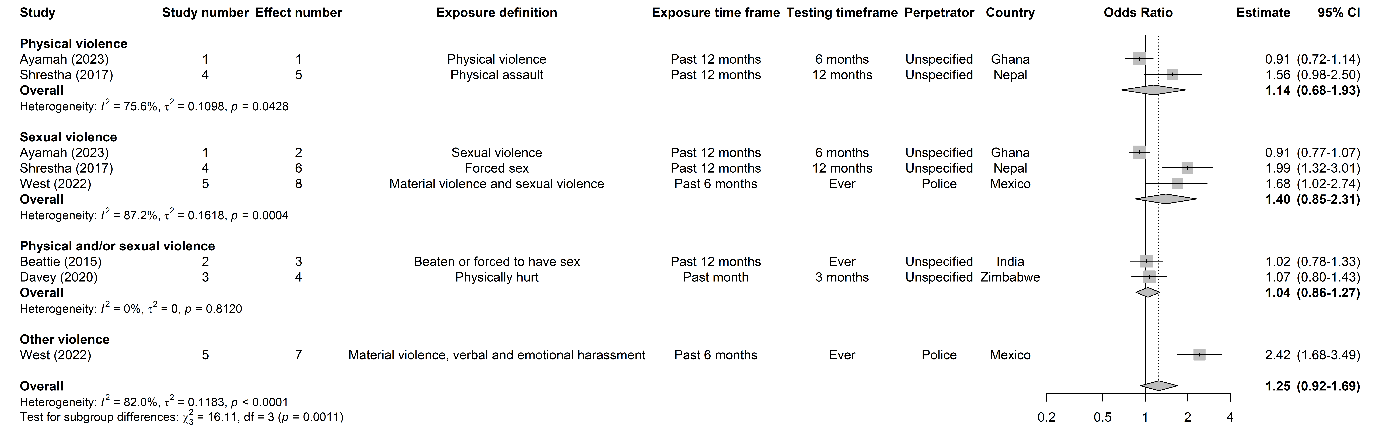

#### Pooled unadjusted estimates of lifetime exposure to all forms of violence and HIV testing among female sex workers

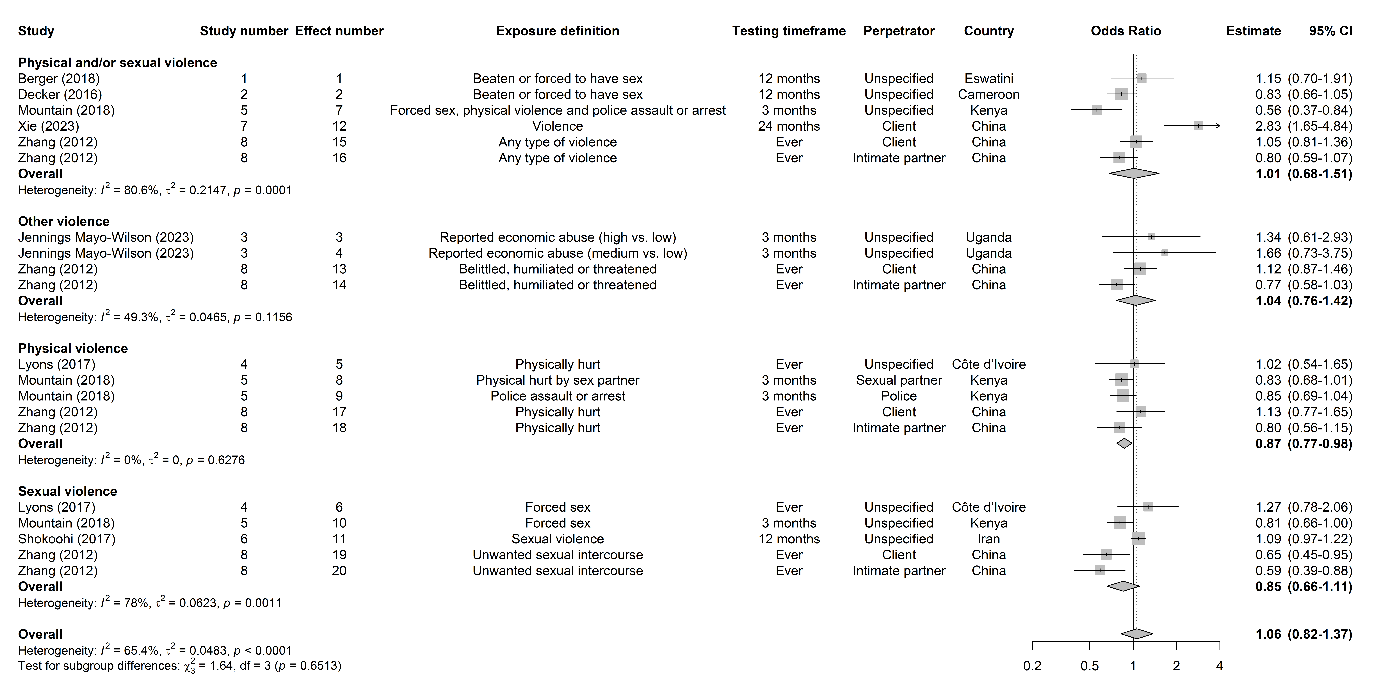

#### Pooled adjusted estimates of recent exposure to all forms of violence and HIV testing among female sex workers

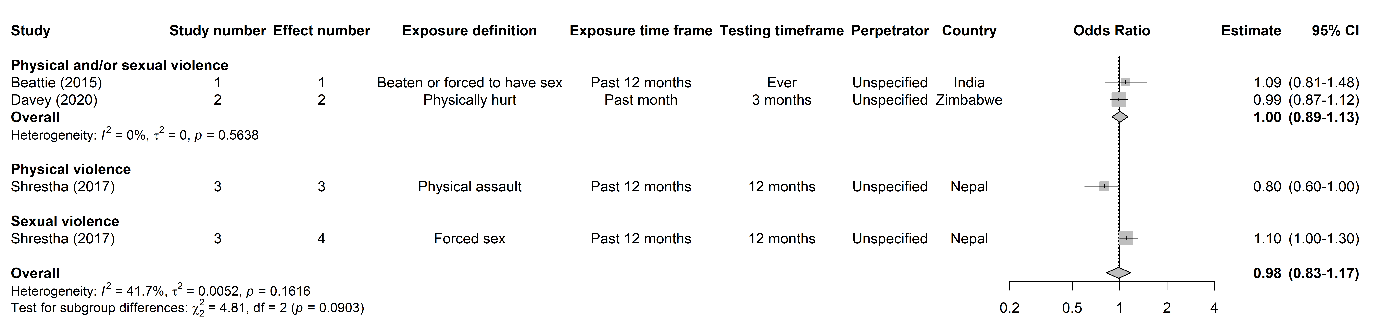

#### Pooled adjusted estimates of lifetime exposure to all forms of violence and HIV testing among female sex workers

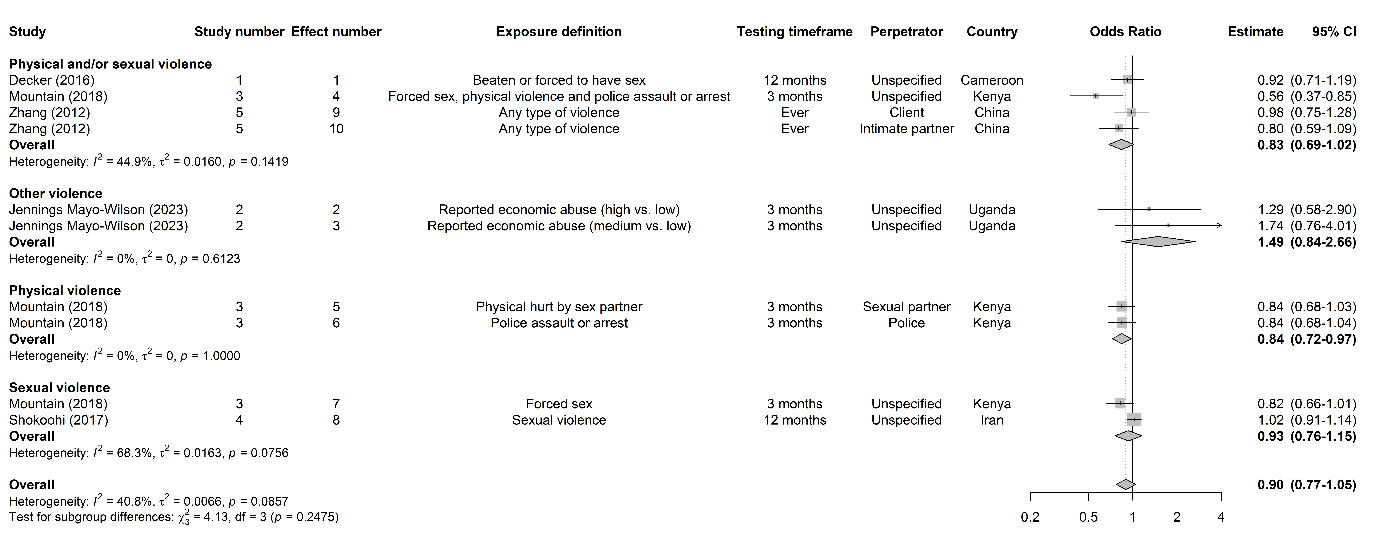

#### Pooled combined unadjusted and adjusted estimates of recent exposure to all forms of violence and HIV testing among female sex workers

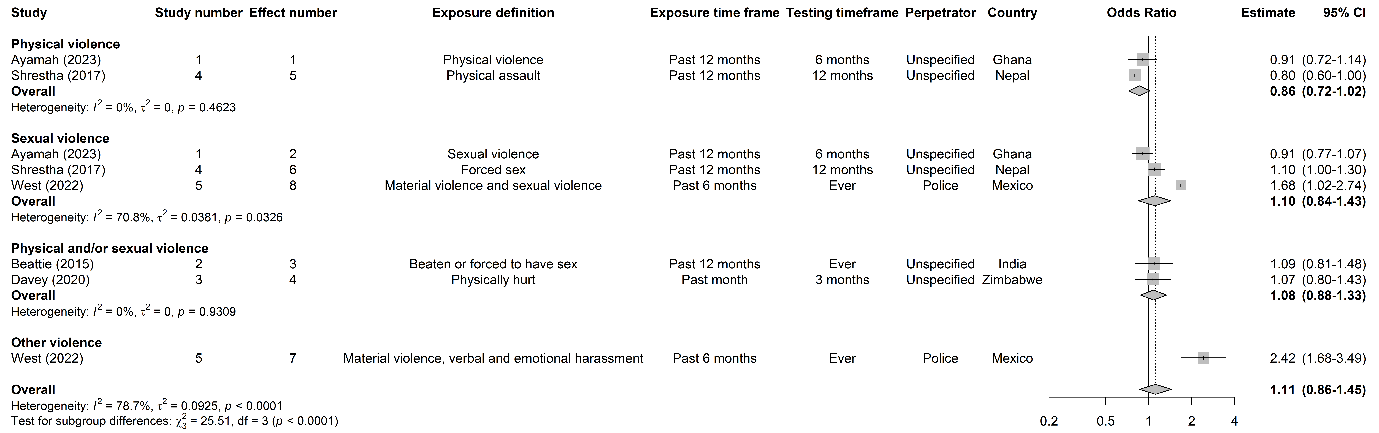

#### Pooled combined unadjusted and adjusted estimates of lifetime exposure to all forms of violence and HIV testing among female sex workers

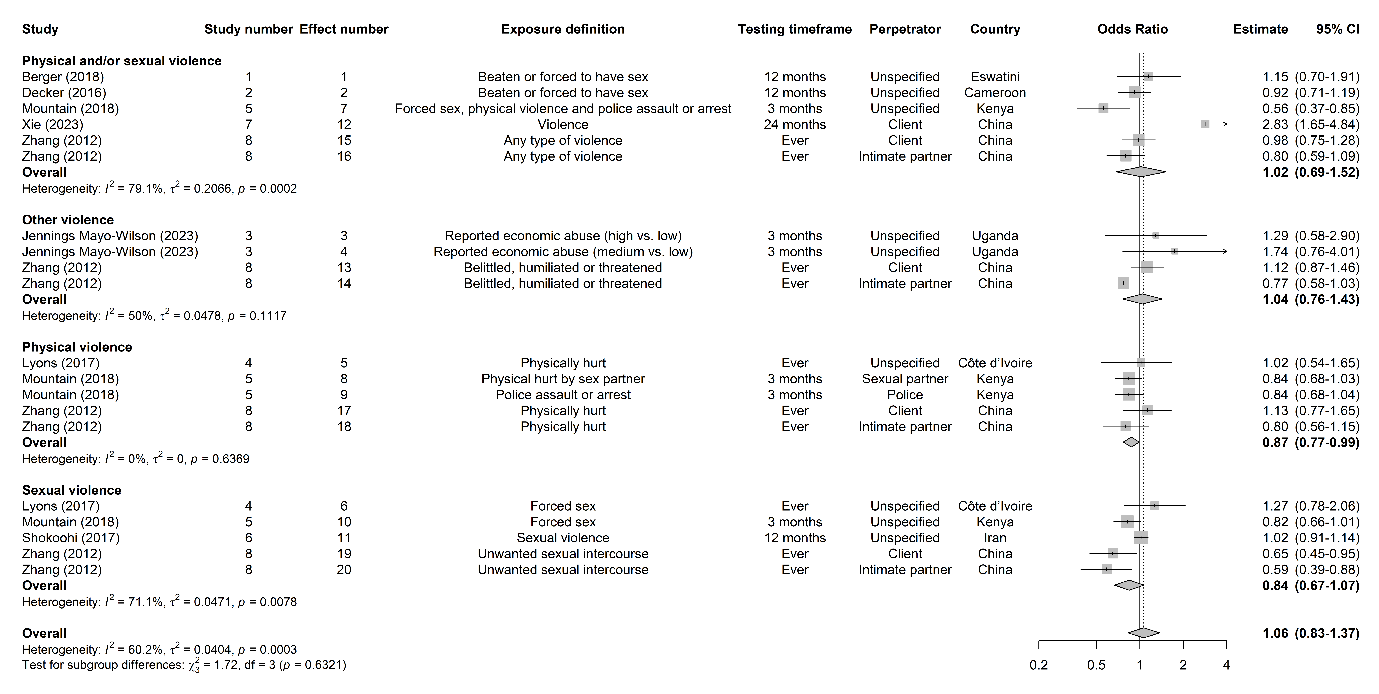

### Appendix 24. Forest plots of violence and ART outcomes

#### Pooled unadjusted estimates of recent exposure to all forms of violence and ART use among female sex workers

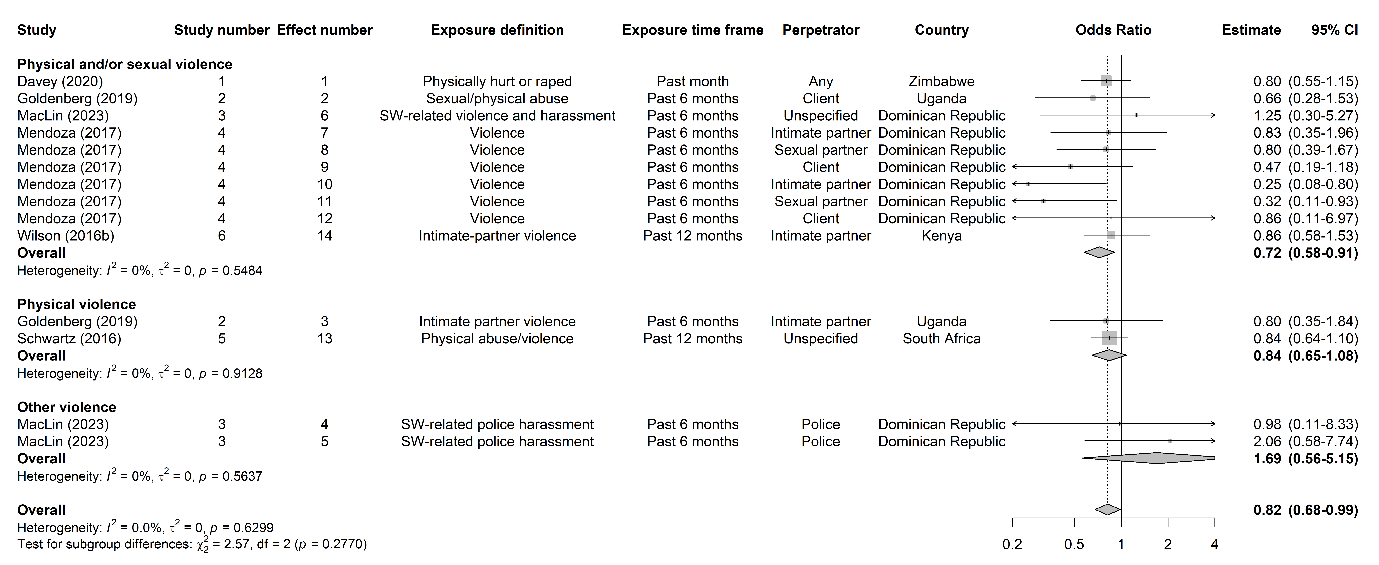

#### Pooled unadjusted estimates of lifetime exposure to all forms of violence and ART use among female sex workers

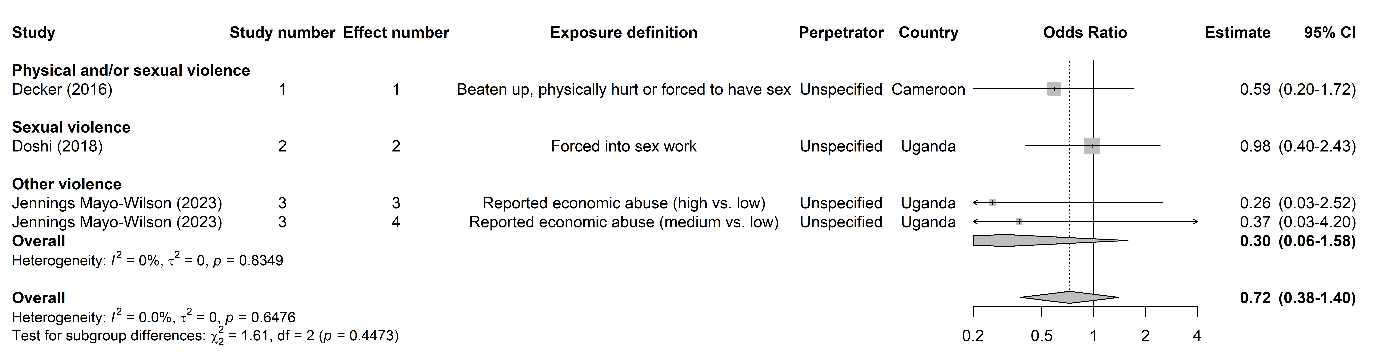

#### Pooled adjusted estimates of recent exposure to all forms of violence and ART use among female sex workers

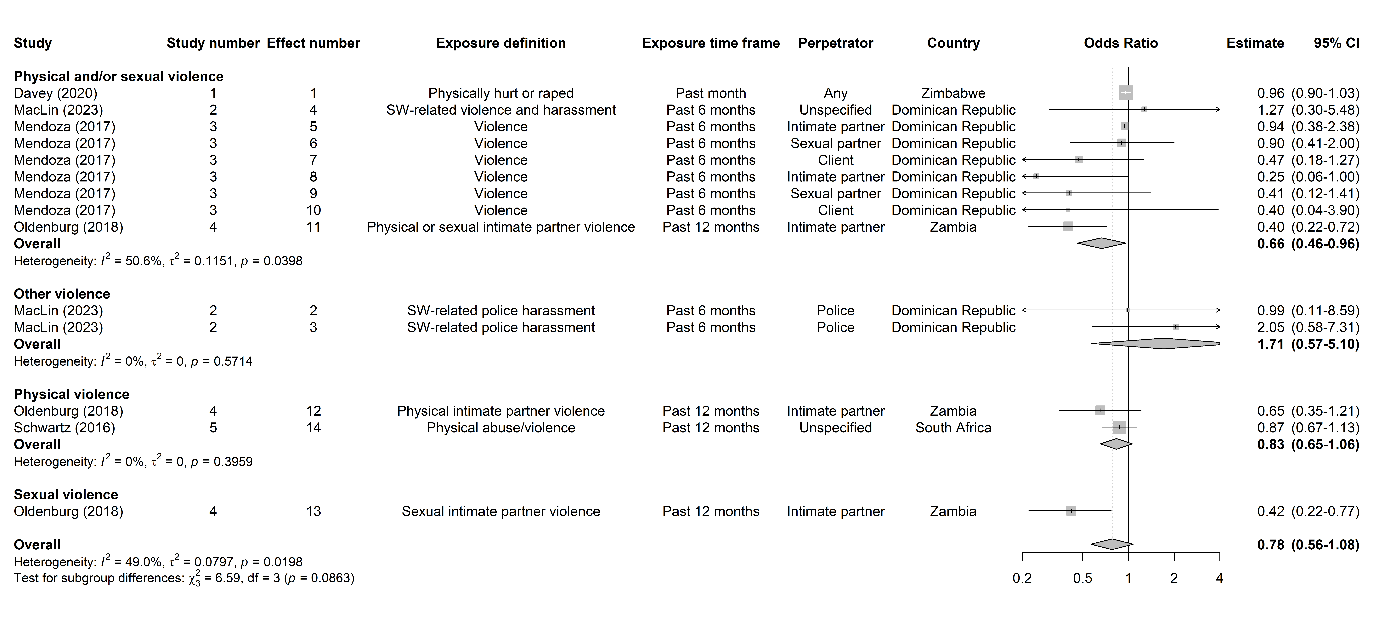

#### Pooled adjusted estimates of lifetime exposure to all forms of violence and ART use among female sex workers

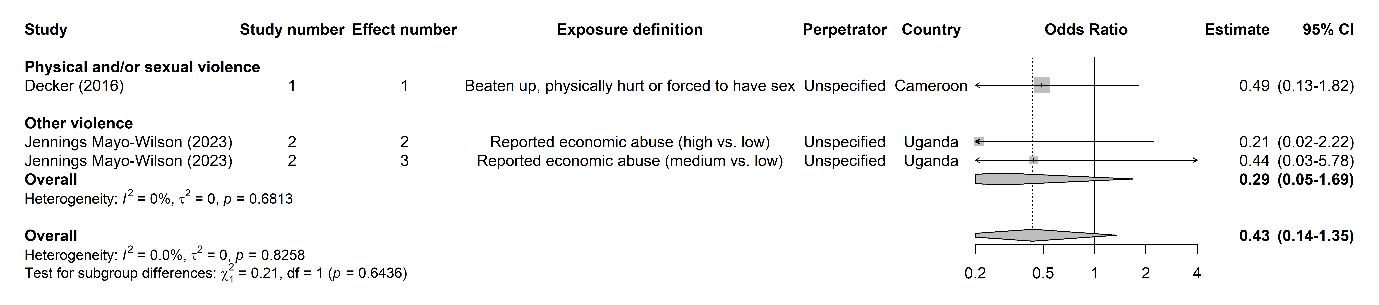

#### Pooled combined unadjusted and adjusted estimates of recent exposure to all forms of violence and ART use among female sex workers

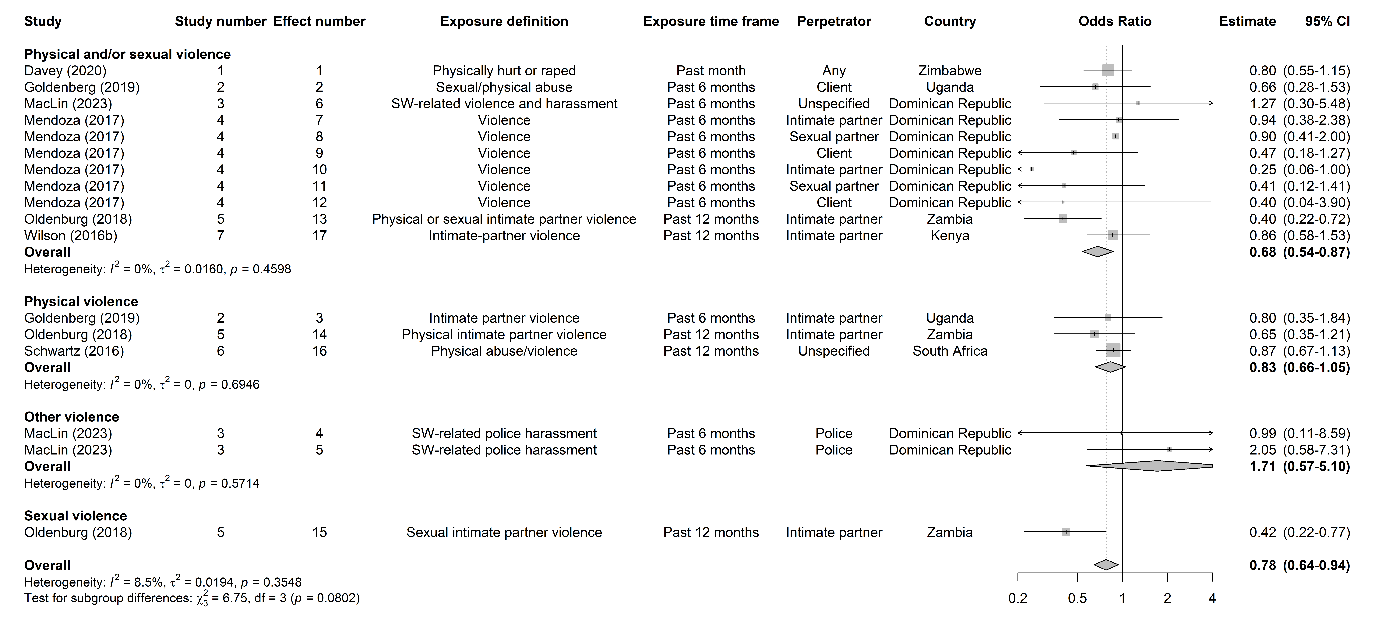

#### Pooled combined unadjusted and adjusted estimates of lifetime exposure to all forms of violence and ART use among female sex workers

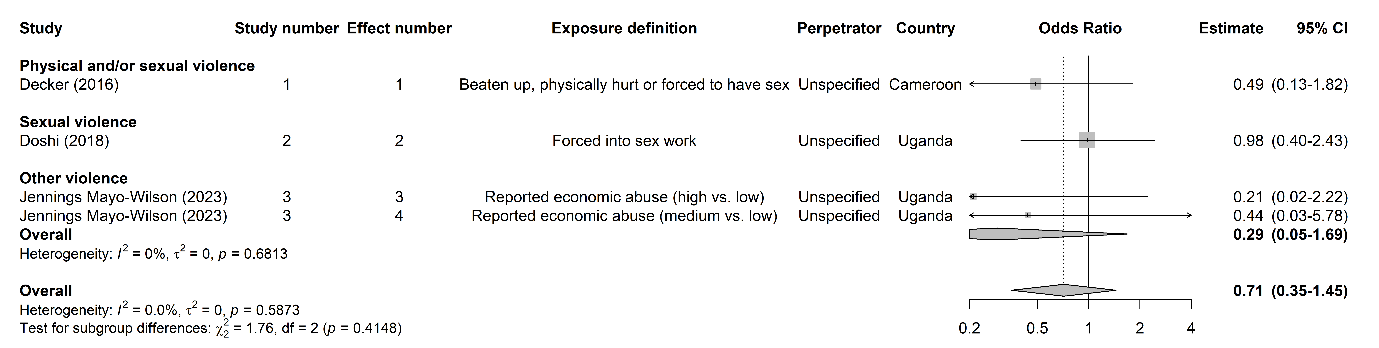

#### Pooled unadjusted estimates of recent exposure to all forms of violence and ART adherence among female sex workers

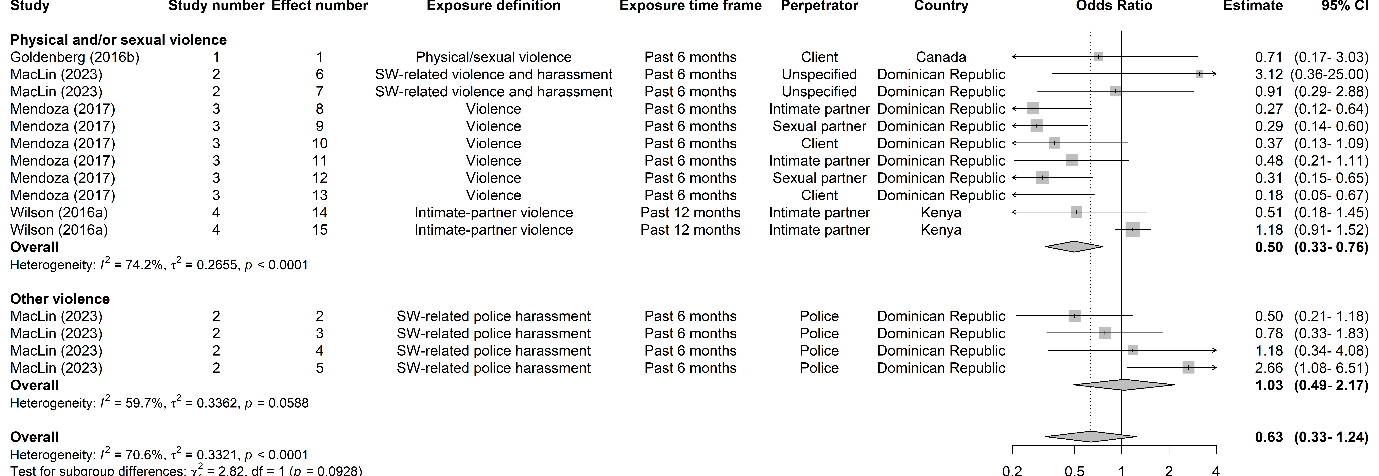

#### Pooled adjusted estimates of recent exposure to all forms of violence and ART adherence among female sex workers

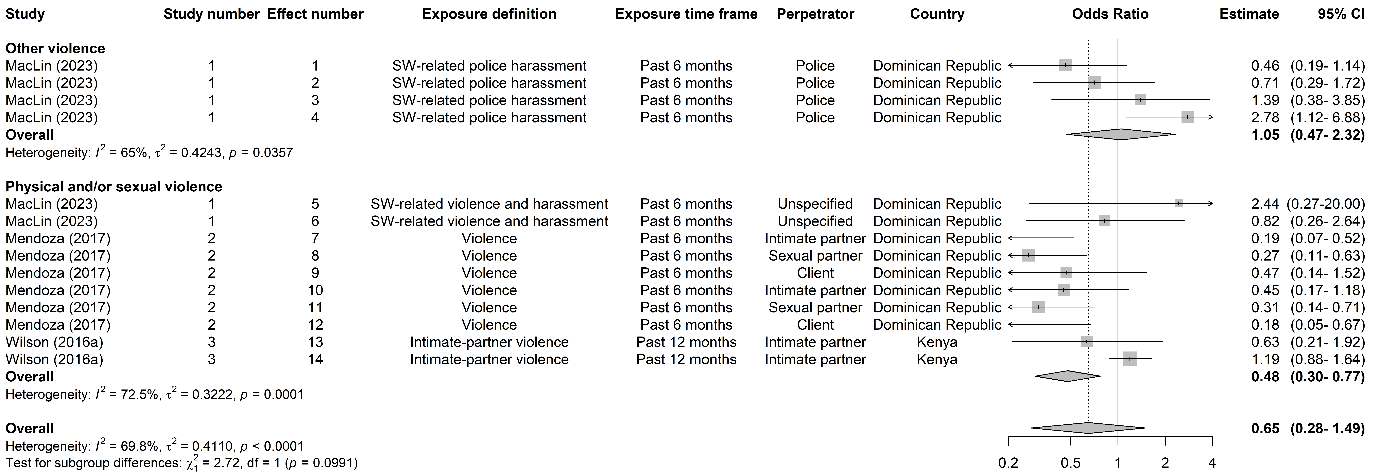

#### Pooled combined unadjusted and adjusted estimates of recent exposure to all forms of violence and ART adherence among female sex workers

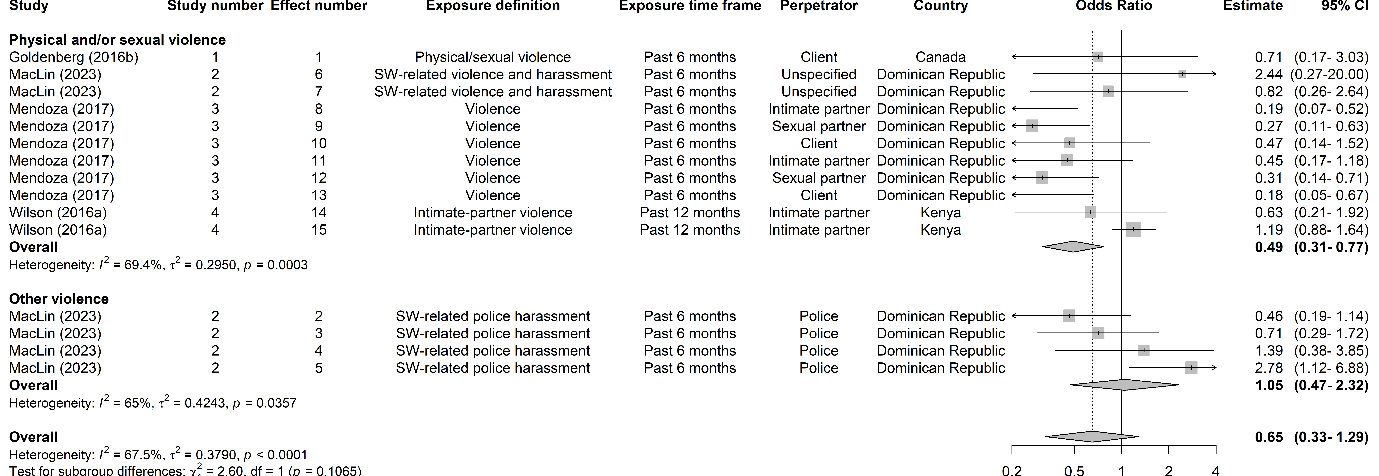

### Appendix 25. Forest plots of violence and HIV viral suppression

#### Pooled unadjusted estimates of recent exposure to all forms of violence and HIV viral suppression among female sex workers

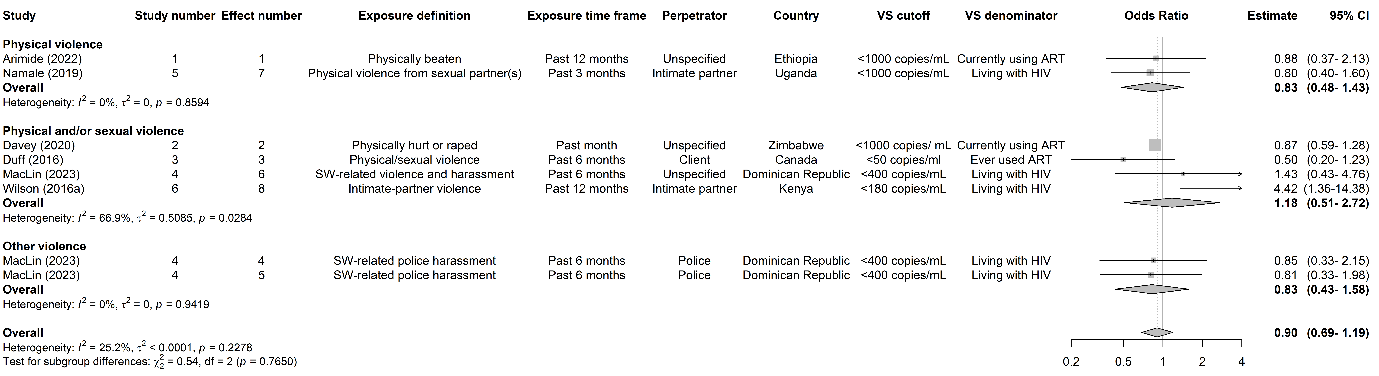

#### Pooled unadjusted estimates of lifetime exposure to all forms of violence and HIV viral suppression among female sex workers

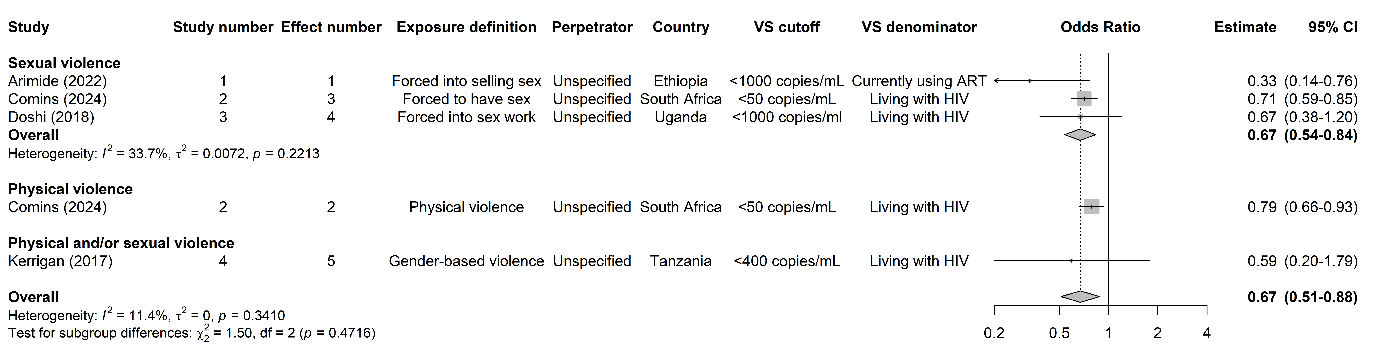

#### Pooled adjusted estimates of lifetime exposure to all forms of violence and HIV viral suppression among female sex workers

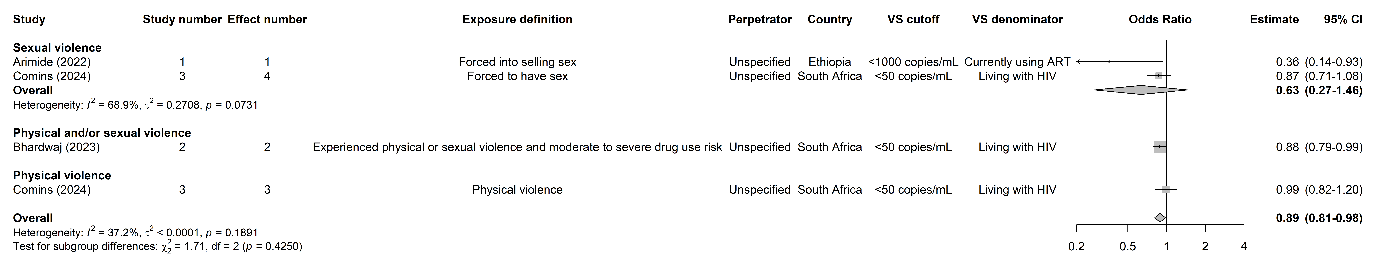

#### Pooled combined unadjusted and adjusted estimates of recent exposure to all forms of violence and HIV viral suppression among female sex workers

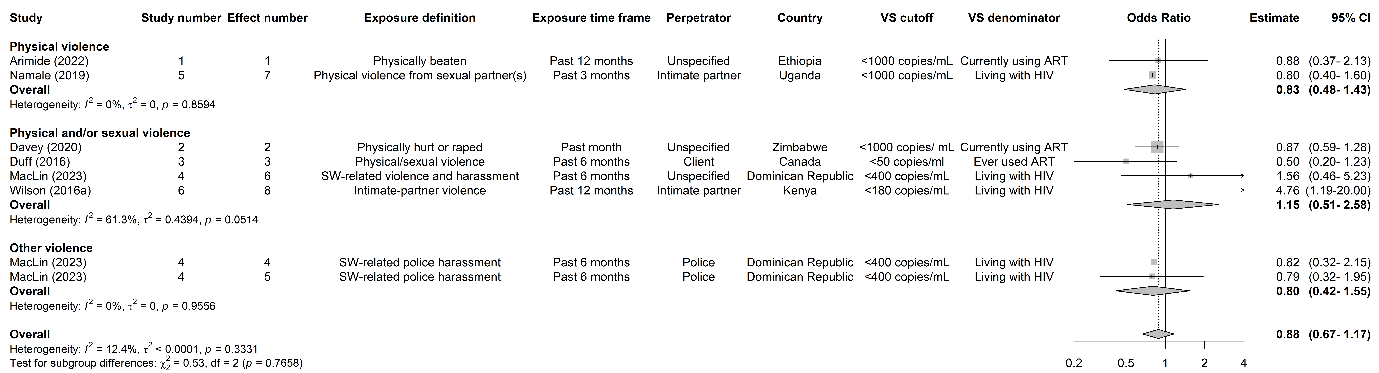

#### Pooled combined unadjusted and adjusted estimates of lifetime exposure to all forms of violence and HIV viral suppression among female sex workers

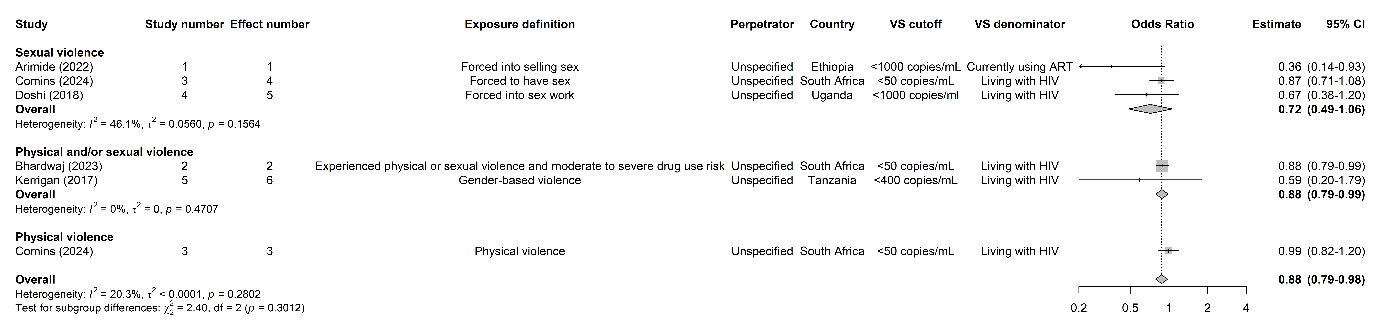

### Appendix 26. Subgroup analyses

#### Subgroup analyses of recent exposure to all forms of violence and HIV infection among female sex workers

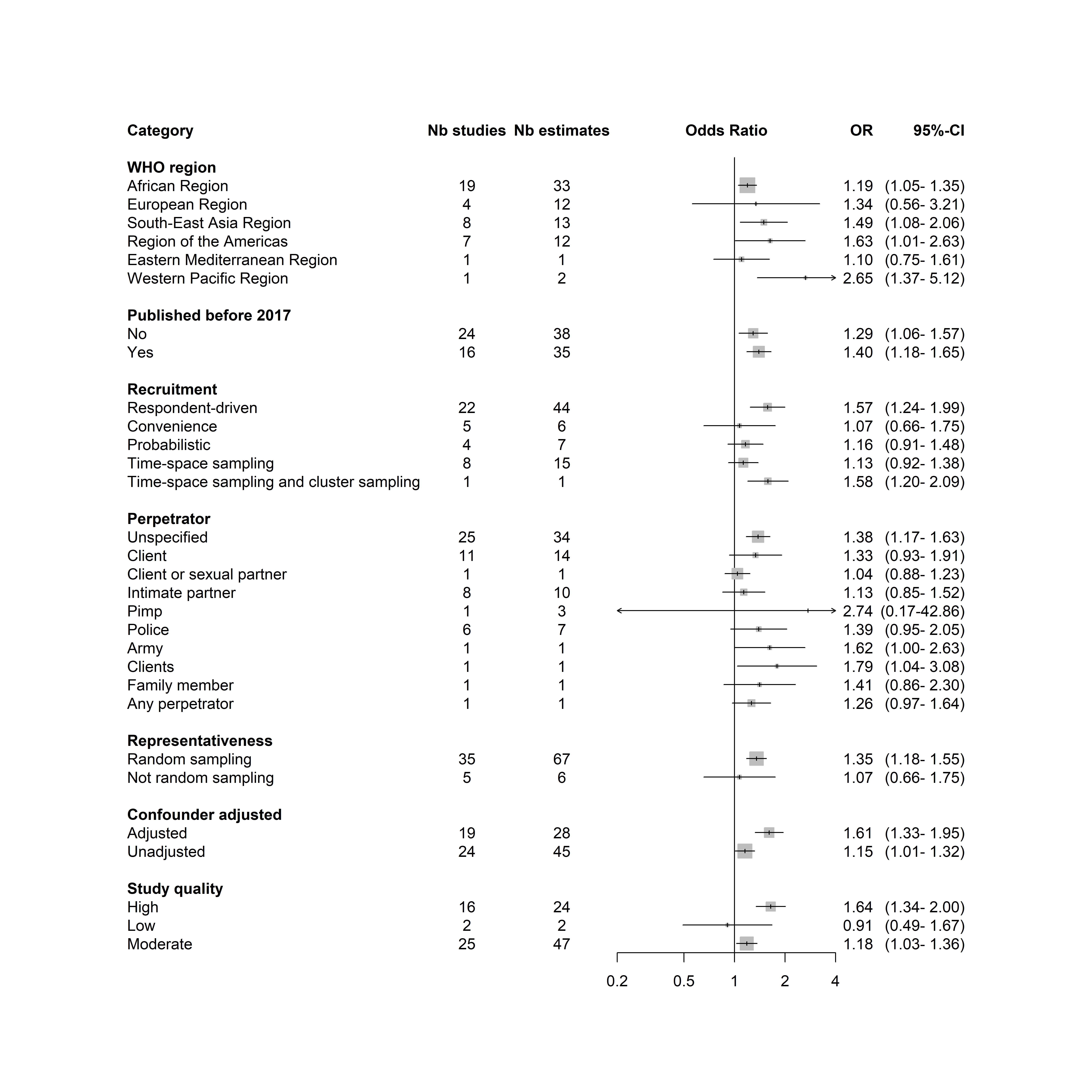

#### Subgroup analyses of lifetime exposure to all forms of violence and HIV infection among female sex workers

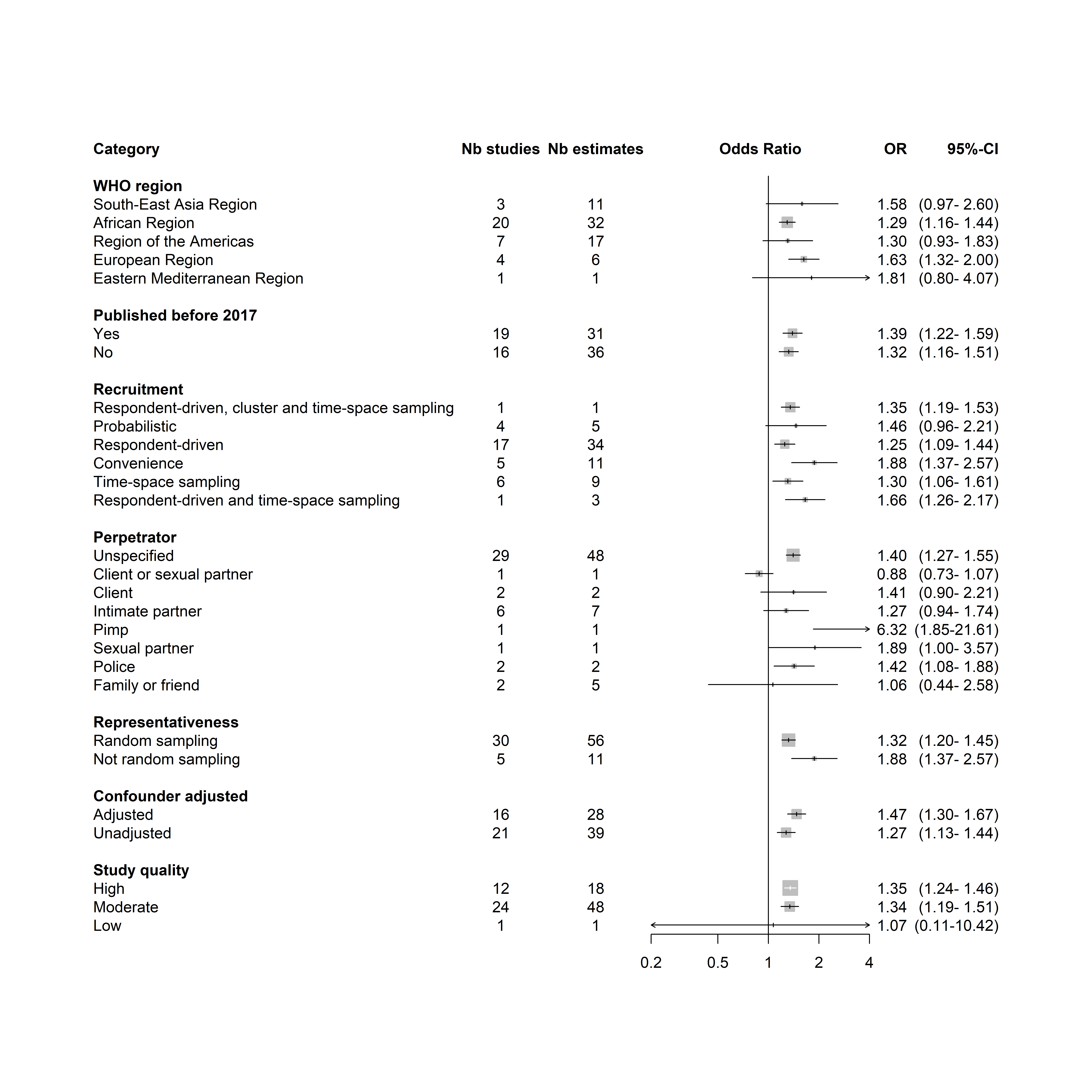

### Appendix 27. Publication asymmetry

For prevalent HIV infection, we observed asymmetry in the funnel plot of the associations with recent experiences of any type of violence, which was confirmed by the Egger’s test (P=0.001). When stratified by violence type, this asymmetry was most evident for recent sexual (P<0.001) or physical violence (P=0.015). There was no evidence of asymmetry for associations between lifetime experiences of violence and prevalent HIV infection (P=0.260). We did not detect asymmetry for estimates of the associations between lifetime violence and HIV testing (P=0.780), and there were fewer than 10 estimates for recent violence. We found evidence of asymmetry for associations between recent violence and ART use from funnel plots and Egger’s tests (P=0.013). There were fewer than 10 estimates for recent and lifetime violence and HIV incidence, ART adherence and HIV viral suppression, so we could not reliably assess for asymmetry.

#### Recent and lifetime exposure to all forms of violence and prevalent HIV infection

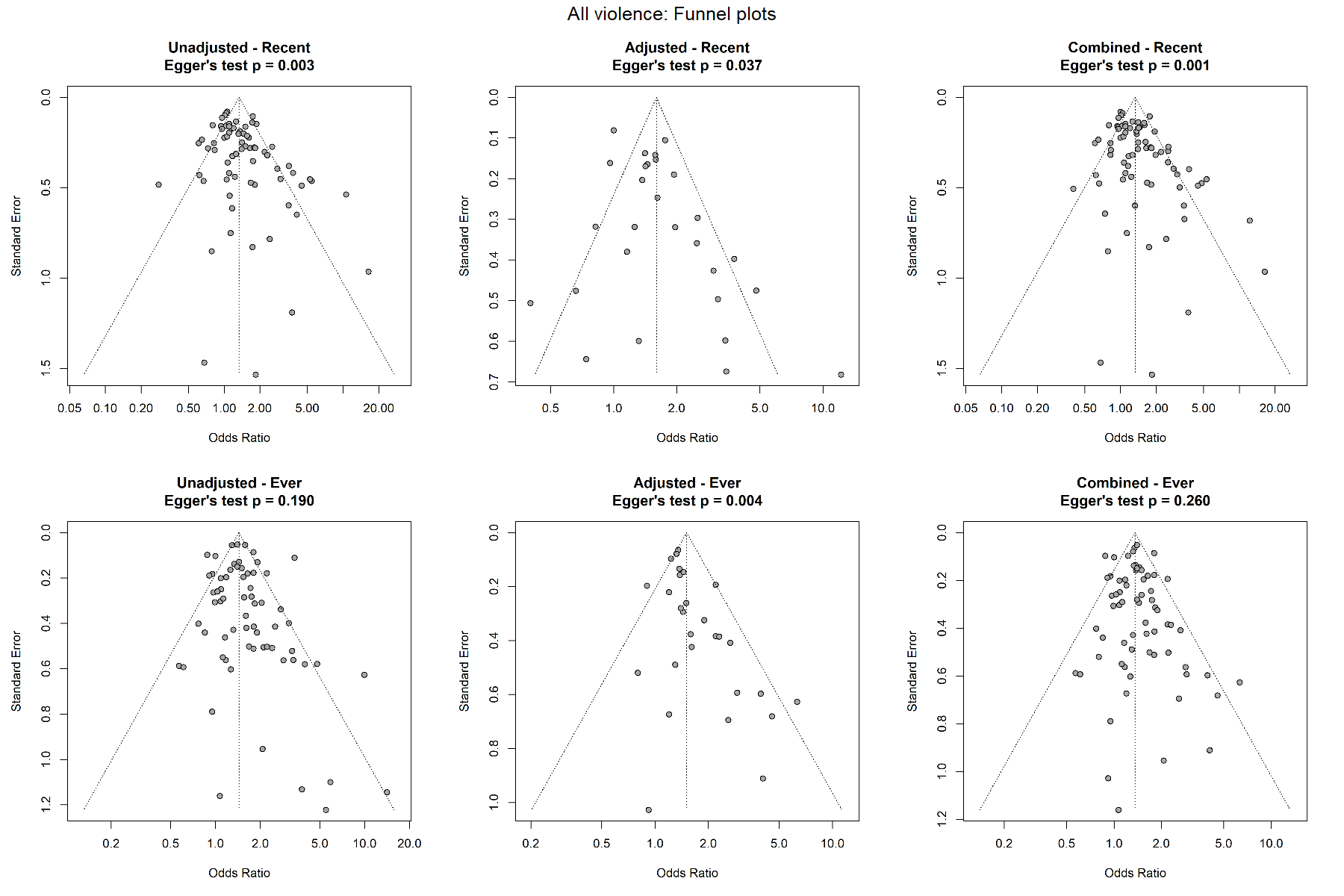

#### Recent and lifetime experiences of physical violence and prevalent HIV infection

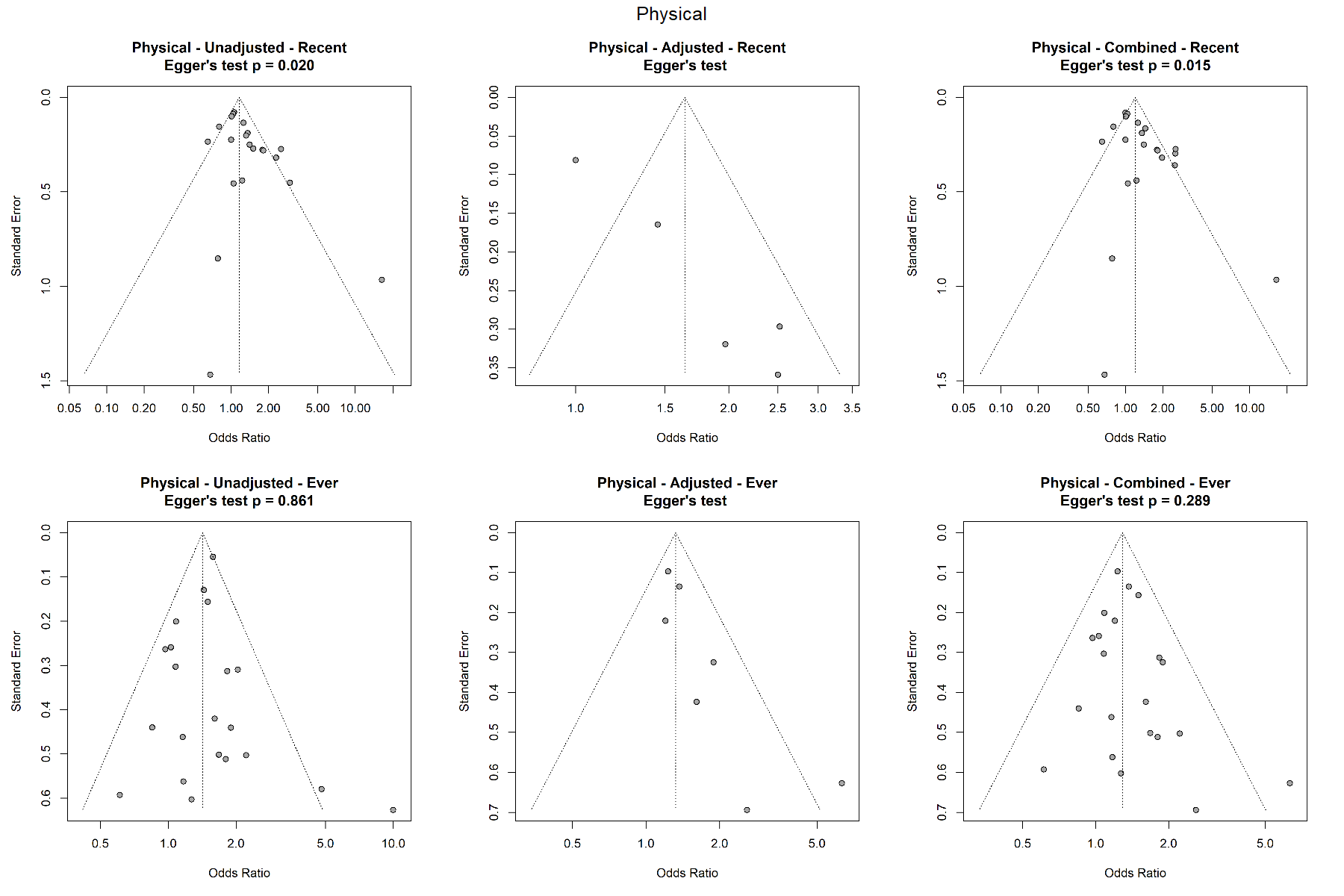

#### Recent and lifetime experiences of sexual violence and prevalent HIV infection

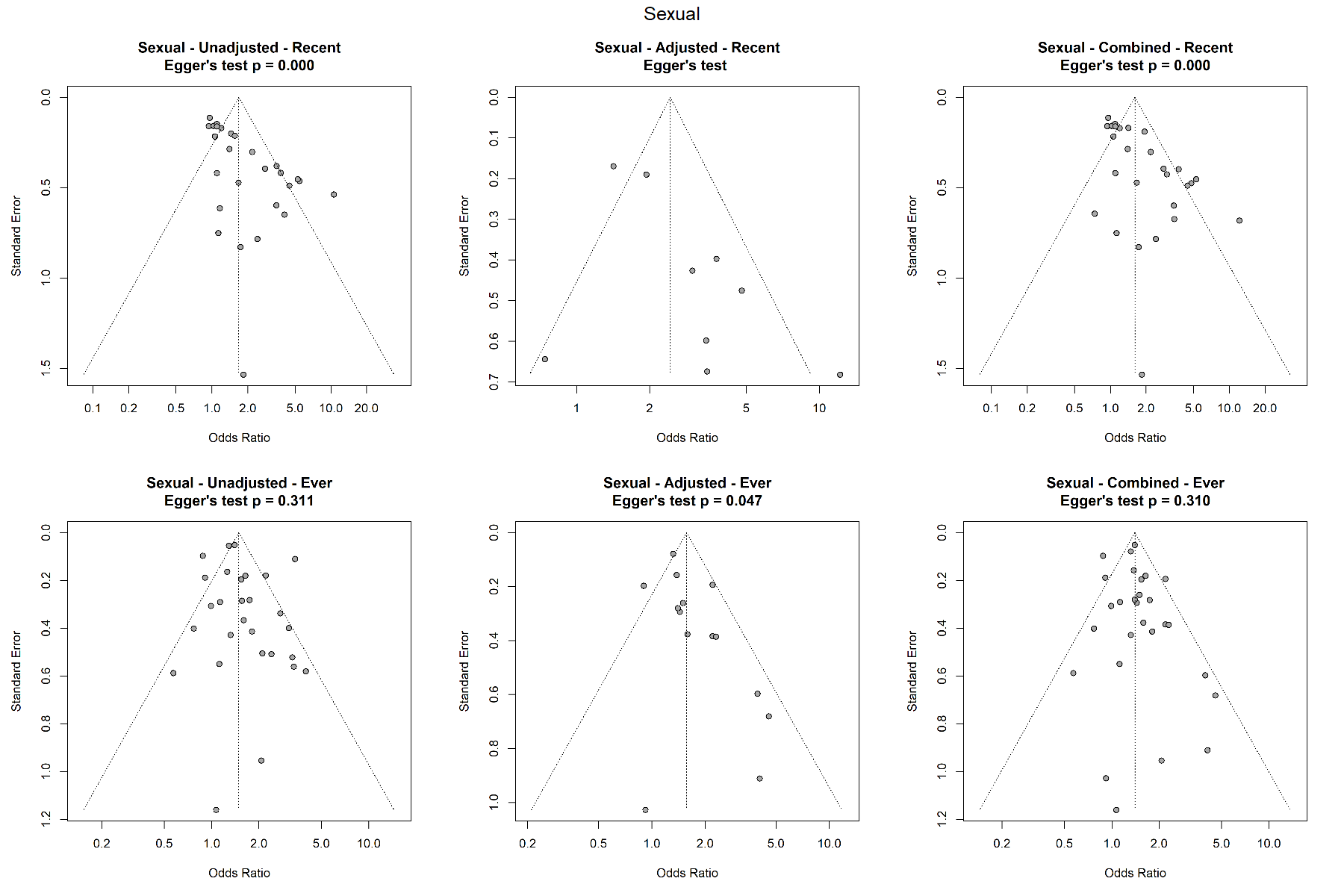

#### Recent and lifetime experiences of physical and/or sexual violence and prevalent HIV infection

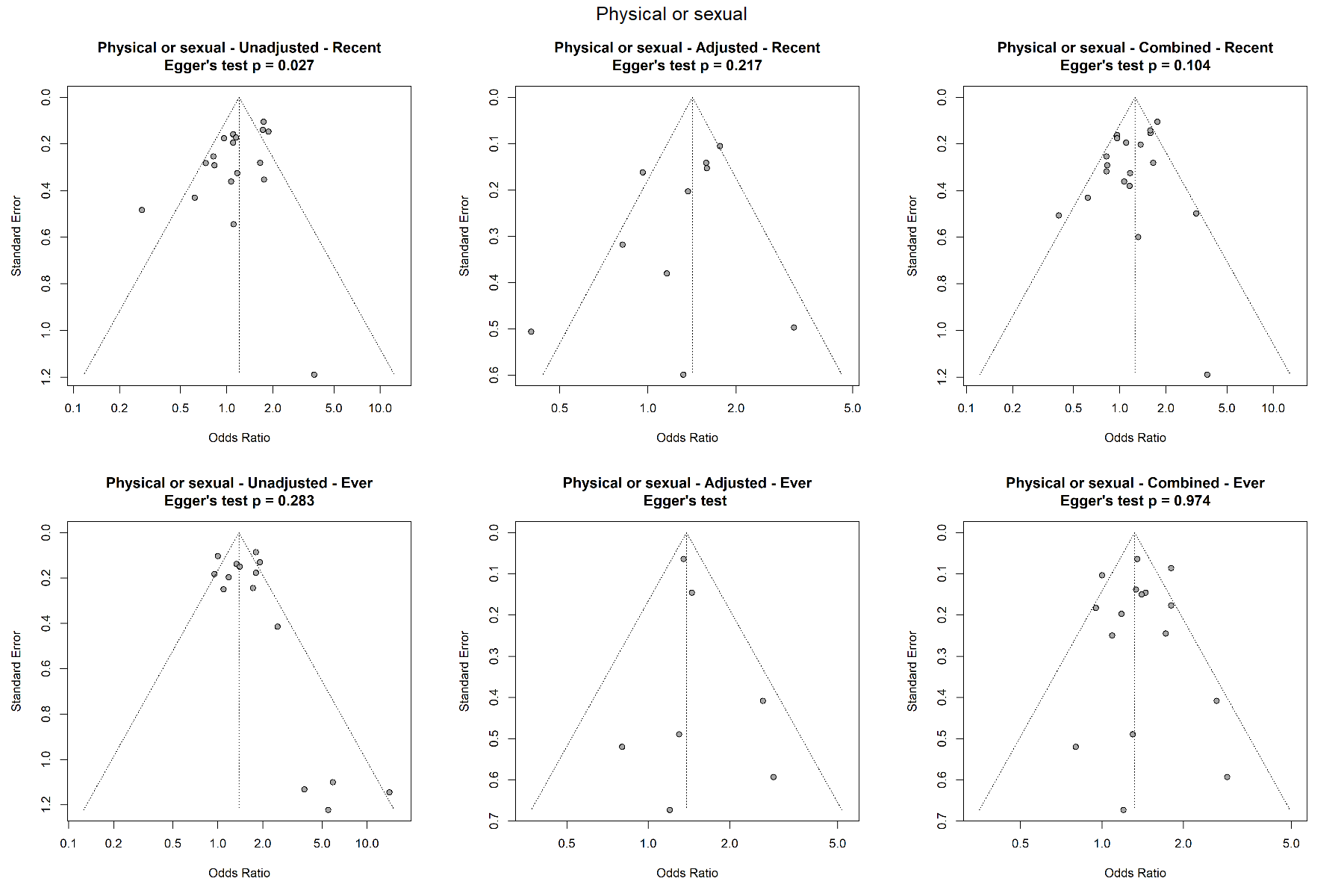

#### Recent and lifetime experiences of all forms of violence and HIV testing

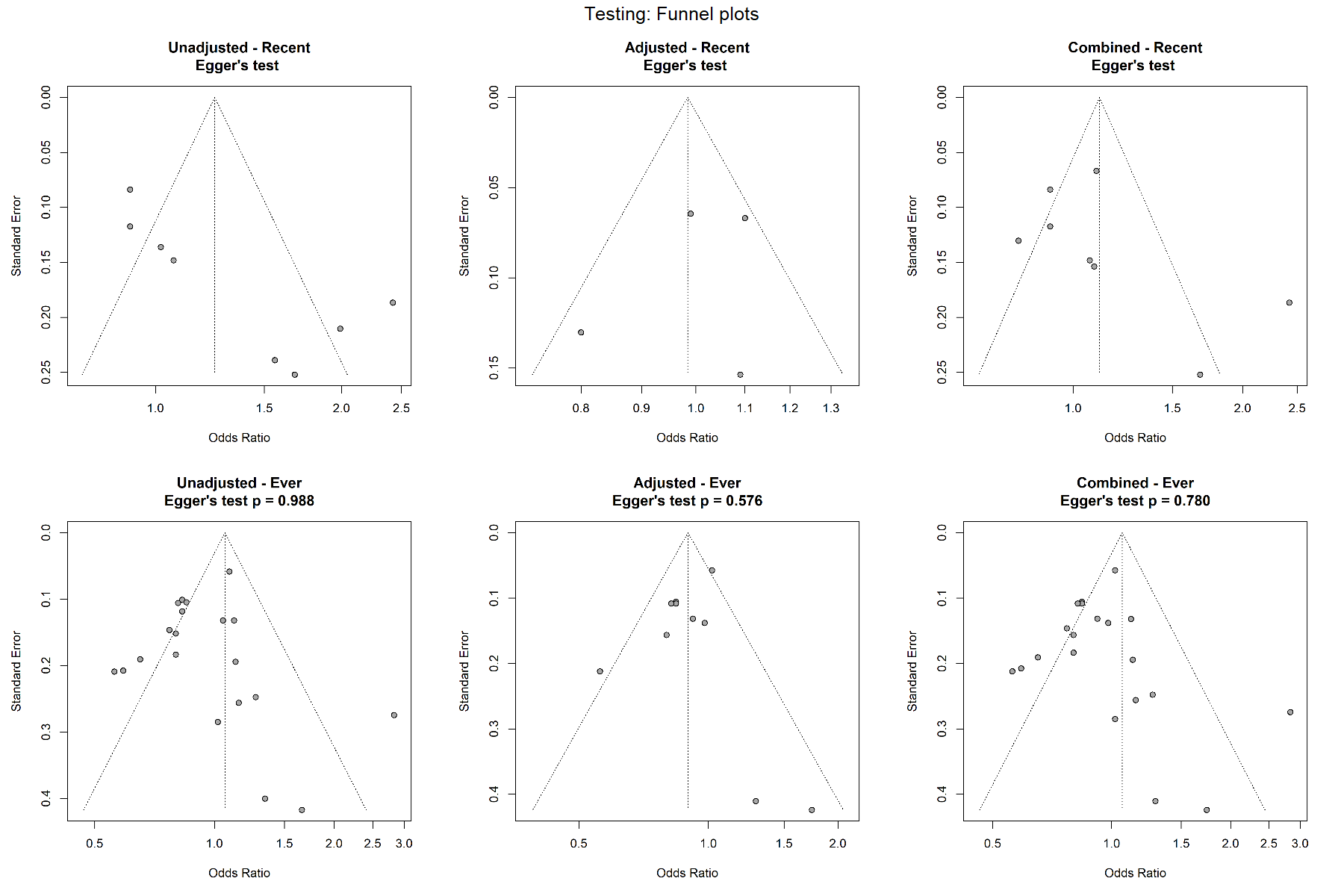

#### Recent and lifetime experiences of all forms of violence and ART use

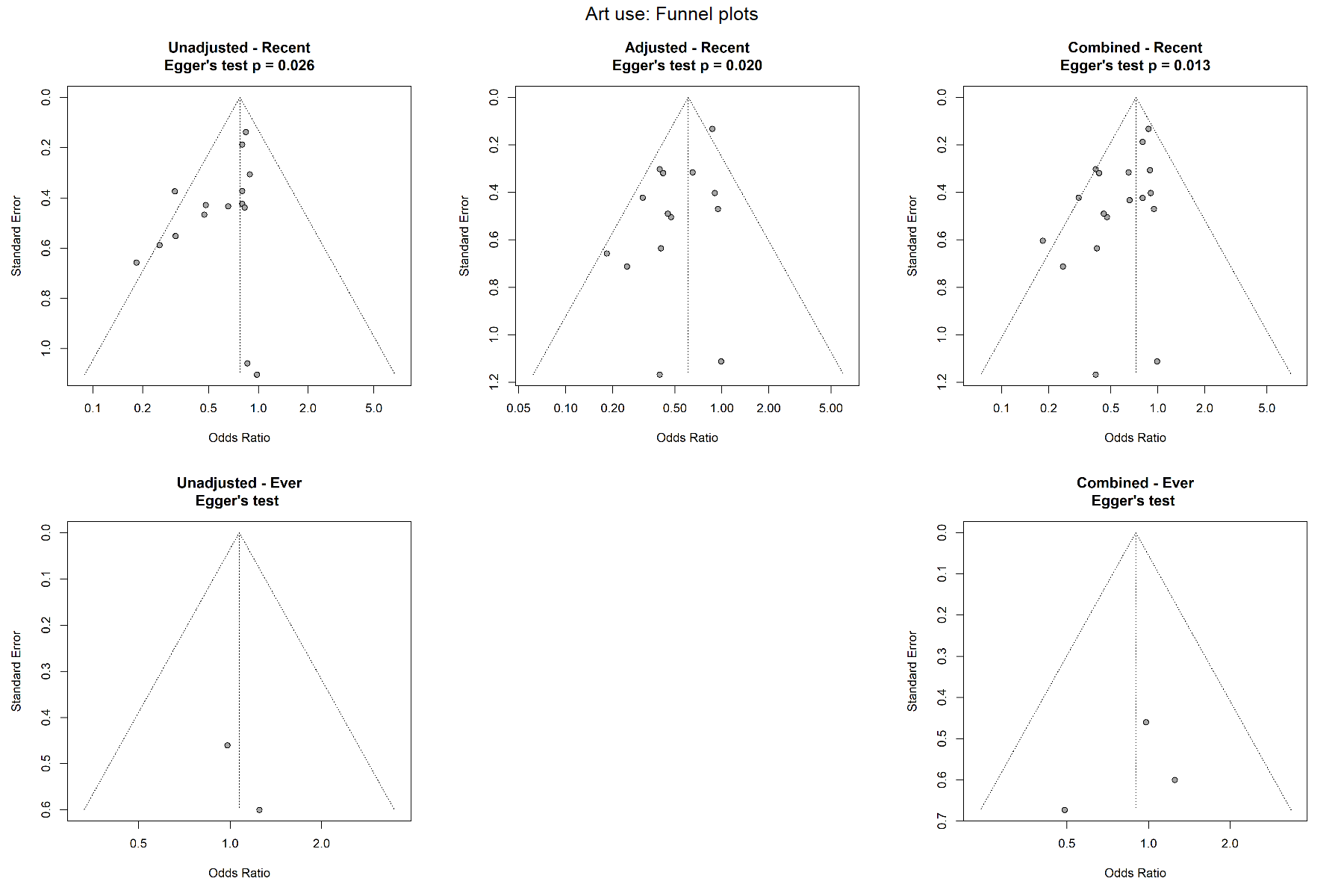

#### Recent experiences of all forms of violence and ART adherence

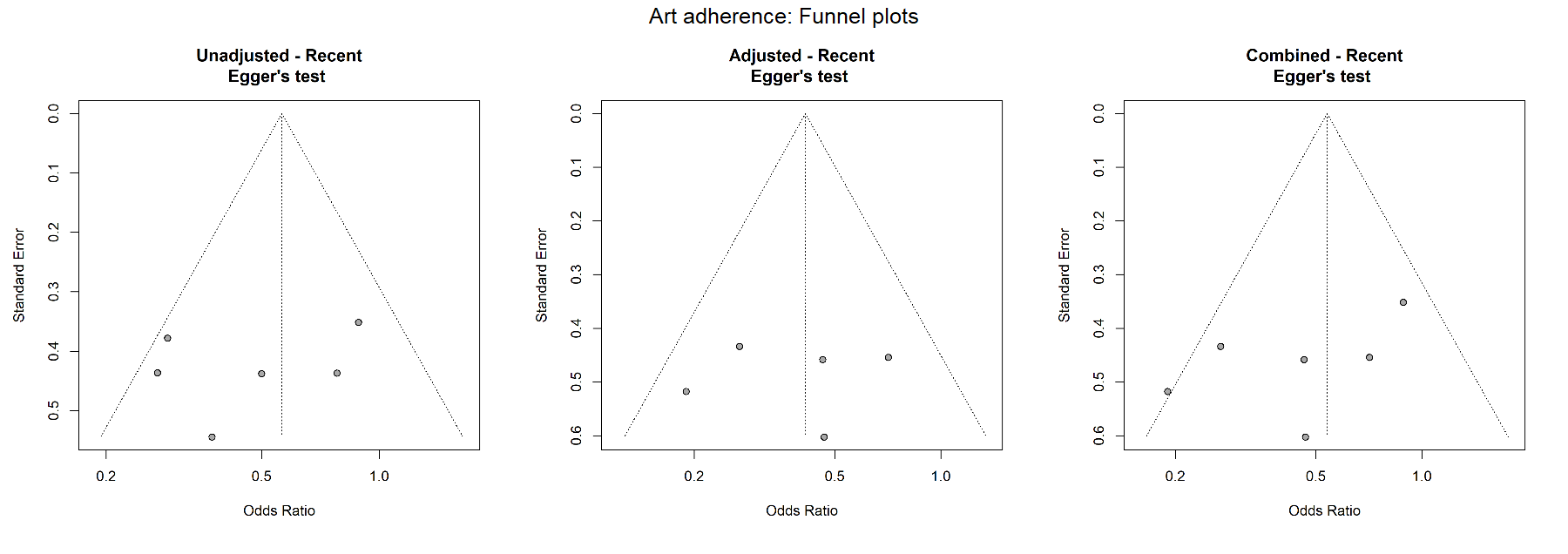

#### Recent and lifetime experiences of all forms of violence and HIV viral suppression

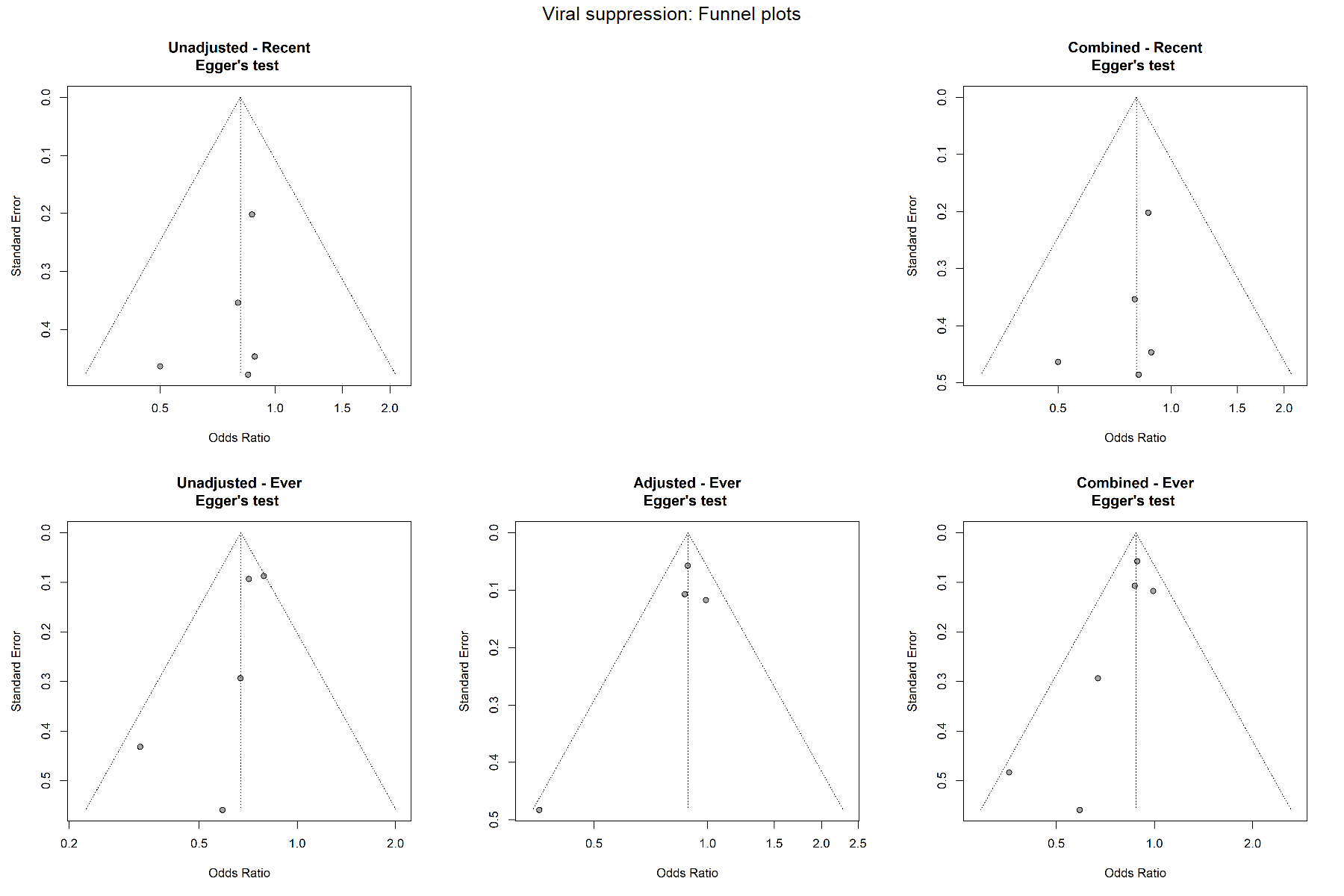

### Appendix 28. Sensitivity analyses of RHO

#### Sensitivity analyses of all forms of violence and HIV outcomes for RHO=0.4, RHO=0.6 and RHO=0.8

| **Outcome** | **Timeframe** | **RHO = 0.4, OR (95% CI)** | **RHO = 0.6, OR (95% CI)** | **RHO = 0.8, OR (95% CI)** |
| --- | --- | --- | --- | --- |
| Prevalent HIV infection | Recent | 1.34 (1.17–1.52) | 1.33 (1.17–1.51) | 1.32 (1.16–1.49) |
| Prevalent HIV infection | Lifetime | 1.35 (1.24–1.47) | 1.36 (1.24–1.49) | 1.38 (1.22–1.55) |
| HIV testing | Recent | 1.12 (0.86–1.46) | 1.11 (0.86–1.45) | 1.11 (0.86–1.43) |
| HIV testing | Lifetime | 1.07 (0.83–1.37) | 1.06 (0.83–1.37) | 1.06 (0.82–1.37) |
| ART use | Recent | 0.76 (0.63–0.93) | 0.78 (0.64–0.94) | 0.74 (0.57–0.97) |
| ART use | Lifetime | 0.71 (0.35–1.42) | 0.71 (0.35–1.45) | 0.72 (0.35–1.47) |
| ART adherence | Recent | 0.65 (0.33–1.30) | 0.65 (0.33–1.29) | 0.65 (0.32–1.29) |
| HIV viral suppression | Recent | 0.89 (0.67–1.17) | 0.88 (0.67–1.17) | 0.90 (0.63–1.29) |
| HIV viral suppression | Lifetime | 0.88 (0.80–0.97) | 0.88 (0.79–0.98) | 0.87 (0.77–0.99) |
